# Evaluation of cancer incidence among Marines and Navy personnel and civilian workers exposed to contaminated drinking water at USMC Base Camp Lejeune: a cohort study

**DOI:** 10.1101/2024.01.27.24301873

**Authors:** Frank J. Bove

## Abstract

**Background:** Drinking water at U.S. Marine Corps Base Camp Lejeune, North Carolina was contaminated with trichloroethylene and other industrial solvents from 1953 to 1985.

**Methods:** A cohort cancer incidence study was conducted of Marines/Navy personnel who, between 1975 and 1985, began service and were stationed at Camp Lejeune, North Carolina (N=154,821) or Camp Pendleton, California (N=163,484), and civilian workers employed at Camp Lejeune (N=6,494) or Camp Pendleton (N=5,797) between October 1972 and December 1985. Camp Pendleton’s drinking water was not known to be contaminated between 1972 and 1985. Individual-level information on all primary invasive cancers and in-situ bladder cancer diagnosed from 1996 to 2017 was obtained from data linkages with 54 cancer registries in the U.S. Survival methods were used to calculate hazard ratios (HRs) comparing cancer incidence between the Camp Lejeune and Camp Pendleton cohorts. Precision of effect estimates were evaluated using the 95% confidence interval (CI) ratio.

**Results:** Cancers among Camp Lejeune Marines/Navy personnel and civilian workers totaled 12,083 (354/100,000) and 1,563 (1,301/100,000), respectively. Cancers among Camp Pendleton Marines/Navy personnel and civilian workers totaled 12,144 (335/100,000) and 1,416 (1,372/100,000), respectively.

Compared to Camp Pendleton, Camp Lejeune Marines/Navy personnel had adjusted HRs ≥1.20 with 95% CI ratios (CIRs) ≤3 for acute myeloid leukemia (HR=1.38, 95% CI: 1.03, 1.85), all myeloid cancers including polycythemia vera (HR=1.24, 95% CI:1.03, 1.49), myelodysplastic and myeloproliferative syndromes (HR=1.68, 95% CI: 1.07, 2.62), polycythemia vera alone (HR=1.41, 95% CI: 0.94, 2.11), cancers of the esophagus (HR=1.27, 95% CI: 1.03, 1.56), larynx (HR=1.21, 95% CI: 0.98, 1.50), soft tissue (HR=1.21, 95% CI: 0.92, 1.59) and thyroid (HR=1.22, 95% CI: 1.03, 1.45). Compared to Camp Pendleton, Camp Lejeune civilian workers had adjusted HRs ≥1.20 with 95% CIRs ≤3 for all myeloid cancers including polycythemia vera (HR=1.40, 95% CI: 0.83, 2.36), squamous cell lung cancer (HR=1.63, 95% CI: 1.10, 2.41) and female ductal breast cancer (HR=1.32, 95% CI:1.02, 1.71). Sensitivity analyses indicated that confounding bias due to unmeasured risk factors (e.g., smoking and alcohol consumption) is unlikely to significantly impact the findings.

**Conclusion:** Increased risks of several cancers were observed among Marines/Navy personnel and civilian workers likely exposed to contaminated drinking water at Camp Lejeune compared to personnel at Camp Pendleton.

## Background

Distribution system drinking water samples collected between 1980 and 1985 at United States Marine Corps (USMC) Base Camp Lejeune, North Carolina found industrial solvents in the drinking water supplied by two of the base’s eight treatment plants. Each drinking water treatment plant served a different area of the base. The Tarawa Terrace (TT) treatment plant began operating in 1952 and served approximately 1,850 family housing units. The TT distribution system was contaminated by an off-base dry-cleaning business. Tetrachloroethylene (PCE) was the primary contaminant in the TT distribution system with measured concentrations of 104 micrograms per liter (µg/L) in July 1982 and a maximum level of 215 µg/L in January 1985. Much lower levels of trichloroethylene (TCE), trans-1,2-dichloroethylene (DCE), and vinyl chloride occurred in the distribution system due to PCE degradation in groundwater [1].

The Hadnot Point (HP) treatment plant began operation in 1942 and served the base’s “mainside” including most of the workplaces, a majority of the bachelor’s quarters (“barracks”), a small number of family housing units, field training areas (via mobile “water buffaloes”) and eating establishments. The HP distribution system was contaminated by on-base sources – leaking underground storage tanks, industrial area spills, and waste disposal sites. TCE and PCE were the primary contaminants, with maximum measured levels in the distribution system of 1,400 µg/L and 100 µg/L, respectively during 1982. A TCE concentration of 1,148 µg/L was measured in drinking water from the HP treatment plant in January 1985. Also detected in the drinking water at the HP treatment plant during 1984 and/or 1985 were benzene, from fuel spills and leaks, and DCE and vinyl chloride from the degradation of PCE and TCE in ground water [2].

The Holcomb Boulevard (HB) treatment plant began operation in 1972 and served approximately 2,100 family housing units and a bachelor officer quarters (BOQ). The HB service area was uncontaminated except for intermittent dry periods when the HP system provided supplementary water. During a two-week period starting in late-January 1985, the HB plant was shut down for repairs and the HP system provided water to the HB service area [2].

No drinking water samples for volatile organic compounds were collected at Camp Lejeune prior to 1980, and there were a limited number of samples taken between 1982 and 1985. Therefore, ATSDR conducted historical reconstruction modeling to estimate the monthly average contaminant levels in the TT and HP distribution systems. Details of the methodology have been summarized elsewhere [1–2]. Based on historical reconstruction modeling estimates, the TT and HP systems were contaminated by the mid-1950s. The highly contaminated supply wells serving the TT and HP systems were shut down by mid-February 1985, although levels of benzene above its maximum contaminant level (MCL) of 5 µg/L were detected on 11/19/1985 (2,500 µg/L) and on 12/10/1985 (38 µg/L) in the HP distribution system. In each system, water from supply wells was mixed together at the treatment plant prior to distribution. Contamination levels in each system varied depending on the wells in use, their levels of contamination, and their pumpage rates [1–2].

Estimated monthly average concentrations of PCE in the TT distribution system between January 1975 and February 1985 ranged from 0 to 158 μg/L with a median of about 85 μg/L [1]. Estimated monthly average concentrations of TCE in the HP distribution system between January 1975 and February 1985 ranged from 0 to 783 μg/L, with a median level of about 366 μg/L [2]. In addition, estimated monthly average levels of PCE and vinyl chloride in the HP distribution system between January 1975 and February 1985 ranged from 0 to 39 μg/L and 0 to 67 μg/L, respectively, with medians of the estimates of 15 μg/L and 22 μg/L, respectively [2].

The United States Environmental Protection Agency (EPA) MCLs are 5 µg/L for TCE, PCE, and benzene; 2 µg/L for vinyl chloride; and 100 µg/L for DCE. EPA and the International Agency for Research on Cancer (IARC) classified TCE as a human carcinogen [3–5]. The EPA classified PCE as a “likely human carcinogen” [6] and IARC classified PCE as “probably carcinogenic to humans” [4–5]. Both benzene and vinyl chloride are known human carcinogens [7–9]. The carcinogenicity of DCE is not classified by EPA.

The drinking water exposures at Camp Lejeune include contributions to total internal body dose from three routes: ingestion, inhalation and dermal. A Marine in training may consume as much as 6 liters/day of drinking water [10]. The combined dose from the inhalation and dermal routes may be as high or higher than the dose from the ingestion route. For example, an internal dose via inhalation to TCE during a 10-minute shower may equal the internal dose via the ingestion of 2 liters of TCE-contaminated drinking water [11].

The ATSDR previously conducted cohort mortality (not cancer incidence) studies of Camp Lejeune Marines/Navy personnel and civilian workers [12–13], and a case-control study of male breast cancer incidence among Camp Lejeune Marines [14]. The mortality studies compared Marines and civilian workers at the base from 1975 to 1985 and 1973 to 1985, respectively, with similar cohorts over the same periods at USMC Base Camp Pendleton, California. Both cohort studies found elevated risks of mortality from cancers of the kidney, rectum, lung, prostate, leukemias, and multiple myeloma [12–13]. Male breast cancer incidence was elevated in the case-control study comparing Camp Lejeune Marines with Marines at other bases [14].

Based on the published ATSDR studies at Camp Lejeune as well as a literature review of occupational and environmental studies conducted elsewhere, an ATSDR report assessed the strength of the evidence supporting causality of cancers from exposures to TCE, PCE, vinyl chloride, and benzene [15]. The assessment integrated findings from ATSDR’s Camp Lejeune mortality studies and male breast cancer study and studies of other populations exposed occupationally or via drinking water to these chemicals. The assessment found sufficient causal evidence for linking TCE and kidney cancer and non-Hodgkin lymphoma (NHL), and “equipoise and above evidence” (i.e., evidence that was as likely as not or greater, but less than sufficient evidence) for TCE and multiple myeloma, leukemias, and liver cancer. Sufficient causal evidence was found for PCE and bladder cancer, and “equipoise and above evidence” for PCE and NHL. Sufficient causal evidence was found for benzene and NHL and leukemias, and “equipoise and above evidence” for benzene and multiple myeloma. Sufficient evidence was found for associating vinyl chloride and liver cancer.

Two epidemiological studies have evaluated cancer incidence and drinking water exposures to TCE or PCE. A New Jersey study observed associations between NHL and TCE and PCE, and leukemia and TCE [16]. A study in Cape Cod, Massachusetts found associations between PCE and cancers of the lung, bladder, rectum, female breast, and leukemia [17–19].

The purpose of this cancer incidence cohort study of Camp Lejeune Marines/Navy personnel and civilian workers was to determine if being stationed or employed at Camp Lejeune between 1975 and 1985 (Marines/Navy personnel) or between October 1972 and December 1985 (civilian workers), a portion of the period when the drinking water was contaminated, increased the risk of cancer incidence ascertained between 1996 and 2017 compared to being stationed or employed at Camp Pendleton. Camp Pendleton was not known to have contaminated drinking water during the years prior to 1986 [20].

## Methods

### Study Populations

ATSDR obtained quarterly personnel data from the Defense Manpower Data Center (DMDC) for full time civilian workers who were employed during any quarter between October 1972 and December 1985 at Camp Lejeune or Camp Pendleton. The DMDC data did not contain information on part-time employees. The DMDC began collection of personnel data for civilian workers in the last quarter of 1972. The end of the year 1985 was selected because drinking water distribution system samples taken at Camp Lejeune from 1986 onward indicated no contamination above the contaminants’ MCLs. The study included a cohort of 6,494 workers employed at Camp Lejeune and a comparison cohort of 5,797 workers employed at Camp Pendleton, who were known to be alive as of January 1, 1996. The DMDC data included base location of employment (state, city and zip codes), social security number, full name (started in the last quarter of 1981), date of birth, paygrade, education level, race, sex, and occupation code. Based on the DMDC data, the average duration of employment at Camp Lejeune between October 1972 and December 1985 was 56 months.

ATSDR also obtained quarterly personnel data from the DMDC for Marines and Navy personnel stationed at Camp Lejeune and Camp Pendleton for the years 1975 to 1985. Although drinking water contamination preceded 1975, the code for unit (e.g., regiment, battalion, company, etc.), necessary to determine the base where the individual was stationed, was not available in the DMDC database until the second quarter of 1975. In addition to the unit code, the DMDC data included date of birth, marital status, rank (paygrade), date active duty started, military occupation code, education level at the start of service, race, sex, full name, and social security number. The USMC provided a list of the unit codes for the units that were stationed at each base. Based on the DMDC data, Marines/Navy personnel in the Camp Lejeune cohort were stationed at the base on average for 18 months.

The full cohort of Marines/Navy personnel for this study included 211,023 at Camp Lejeune and 224,419 at Camp Pendleton, who were known to be alive as of January 1, 1996. Some members of the full cohort began active duty prior to 1975 when information on base location (i.e., unit code) was not available in the DMDC data. For these Marines/Navy personnel, it would be unknown whether those stationed at Camp Pendleton between 1975 and 1985 were stationed at Camp Lejeune prior to 1975. Since it was not unusual for Marines/Navy personnel to be stationed at both bases, it was likely that some who began active duty prior to 1975 and were stationed at Camp Pendleton between 1975 and 1985, were stationed at Camp Lejeune prior to 1975. To address this problem, a subgroup of the full cohort was identified consisting of Marines/Navy personnel who began active duty between 1975 and 1985 when information on base location was available in the DMDC database. This subgroup consisted of 154,821 at Camp Lejeune and 163,484 at Camp Pendleton, who were known to be alive as of January 1, 1996. Comparisons between the Camp Lejeune and Camp Pendleton subgroup are the main focus of the evaluation of cancer incidence among Marines and Navy personnel.

Camp Pendleton Marines/Navy personnel and civilian workers were chosen as the comparison groups in this study because the base’s finished drinking water was not known to be contaminated prior to 1986 [20]. Moreover, Camp Pendleton’s Marines/Navy personnel and civilian workers were similar to Camp Lejeune in terms of demographics, socioeconomic factors, training activities, personnel trained, and types of civilian employee occupations. Biases due to the “healthy veteran effect” [21–23] or the “healthy worker effect” [24], or due to unmeasured confounders, should be reduced by having comparison cohorts with similar risk factor characteristics as the Camp Lejeune cohorts.

### Cancer Ascertainment

Linkage between the cohort data and a commercial tracing service was used to correct discrepant names, social security numbers, and dates of birth and to obtain the most recent five residential street addresses and vital status. Vital status and date of death were obtained via linkage with the Social Security Administration (SSA) Data for Epidemiological Researchers and the National Death Index. The resulting information was used in the data linkages with cancer registries.

Individual-level information on all primary invasive cancers and in situ bladder cancer from 1996 to 2017 was obtained from data linkages with 49 state cancer registries, the cancer registries of Puerto Rico and the Pacific Islands, the District of Columbia cancer registry, and the cancer registries at the Department of Defense (DOD) and the Department of Veterans Affairs (VA). Due to state law restrictions requiring consent of the living patient, the West Virginia Cancer Registry provided aggregate data on specific cancers by age group, sex, whether Marine or civilian employee, and base stationed or employed. The aggregate data did not distinguish the 1975-1985 subgroup of Marines/Navy personnel from the full cohort. The Kansas Cancer Registry had a similar state law restriction but was able to obtain consent from, and provide individual-level data for, most of the patients that matched to the cohorts. For those who matched but did not provide consent, the Kansas Cancer Registry provided similar aggregate data as the West Virginia Cancer Registry.

The start of follow-up was January 1, 1996, because all registries were operating by 1996 (some registries were not operating prior to 1996). December 31, 2017 was chosen as the end date for data collection from the registries because some of the registries did not have complete and verified data beyond 2017 at the time the linkages were scheduled to be performed. In situ bladder cancers were included in the study “…because the information needed to distinguish between *in-situ* and invasive bladder cancers is not always available or reliable” (see https://www.cdc.gov/cancer/uscs/technical_notes/data_sources/incidence.htm).

All cancer registries except the DOD cancer registry utilized the same linkage software (Match*Pro, a Java-based application developed by Information Management Services, Inc). Similar manual review procedures were performed at all the registries except the VA and DOD registries which did not perform manual review. The matching parameters used by the linkage software were first, middle, and last name (using a Soundex algorithm that matches names that have similar pronunciation but may have different spellings) for both the cancer registry data and the cohort personnel data), social security number, date of birth, and street address. Blocking parameters (first name, last name, social security number, and date of birth) were used to limit the number of comparisons to those records for which two or more blocking parameters matched.

The linkage software produced three classes of matches: high quality, uncertain, and non-matches. The thresholds for these three classes were based on pilot tests with three of the cancer registries and were consistent across all linkages. Registries manually reviewed all uncertain matches to identify any missed cases. Most registries also reviewed all high-quality matches for potential false positives. Based on this review, about 0.1% of the high-quality matches were identified as false positives. Many registries also reviewed records in the unmatched category for any false negatives. Once all the cancer data were received, duplicate records were removed.

Cancer registries provided the following information for each matched tumor record: primary site of the cancer, histologic type, laterality, behavior code (benign, in situ, malignant), grade, diagnostic confirmation, cancer stage (Surveillance, Epidemiology and End Results Program (SEER) summary stage-1977 for 1977 to 2000; SEER summary stage-2000 for 2001 to 2017), sequence number, state of diagnosis, age at diagnosis, date of diagnosis, and whether the cancer was identified solely by death certificate (“DCO” case). Histological subtypes were defined using the SEER site recode definitions based on the cancer site and International Classification of Diseases for Oncology, 3^rd^ edition (ICD-O-3) histology codes, updated for hematopoietic codes based on the World Health Organization (WHO) Classification of Tumours of Hematopoietic and Lymphoid Tissues [25]. The histology coding schemes for the histological subtypes are provided in Supplemental file 1, Table S1-1.

### Data Analyses

The analyses focused on comparisons between the Camp Lejeune and Camp Pendleton cohorts. For the Marines/Navy personnel, the analyses focused on comparisons between the Camp Lejeune and Camp Pendleton 1975-1985 subgroup. Analyses of the full cohort of Marines/Navy personnel are presented in Supplemental file 1, Tables S1-2 to S1-4.

Follow-up began on January 1, 1996, and continued until date of death or December 31, 2017, whichever was earlier. Because exposures among the Camp Lejeune cohorts occurred more than 10 years before the start of follow-up, the data analyses did not lag exposures to account for a latency period. Data analyses evaluated each primary cancer site as well as the histological subtypes for some primary cancer sites.

Descriptive analyses included the calculation of standardized incidence ratios (SIRs) for each base and primary cancer site. The sex, race and five-year age-specific cancer incidence statistics for 1999-2017 for the United States and Puerto Rico from the CDC WONDER online database were used as the basis for calculating the SIRs. Poisson regressions comparing the sex, race, and five-year age-specific cancer incidence rates for Camp Lejeune versus Camp Pendleton were conducted as part of the descriptive analyses because comparisons of the SIRs between the two bases could be impacted by residual confounding bias due to differences in the distributions of age, sex and/or race.

To calculate the SIRs and conduct the Poisson regressions, person-years at risk were accumulated during the follow-up period from 1996 to 2017 and were stratified by base, sex, race and 5-year age categories. Person-years at risk were assigned to Camp Lejeune if the individual was stationed or employed at the base anytime between 1975 and 1985 (Marines/Navy personnel) or between October 1972 and 1985 (civilian workers), regardless of whether the individual was also stationed or employed at Camp Pendleton during these periods. Person-years at risk were assigned to Camp Pendleton only if the individual was stationed or employed at that base between 1975 and 1985 (Marines/Navy personnel) or October 1972 to 1985 (civilian workers) and not stationed at Camp Lejeune during these periods.

The aggregate data from the Kansas and West Virginia registries did not identify Marines and Navy personnel belonging to the subgroup, so the aggregate data were only used in the SIR and Poisson regression analyses comparing the Camp Lejeune and Camp Pendleton full cohort. In addition to the individual-level cancer data, a total of 510 cancers from the aggregate data obtained from the West Virginia and Kansas cancer registries were included in the SIR and Poisson regression analyses of the full cohort. For the civilian workers, the SIR and Poisson regression analyses included the individual-level cancer data as well as 21 cancers from the aggregate data obtained from the West Virginia and Kansas cancer registries.

The main analysis evaluated individual-level data using Cox proportional hazards (Cox) regression to estimate hazard ratios (HRs) and 95% confidence intervals (CIs) for each cancer site and histological subtype. Age was the time variable. Marines/Navy personnel, and civilian workers stationed or employed at Camp Lejeune were compared to those stationed or employed at Camp Pendleton. For the analyses of Marines/Navy personnel, the adjusted models included sex, race, rank, and education level (not a high school graduate, high school graduate, college graduate and higher). For the analyses of civilian workers, the adjusted models included sex, race, blue collar work (y/n), and education level. Blue collar work included manual jobs such as maintenance workers, mechanics, construction workers, laundry and dry-cleaning workers, pest control workers and water treatment plant workers. Evaluation of Schoenfeld residuals was used to check the proportional hazards assumption. The Schoenfeld residuals are calculated for all covariates for each individual experiencing an event at a given age and consist of the differences between that individual’s covariate values at the age when the event occurred and the corresponding risk-weighted average of covariate values among all those then at risk at that age. The proportional hazards assumption is met if there is no pattern in the residuals over age.

The main analyses of the Marines/Navy personnel focused on comparisons between the Camp Lejeune and Camp Pendleton 1975-1985 subgroup. Secondary analyses evaluated the full cohort comparing Camp Lejeune and Camp Pendleton. For civilian workers, the main analyses also focused on comparisons between Camp Lejeune and Camp Pendleton.

In the previous Camp Lejeune mortality studies, residential cumulative exposure to each contaminant was evaluated based on linking the estimated monthly concentrations in the TT, HP and HB water systems from the historical reconstruction modeling and Camp Lejeune base family housing records and information on the barrack location of each military unit [12–13]. In this study, cumulative residential exposure to each contaminant was not conducted because drinking water exposures during training and other base activities would likely contribute significantly to overall cumulative exposure. Since information on training and other base activities was not available, the study focused instead on duration of assignment (Marines/Navy personnel) or duration of employment (civilian workers) at Camp Lejeune as a surrogate for overall cumulative exposure. Duration at Camp Lejeune is defined as the number of quarters in the DMDC database an individual is stationed or employed at Camp Lejeune during 1975-1985 for Marines/Navy personnel and during October 1972 and December 1985 for civilian workers. Cox regression analyses using categorical variables for duration were conducted with Camp Pendleton Marines/Navy personnel and civilian workers as the comparison groups.

In the Cox regression analyses, an individual could contribute cancers at more than one cancer site but not more than one per site. For example, if a person had recurrent lung cancer records during the follow-up period, only the first lung cancer during the period was included in the analysis of the lung cancer site. However, an individual could contribute to more than one subtype of a particular cancer site. For example, an individual who had a lung cancer adenocarcinoma histology and later had a lung cancer squamous cell histology would be included in the analysis of each of these histological subtypes.

Information on smoking and alcohol consumption was not available. Occupational history prior to or after active-duty service or employment at Camp Lejeune or Camp Pendleton was also unavailable.

To assess the possible confounding effects of smoking and alcohol consumption, the study evaluated “negative control” diseases that are associated with the unmeasured risk factor (i.e., the potential confounder) but were not known to be associated with the exposures of interest, i.e., exposure to the drinking water contaminants at Camp Lejeune [26]. Negative controls were used to estimate prevalence differences in smoking and alcohol consumption between Camp Lejeune and Camp Pendleton. The negative control diseases for smoking were mortality due to chronic obstructive pulmonary disease (COPD) and cardiovascular disease. Several smoking-related cancers, such as cancers of the lung, larynx, and bladder [27], were included in the study but were not considered negative controls because there was at least some evidence in the scientific literature linking these cancers to one or more of the contaminants in the drinking water [15, 18, 28–33]. The negative control diseases included for alcohol consumption were mortality due to alcoholism, alcoholic liver disease and chronic liver disease. Several alcohol-related cancers, such as cancers of the oral cavity and pharynx (“oral cancers”), larynx, liver, esophagus, colon and female breast [34] were included in the study but were not considered negative controls because there was at least some evidence in the scientific literature linking these cancers to one or more of the contaminants in the drinking water [15, 18, 30–31, 35–36].

Quantitative bias analyses were conducted to estimate quantitatively, and adjust the HR estimates for, the systematic errors (or biases) due to unmeasured confounding factors and exposure misclassification. The analyses focused on the dichotomous subgroup comparisons between Camp Lejeune and Camp Pendleton, and used Excel spreadsheets included with the textbook, <u>Applying</u> <u>Quantitative Bias Analysis to Epidemiologic Data, Second Edition</u> [37]. A quantitative bias analysis involves choosing a bias model (e.g., exposure misclassification), an analytic technique (e.g., a multidimensional analysis), and values for the parameters of the bias model (e.g., for exposure misclassification, the bias parameters could be the sensitivity and specificity of the exposure classification). The values of the bias parameters are applied to the observed data using bias adjustment equations to calculate what the data would have been if the bias were absent. The quantitative bias analyses of the impacts of unmeasured confounding due to smoking and alcohol consumption used the negative control results to determine the values for the bias parameters of the bias model.

Quantitative bias analyses of exposure misclassification assumed that the misclassification was non-differential and independent because: (1) the base assignments derived from the unit codes for Marines/Navy personnel were completed over ten years prior to cancer data collection, and (2) the base location of employment for civilian workers was recorded in the DMDC database more than thirty years prior to cancer data collection [37].

For Camp Lejeune Marines/Navy personnel, the sources of possible exposure misclassification were due to using unit assignment to a base as a proxy for exposure to the drinking water. First, errors were possible in the historical research conducted by the DMDC and USMC to determine the base where each unit was located. Second, even if the base assignment of the unit was correct, some individuals may not have been exposed to the contaminated drinking water because they were deployed to a different base (e.g., outside the country) or trained at a different base. Third, some individuals stationed at Camp Lejeune may not have been exposed because all their water consumption (including showering and other water uses) occurred off-base (e.g., in off-base housing) or in areas of the base not served by the HP or TT drinking water systems. On the other hand, most of those classified as stationed at Camp Pendleton likely were truly unexposed to the contaminated drinking water.

For Camp Lejeune civilian workers, the main source of exposure misclassification was due to water consumption (including showering and other water uses) occurring mostly or entirely off-base (e.g., at their residences). In addition, the workplaces of some of the Camp Lejeune civilian workers may have been located in areas not served by the contaminated drinking water. All civilian workers at Camp Pendleton were assumed to be truly unexposed to contaminated drinking water during the study period.

To conduct the quantitative bias analyses, it was assumed that the sensitivity of the exposure classification for the Marines/Navy personnel and civilian workers, i.e., the probability that the truly exposed individuals were correctly classified as exposed (i.e., assigned to Camp Lejeune) was near 1.0. The specificity of the exposure classification, i.e., the probability that the truly unexposed individuals were correctly classified as unexposed (i.e., assigned to Camp Pendleton) was assumed to range from 0.81 to 0.91. The chosen values for sensitivity and specificity used in the quantitative bias analysis reflected the assumptions that between 75% and 90% of those stationed or employed at Camp Lejeune were truly exposed, and all (or virtually all) of those stationed or employed at Camp Pendleton were truly unexposed.

Interpretation of study findings was based primarily on the magnitude of the adjusted HR, its precision, and whether a finding was supported by other studies published in the scientific literature of occupational or drinking water exposures to the chemicals found in the drinking water at Camp Lejeune. Because many of the meta-analyses published in the scientific literature for TCE occupational exposures and kidney cancer, NHL and liver cancer observed summary risk ratios between 1.20 and 1.40 [15], the present study emphasized HRs ≥1.20. A HR of 1.20 implies that the cancer occurs 1.2 times more often in the Camp Lejeune cohort compared to the Camp Pendleton cohort.

For Marines/Navy personnel, the interpretation of the findings for rare cancers that primarily occur among older populations, such as male breast cancer, was supplemented by the findings from the Cox regression analyses of the Camp Lejeune and Camp Pendleton full cohort. The analyses of duration stationed or employed at Camp Lejeune provided additional information that was used in the interpretation of the findings. Emphasis was on monotonic trends, i.e., when every change in the adjusted HR with increasing duration is in the same direction (e.g., the HR increases), although the trend could have flat segments but never reverse direction [38].

The 95% confidence interval ratio (CIR), measured by the quotient of the upper to lower limit, was used to indicate the precision (or degree of random variability) of the effect estimates i.e., the SIR, the risk ratio (RR) and the HR estimates [39–40]. The ratio is primarily impacted by the level of the confidence interval (e.g., a 95% CI) and the number of cases of a cancer in the groups being compared. The smaller the number of cases, the wider the confidence interval and therefore the larger the CIR. The study emphasized adjusted HRs ≥1.20 with 95% CIRs ≤3.

Because p-values and statistical significance testing are “commonly misused and misinterpreted” [41], significance testing was not used to interpret findings [38, 42]. Instead, the interpretation of findings was based on: (1) the magnitude of the adjusted HR estimate (i.e., ≥1.20), (2) the precision of the estimate (i.e., the 95% CIR ≤3), (3) the quantitative impacts of unmeasured potential confounders (e.g., smoking and alcohol consumption) and exposure misclassification on the adjusted HR estimate, and (4) supporting information from the scientific literature on the health effects of TCE, PCE, vinyl chloride and benzene [40, 42–43]. Analyses were conducted using SAS 9.4 and STATA 16, and SPSS was used for data management.

This study was approved by the Centers for Disease Control and Prevention Institutional Review Board.

## Results

Demographic information for the civilian workers and the subgroup of Marines/Navy personnel is provided in Tables 1a and 1b. Tables providing demographic information and all statistical results for the Camp Pendleton and Camp Lejeune full cohort of Marines/Navy personnel are included in the Supplemental file 1, Tables S1-2 to S1-4.

The median age of the Camp Lejeune and Camp Pendleton Marines/Navy personnel subgroup at the start of follow-up was 35 years, and the median age at the end of follow-up was 57 years (Table 1a). Most of the Marines/Navy personnel were male (95.6%), White (75.7%), and ranged in rank from E1 to E4 (81.5%). The average length of follow-up was 20 years, and the total number of person-years was approximately 7.04 million (Camp Lejeune: 3.42 million, Camp Pendleton: 3.63 million). The total number of malignancies (including bladder cancer in situ) was 24,227 (Camp Lejeune: 12,083 and Camp Pendleton: 12,144). The total number of individuals with a malignancy or with bladder cancer in situ was 22,536 (Camp Lejeune: 11,207, Camp Pendleton: 11,329). The incidence rates were 354 per 100,000 person-years for Camp Lejeune and 335 per 100,000 person-years for Camp Pendleton.

**Table 1a.** Demographic information for the Marines/Navy personnel subgroup: Marines/Navy personnel at risk during the follow-up period who began active duty and were stationed at Camp Lejeune or Camp Pendleton between 1975 and 1985.

| Factor | Camp Lejeune<br>N=154,821 (48.6%) | Camp Pendleton (ref)<br>N=163,484 (51.4%) | Total<br>N=318,305 |
| --- | --- | --- | --- |
| Male | 146,772 (94.8%) | 157,617 (96.4%) | 304,389 (95.6%) |
| Female | 8,049 (5.2%) | 5,867 (3.6%) | 13,916 (4.4%) |
| White | 113,525 (73.3%) | 127,385 (77.9%) | 240,910 (75.7%) |
| African American | 37,138 (24.0%) | 27,599 (16.9%) | 64,737 (20.3%) |
| Other or unknown race | 4,158 (2.7%) | 8,500 (5.2%) | 12,658 (4.0%) |
| Rank E1 – E4 | 126,471 (81.7%) | 132,874 (81.3%) | 259,345 (81.5%) |
| Rank E5 – E9 | 22,662 (14.6%) | 23,051 (14.1%) | 45,713 (14.4%) |
| WO or CO | 5,688 (3.7%) | 7,559 (4.6%) | 13,247 (4.2%) |
| Not a high school graduate | 19,035 (12.3%) | 26,039 (15.9%) | 45,074 (14.2%) |
| High school graduate | 129,843 (83.9%) | 129,419 (79.2%) | 259,262 (81.5%) |
| College graduate | 5,943 (3.8%) | 8,026 (4.9%) | 13,969 (4.4%) |
| Age at start of follow-up<br>(1/1/1996) |  |  |  |
| Mean (years) | 35.0 | 35.2 | 35.1 |
| Median (years) | 35 | 35 | 35 |
| Age at end of follow-up<br>(12/31/2017 or date of death) |  |  |  |
| Mean | 56.3 | 56.5 | 56.4 |
| Median | 57 | 57 | 57 |
| Age ≥60 years | 35,426 (22.9%) | 39,734 (24.3%) | 75,160 (23.6%) |
| Age >69 years | 292 (0.2%) | 277 (0.2%) | 569 (0.2%) |
| Died during 1/2/1996 –<br>12/31/2017 | 13,632 (8.8%) | 14,904 (9.1%) | 28,536 (9.0%) |
| Length of follow-up (years) |  |  |  |
| Mean (years) | 20.3 | 20.3 | 20.3 |
| Median (years) | 21 | 21 | 21 |
| Total person-years of follow-up | 3,417,738 (48.5%) | 3,626,570 (51.5%) | 7,044,308 |

| Quarters in the DMDC data, 1975-1985* | Camp Lejeune | Camp Pendleton |  |
| --- | --- | --- | --- |
| Mean | 7.7 | 7.2 |  |
| Median | 7.0 | 6.0 |  |
| Minimum | 1 | 1 |  |
| Maximum | 41 | 42 |  |
| Interquartile range (25<sup>th</sup>-75<sup>th</sup> percentiles) | 8 (3 – 11) | 8 (3 – 11) |  |
| Cancers | Camp Lejeune | Camp Pendleton | Total |
| Total number of malignancies (including bladder cancer in situ) | 12,083 | 12,144 | 24,227 |
| Total number of individuals with any malignancy or bladder cancer in situ | 11,207 | 11,329 | 22,536 |

Abbreviations: E1 – E4: private to corporal;
E5 – E9: sergeant to sergeant major;
WO: warrant officer;CO: commissioned officer
The table does not include aggregate cancer data obtained from the West Virginia and Kansas cancer registries.
\* Number of quarters at either Camp Lejeune or Camp Pendleton during 1975-1985. Some members of the Camp Lejeune cohort, who were stationed at least one quarter at Camp Lejeune during 1975-1985, were also stationed at Camp Pendleton during 1975-1985. So, the statistics for the Camp Lejeune cohort include quarters at Camp Pendleton during 1975-1985. The Camp Pendleton cohort members were not stationed at Camp Lejeune during 1975-1985.

**Table 1b.** Demographic information for civilian workers. Civilian workers at risk during the follow-up period who were employed at Camp Lejeune or Camp Pendleton between October 1972 and December 1985

| Factor | Camp Lejeune<br>N = 6,494 (52.8%) | Camp Pendleton (ref)<br>N = 5,797 (47.2%) | Total<br>N=12,291 |
| --- | --- | --- | --- |
| Male | 3,026 (46.6%) | 2,992 (51.6%) | 6,018 (49.0%) |
| Female | 3,468 (53.4%) | 2,805 (48.4%) | 6,273 (51.0%) |
| White | 4,998 (77.0%) | 4,483 (77.3%) | 9,481 (77.1%) |
| African American | 1,178 (18.1%) | 461 (8.0%) | 1,639 (13.3%) |
| Other or unknown race | 318 (4.9%) | 853 (14.7%) | 1,171 (9.5%) |
| Blue collar | 2,251 (34.7%) | 2,260 (39.0%) | 4,511 (36.7%) |
| White collar | 4,243 (65.3%) | 3,537 (61.0%) | 7,780 (63.3%) |
| Not a high school graduate | 700 (10.8%) | 483 (8.3%) | 1,183 (9.6%) |
| High school graduate | 4,746 (73.1%) | 4,887 (84.3%) | 9,633 (78.4%) |
| College graduate | 1,048 (16.1%) | 427 (7.4%) | 1,475 (12.0%) |
| Age at start of follow-up<br>(1/1/1996) |  |  |  |
| Mean (years) | 52.9 | 55.1 | 53.0 |
| Median (years) | 50 | 53 | 51 |
| Age at end of follow-up<br>(12/31/2017 or date of death) |  |  |  |
| Mean | 72.1 | 73.4 | 72.7 |
| Median | 71 | 73 | 71 |
| Age >65 years | 4,728 (72.8%) | 4,288 (74.0%) | 9,016 (73.4%) |
| Age >70 years | 3,288 (50.6%) | 3,270 (56.4%) | 6,558 (53.4%) |
| Age >75 years | 2,228 (34.3%) | 2,415 (41.7%) | 4,643 (37.8%) |
| Died during 1/2/1996 –<br>12/31/2017 | 2,251 (34.7%) | 2,433 (42.0%) | 4,684 (38.1%) |
| Length of follow-up (years) |  |  |  |
| Mean (years) | 17.7 | 17.0 | 17.4 |
| Median (years) | 21 | 21 | 21 |
| Total person-years of follow-up | 120,148 (53.8%) | 103,234 (46.2%) | 223,382 |

| Quarters in the DMDC data,<br>10/1972-12/1985* | Camp Lejeune | Camp Pendleton |  |
| --- | --- | --- | --- |
| Mean | 19.5 | 17.6 |  |
| Median | 12.0 | 10.0 |  |
| Minimum | 1 | 1 |  |
| Maximum | 53 | 53 |  |
| Interquartile range (25<sup>th</sup>-75<sup>th</sup><br>percentiles) | 32 (3 – 35) | 24 (4 – 28) |  |
| Cancers | Camp Lejeune | Camp Pendleton (ref) | Total |
| Total number of malignancies<br>(including bladder cancer in<br>situ) | 1,563 | 1,416 | 2,979 |
| Total number of individuals<br>with any malignancy or with<br>bladder cancer in situ | 1,359 | 1,240 | 2,599 |

The table does not include aggregate cancer data obtained from the West Virginia and Kansas cancer registries.
\* Number of quarters employed at either Camp Lejeune or Camp Pendleton during 10/72- 12/1985. Some members of the Camp Lejeune cohort, who were employed at least one quarter at Camp Lejeune during 10/72-12/1985, were also employed at Camp Pendleton during 10/72- 12/1985. So, the statistics for the Camp Lejeune cohort include quarters at Camp Pendleton during 10/72-12/1985. The Camp Pendleton cohort members were not employed at Camp Lejeune during 10/72-12/1985.

For civilian workers (Table 1b), the percentages of women in the workforce at Camp Lejeune and Camp Pendleton were 53.4% and 48.4%, respectively. Most of the workforce at both bases were white (77%). A much higher percentage of the Camp Lejeune workforce was African American (18.1%) compared to Camp Pendleton (8.0%). A higher percentage of workers at Camp Lejeune graduated from college (16.1%) compared to Camp Pendleton (7.4%). Over half of the workers in the study were above 70 years of age at the end of follow-up. The average length of follow-up was slightly over 17 years, and the total amount of person-years was 223,382. The total number of malignant cancers (including bladder cancer in situ) was 2,979 (Camp Lejeune: 1,563, Camp Pendleton: 1,416). The total number of individuals with a malignancy or with bladder cancer in situ was 2,599 (Camp Lejeune: 1,359, Camp Pendleton: 1,240). The incidence rates were 1,301 per 100,000 person-years for Camp Lejeune and 1,372 per 100,000 person-years for Camp Pendleton.

The results of the SIR and Poisson regression analyses for the Camp Lejeune and Camp Pendleton Marines/Navy personnel subgroup are shown in Table 2. The SIRs for many of the cancers evaluated were less than 1.00, consistent with a “healthy veteran effect.” [21–23]. The healthy veteran effect is due to several factors including the initial physical screening for healthy recruits, physical fitness standards during military service, and access to quality health care during and after service. The healthy veteran effect may have been especially strong in the subgroup because over three-quarters of the members of the subgroup were less than 60 years of age at the end of follow-up (Table 1a). However, SIRs were above 1.00 at both Camp Lejeune and Camp Pendleton for melanoma, oral cancers and cancers of the brain and central nervous system, and female breast (Table 2).

**Table 2.**
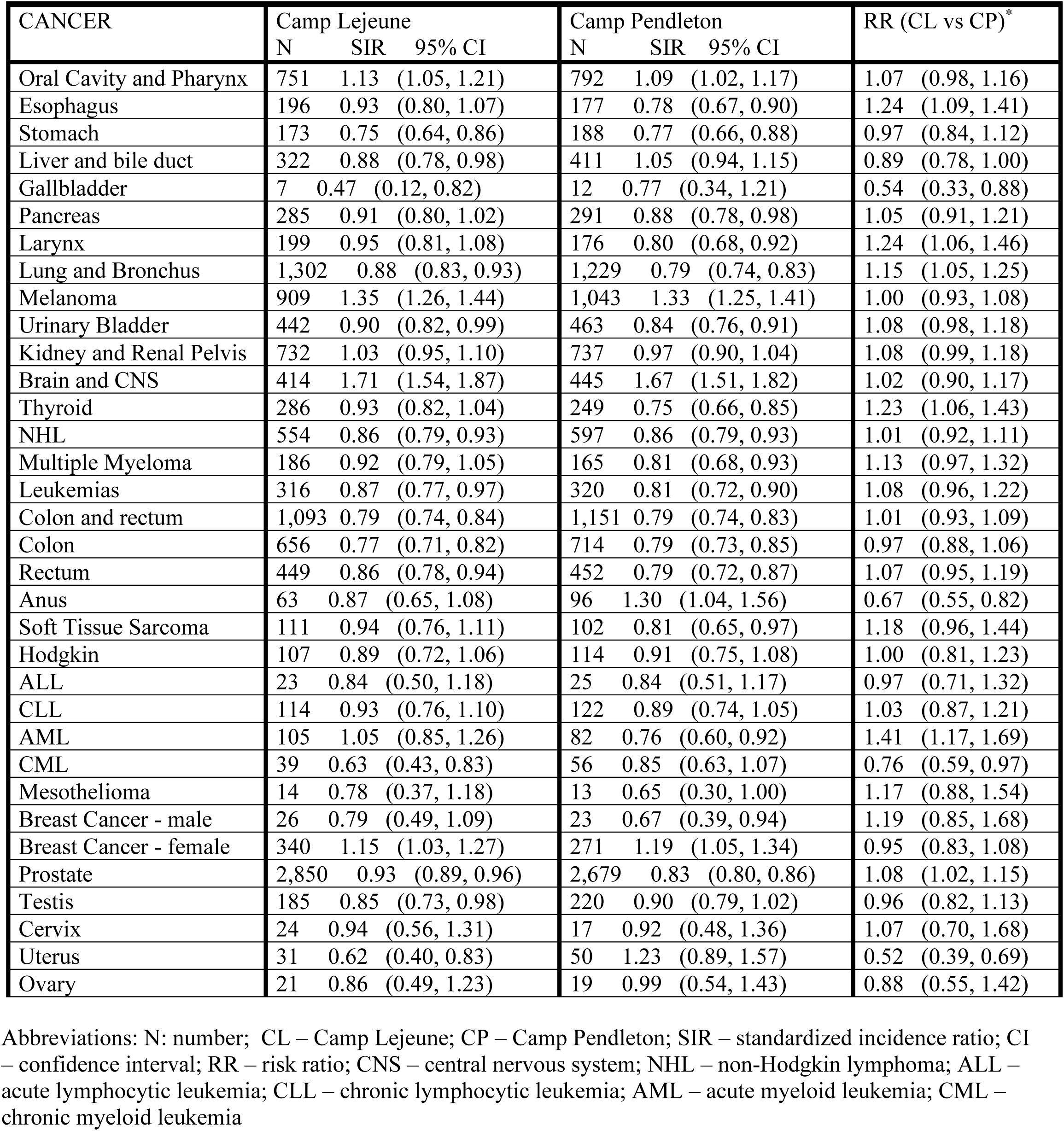
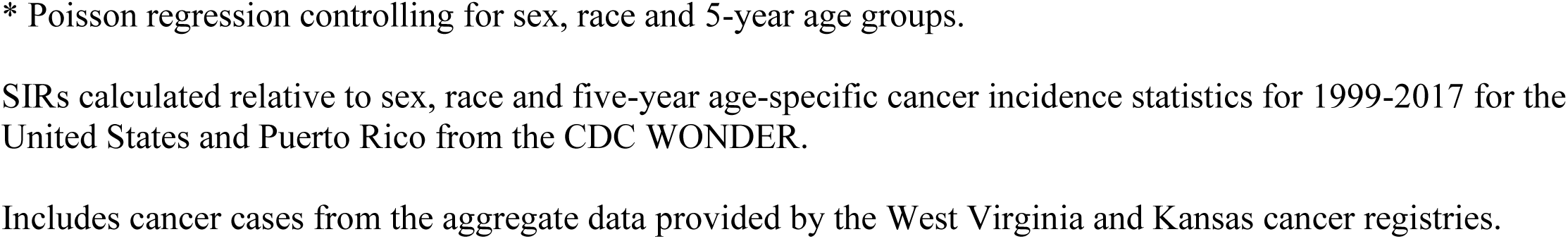
Standardized incidence rates and Poisson regression results: Marines/Navy personnel subgroup.

| CANCER | Camp Lejeune |  |  | Camp Pendleton |  |  | RR (CL vs CP)* |
| --- | --- | --- | --- | --- | --- | --- | --- |
|  | N | SIR | 95% CI | N | SIR | 95% CI |  |
| Oral Cavity and Pharynx | 751 | 1.13 | (1.05, 1.21) | 792 | 1.09 | (1.02, 1.17) | 1.07 (0.98, 1.16) |
| Esophagus | 196 | 0.93 | (0.80, 1.07) | 177 | 0.78 | (0.67, 0.90) | 1.24 (1.09, 1.41) |
| Stomach | 173 | 0.75 | (0.64, 0.86) | 188 | 0.77 | (0.66, 0.88) | 0.97 (0.84, 1.12) |
| Liver and bile duct | 322 | 0.88 | (0.78, 0.98) | 411 | 1.05 | (0.94, 1.15) | 0.89 (0.78, 1.00) |
| Gallbladder | 7 | 0.47 | (0.12, 0.82) | 12 | 0.77 | (0.34, 1.21) | 0.54 (0.33, 0.88) |
| Pancreas | 285 | 0.91 | (0.80, 1.02) | 291 | 0.88 | (0.78, 0.98) | 1.05 (0.91, 1.21) |
| Larynx | 199 | 0.95 | (0.81, 1.08) | 176 | 0.80 | (0.68, 0.92) | 1.24 (1.06, 1.46) |
| Lung and Bronchus | 1,302 | 0.88 | (0.83, 0.93) | 1,229 | 0.79 | (0.74, 0.83) | 1.15 (1.05, 1.25) |
| Melanoma | 909 | 1.35 | (1.26, 1.44) | 1,043 | 1.33 | (1.25, 1.41) | 1.00 (0.93, 1.08) |
| Urinary Bladder | 442 | 0.90 | (0.82, 0.99) | 463 | 0.84 | (0.76, 0.91) | 1.08 (0.98, 1.18) |
| Kidney and Renal Pelvis | 732 | 1.03 | (0.95, 1.10) | 737 | 0.97 | (0.90, 1.04) | 1.08 (0.99, 1.18) |
| Brain and CNS | 414 | 1.71 | (1.54, 1.87) | 445 | 1.67 | (1.51, 1.82) | 1.02 (0.90, 1.17) |
| Thyroid | 286 | 0.93 | (0.82, 1.04) | 249 | 0.75 | (0.66, 0.85) | 1.23 (1.06, 1.43) |
| NHL | 554 | 0.86 | (0.79, 0.93) | 597 | 0.86 | (0.79, 0.93) | 1.01 (0.92, 1.11) |
| Multiple Myeloma | 186 | 0.92 | (0.79, 1.05) | 165 | 0.81 | (0.68, 0.93) | 1.13 (0.97, 1.32) |
| Leukemias | 316 | 0.87 | (0.77, 0.97) | 320 | 0.81 | (0.72, 0.90) | 1.08 (0.96, 1.22) |
| Colon and rectum | 1,093 | 0.79 | (0.74, 0.84) | 1,151 | 0.79 | (0.74, 0.83) | 1.01 (0.93, 1.09) |
| Colon | 656 | 0.77 | (0.71, 0.82) | 714 | 0.79 | (0.73, 0.85) | 0.97 (0.88, 1.06) |
| Rectum | 449 | 0.86 | (0.78, 0.94) | 452 | 0.79 | (0.72, 0.87) | 1.07 (0.95, 1.19) |
| Anus | 63 | 0.87 | (0.65, 1.08) | 96 | 1.30 | (1.04, 1.56) | 0.67 (0.55, 0.82) |
| Soft Tissue Sarcoma | 111 | 0.94 | (0.76, 1.11) | 102 | 0.81 | (0.65, 0.97) | 1.18 (0.96, 1.44) |
| Hodgkin | 107 | 0.89 | (0.72, 1.06) | 114 | 0.91 | (0.75, 1.08) | 1.00 (0.81, 1.23) |
| ALL | 23 | 0.84 | (0.50, 1.18) | 25 | 0.84 | (0.51, 1.17) | 0.97 (0.71, 1.32) |
| CLL | 114 | 0.93 | (0.76, 1.10) | 122 | 0.89 | (0.74, 1.05) | 1.03 (0.87, 1.21) |
| AML | 105 | 1.05 | (0.85, 1.26) | 82 | 0.76 | (0.60, 0.92) | 1.41 (1.17, 1.69) |
| CML | 39 | 0.63 | (0.43, 0.83) | 56 | 0.85 | (0.63, 1.07) | 0.76 (0.59, 0.97) |
| Mesothelioma | 14 | 0.78 | (0.37, 1.18) | 13 | 0.65 | (0.30, 1.00) | 1.17 (0.88, 1.54) |
| Breast Cancer - male | 26 | 0.79 | (0.49, 1.09) | 23 | 0.67 | (0.39, 0.94) | 1.19 (0.85, 1.68) |
| Breast Cancer - female | 340 | 1.15 | (1.03, 1.27) | 271 | 1.19 | (1.05, 1.34) | 0.95 (0.83, 1.08) |
| Prostate | 2,850 | 0.93 | (0.89, 0.96) | 2,679 | 0.83 | (0.80, 0.86) | 1.08 (1.02, 1.15) |
| Testis | 185 | 0.85 | (0.73, 0.98) | 220 | 0.90 | (0.79, 1.02) | 0.96 (0.82, 1.13) |
| Cervix | 24 | 0.94 | (0.56, 1.31) | 17 | 0.92 | (0.48, 1.36) | 1.07 (0.70, 1.68) |
| Uterus | 31 | 0.62 | (0.40, 0.83) | 50 | 1.23 | (0.89, 1.57) | 0.52 (0.39, 0.69) |
| Ovary | 21 | 0.86 | (0.49, 1.23) | 19 | 0.99 | (0.54, 1.43) | 0.88 (0.55, 1.42) |

Abbreviations: N: number; CL – Camp Lejeune; CP – Camp Pendleton; SIR – standardized incidence ratio; CI – confidence interval; RR – risk ratio; CNS – central nervous system; NHL – non-Hodgkin lymphoma; ALL – acute lymphocytic leukemia; CLL – chronic lymphocytic leukemia; AML – acute myeloid leukemia; CML – chronic myeloid leukemia
\* Poisson regression controlling for sex, race and 5-year age groups.
SIRs calculated relative to sex, race and five-year age-specific cancer incidence statistics for 1999-2017 for the United States and Puerto Rico from the CDC WONDER.
Includes cancer cases from the aggregate data provided by the West Virginia and Kansas cancer registries.

The Poisson regression analyses comparing the Camp Lejeune and Camp Pendleton Marines/Navy personnel subgroup observed RRs ≥1.20 (i.e., an increase in risk of ≥20%) with 95% CIRs ≤3 for acute myeloid leukemia (AML) (RR=1.41, 95% CI: 1.17, 1.69), and cancers of the esophagus (RR=1.24, 95% CI: 1.09, 1.41), larynx (RR=1.24, 95% CI: 1.06, 1.46) and thyroid (RR=1.23, 95% CI: 1.06, 1.43) (Table 2). In the Poisson regression analyses comparing the Camp Lejeune and Camp Pendleton full cohort, most of the results were similar to the subgroup results. However, for male breast cancer, the number of cases in the full cohort was nearly double that in the subgroup and the RR was 1.39 (95% CI: 1.05, 1.85) (Supplemental file 1, Table S1-3). For comparison, the RR for male breast cancer was 1.19 (95% CI: 0.85, 1.68) in the subgroup (Table 2).

The results of the SIR and Poisson regression analyses for the civilian workers are shown in Table 3. Compared to Camp Pendleton, civilian workers at Camp Lejeune had RRs ≥1.20 with 95% CIR ≤3 for NHL (RR=1.24, 95% CI: 0.91, 1.68), female breast cancer (RR=1.23, 95% CI: 0.96, 1.58), oral cancers (RR=1.65, 95% CI: 1.00, 2.72) and AML (RR=1.30, 95% CI: 0.77, 2.19). Thyroid cancer and male breast cancer had RRs ≥1.20 but with 95% CIRs >3.

**Table 3.**
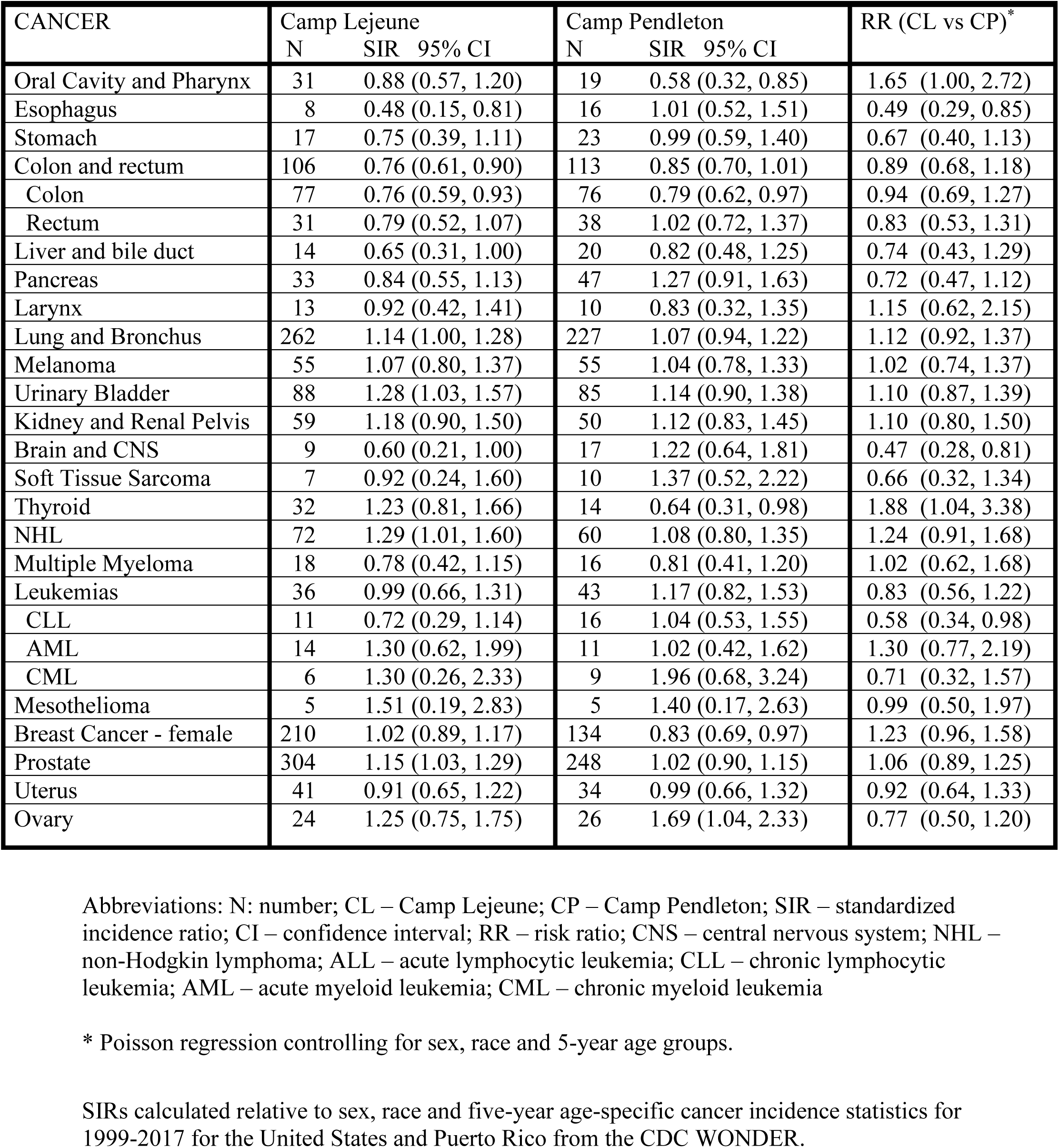
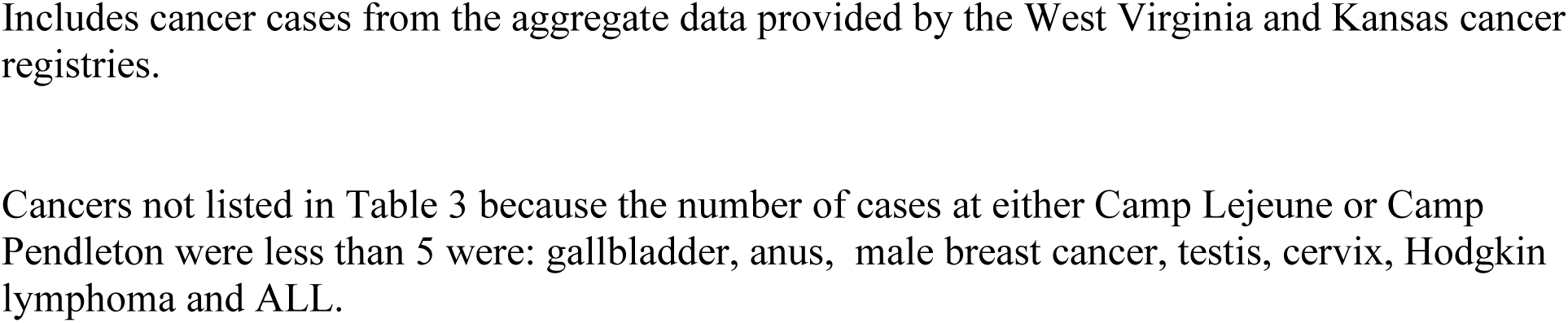
Standardized incidence rates (SIR) and Poisson regression results: Civilian workers.

| CANCER | Camp Lejeune |  |  | Camp Pendleton |  |  | RR (CL vs CP)* |
| --- | --- | --- | --- | --- | --- | --- | --- |
|  | N | SIR | 95% CI | N | SIR | 95% CI |  |
| Oral Cavity and Pharynx | 31 | 0.88 | (0.57, 1.20) | 19 | 0.58 | (0.32, 0.85) | 1.65 (1.00, 2.72) |
| Esophagus | 8 | 0.48 | (0.15, 0.81) | 16 | 1.01 | (0.52, 1.51) | 0.49 (0.29, 0.85) |
| Stomach | 17 | 0.75 | (0.39, 1.11) | 23 | 0.99 | (0.59, 1.40) | 0.67 (0.40, 1.13) |
| Colon and rectum | 106 | 0.76 | (0.61, 0.90) | 113 | 0.85 | (0.70, 1.01) | 0.89 (0.68, 1.18) |
| Colon | 77 | 0.76 | (0.59, 0.93) | 76 | 0.79 | (0.62, 0.97) | 0.94 (0.69, 1.27) |
| Rectum | 31 | 0.79 | (0.52, 1.07) | 38 | 1.02 | (0.72, 1.37) | 0.83 (0.53, 1.31) |
| Liver and bile duct | 14 | 0.65 | (0.31, 1.00) | 20 | 0.82 | (0.48, 1.25) | 0.74 (0.43, 1.29) |
| Pancreas | 33 | 0.84 | (0.55, 1.13) | 47 | 1.27 | (0.91, 1.63) | 0.72 (0.47, 1.12) |
| Larynx | 13 | 0.92 | (0.42, 1.41) | 10 | 0.83 | (0.32, 1.35) | 1.15 (0.62, 2.15) |
| Lung and Bronchus | 262 | 1.14 | (1.00, 1.28) | 227 | 1.07 | (0.94, 1.22) | 1.12 (0.92, 1.37) |
| Melanoma | 55 | 1.07 | (0.80, 1.37) | 55 | 1.04 | (0.78, 1.33) | 1.02 (0.74, 1.37) |
| Urinary Bladder | 88 | 1.28 | (1.03, 1.57) | 85 | 1.14 | (0.90, 1.38) | 1.10 (0.87, 1.39) |
| Kidney and Renal Pelvis | 59 | 1.18 | (0.90, 1.50) | 50 | 1.12 | (0.83, 1.45) | 1.10 (0.80, 1.50) |
| Brain and CNS | 9 | 0.60 | (0.21, 1.00) | 17 | 1.22 | (0.64, 1.81) | 0.47 (0.28, 0.81) |
| Soft Tissue Sarcoma | 7 | 0.92 | (0.24, 1.60) | 10 | 1.37 | (0.52, 2.22) | 0.66 (0.32, 1.34) |
| Thyroid | 32 | 1.23 | (0.81, 1.66) | 14 | 0.64 | (0.31, 0.98) | 1.88 (1.04, 3.38) |
| NHL | 72 | 1.29 | (1.01, 1.60) | 60 | 1.08 | (0.80, 1.35) | 1.24 (0.91, 1.68) |
| Multiple Myeloma | 18 | 0.78 | (0.42, 1.15) | 16 | 0.81 | (0.41, 1.20) | 1.02 (0.62, 1.68) |
| Leukemias | 36 | 0.99 | (0.66, 1.31) | 43 | 1.17 | (0.82, 1.53) | 0.83 (0.56, 1.22) |
| CLL | 11 | 0.72 | (0.29, 1.14) | 16 | 1.04 | (0.53, 1.55) | 0.58 (0.34, 0.98) |
| AML | 14 | 1.30 | (0.62, 1.99) | 11 | 1.02 | (0.42, 1.62) | 1.30 (0.77, 2.19) |
| CML | 6 | 1.30 | (0.26, 2.33) | 9 | 1.96 | (0.68, 3.24) | 0.71 (0.32, 1.57) |
| Mesothelioma | 5 | 1.51 | (0.19, 2.83) | 5 | 1.40 | (0.17, 2.63) | 0.99 (0.50, 1.97) |
| Breast Cancer - female | 210 | 1.02 | (0.89, 1.17) | 134 | 0.83 | (0.69, 0.97) | 1.23 (0.96, 1.58) |
| Prostate | 304 | 1.15 | (1.03, 1.29) | 248 | 1.02 | (0.90, 1.15) | 1.06 (0.89, 1.25) |
| Uterus | 41 | 0.91 | (0.65, 1.22) | 34 | 0.99 | (0.66, 1.32) | 0.92 (0.64, 1.33) |
| Ovary | 24 | 1.25 | (0.75, 1.75) | 26 | 1.69 | (1.04, 2.33) | 0.77 (0.50, 1.20) |

Abbreviations: N: number; CL – Camp Lejeune; CP – Camp Pendleton; SIR – standardized incidence ratio; CI – confidence interval; RR – risk ratio; CNS – central nervous system; NHL – non-Hodgkin lymphoma; ALL – acute lymphocytic leukemia; CLL – chronic lymphocytic leukemia; AML – acute myeloid leukemia; CML – chronic myeloid leukemia
\* Poisson regression controlling for sex, race and 5-year age groups.
SIRs calculated relative to sex, race and five-year age-specific cancer incidence statistics for 1999-2017 for the United States and Puerto Rico from the CDC WONDER.
Includes cancer cases from the aggregate data provided by the West Virginia and Kansas cancer registries.
Cancers not listed in Table 3 because the number of cases at either Camp Lejeune or Camp Pendleton were less than 5 were: gallbladder, anus, male breast cancer, testis, cervix, Hodgkin lymphoma and ALL.

In the analyses using Cox regression methods, the unadjusted and adjusted HRs comparing the Camp Lejeune and Camp Pendleton Marines/Navy personnel subgroup are shown in Table 4. (The Cox regression results for the comparisons between the Camp Lejeune and Camp Pendleton full cohort are provided in Supplemental file 1, Table S1-4). The Cox regressions included age as the time variable, base where the individual’s unit was stationed, sex, race, rank, and education level during the study period. Adjusted HRs ≥1.20 with 95% CIRs ≤3 were observed for AML (HR=1.38, 95% CI: 1.03, 1.85), all myeloid cancers including polycythemia vera (HR=1.24, 95% CI: 1.03, 1.49) and cancers of the esophagus (HR=1.27, 95% CI: 1.03, 1.56), larynx (HR=1.21, 95% CI: 0.98, 1.50), soft tissue (HR=1.21, 95% CI: 0.92, 1.59) and thyroid (HR=1.22, 95% CI: 1.03, 1.45). Adjusted HRs ≥1.20 with 95% CIRs ≤3 were also observed for lung cancer histological subtypes, non-small cell carcinoma (HR=1.23, 95% CI: 0.97, 1.56), large cell lung cancer (HR=1.38, 95% CI: 0.84, 2.28) and adenocarcinoma (HR=1.25, 95% CI: 1.10, 1.41). In addition, adjusted HRs ≥1.20 with 95% CIRs ≤3 were observed for myelodysplastic and myeloproliferative syndromes (HR = 1.68, 95% CI: 1.07, 2.62), polycythemia vera (HR=1.41, 95% CI: 0.94, 2.11), marginal zone B-cell (MZBCL) lymphoma (HR=1.45, 95% CI: 0.92, 2.28), and squamous cell esophageal cancer (HR=1.47, 95% CI: 0.96, 2.25).

**Table 4.**
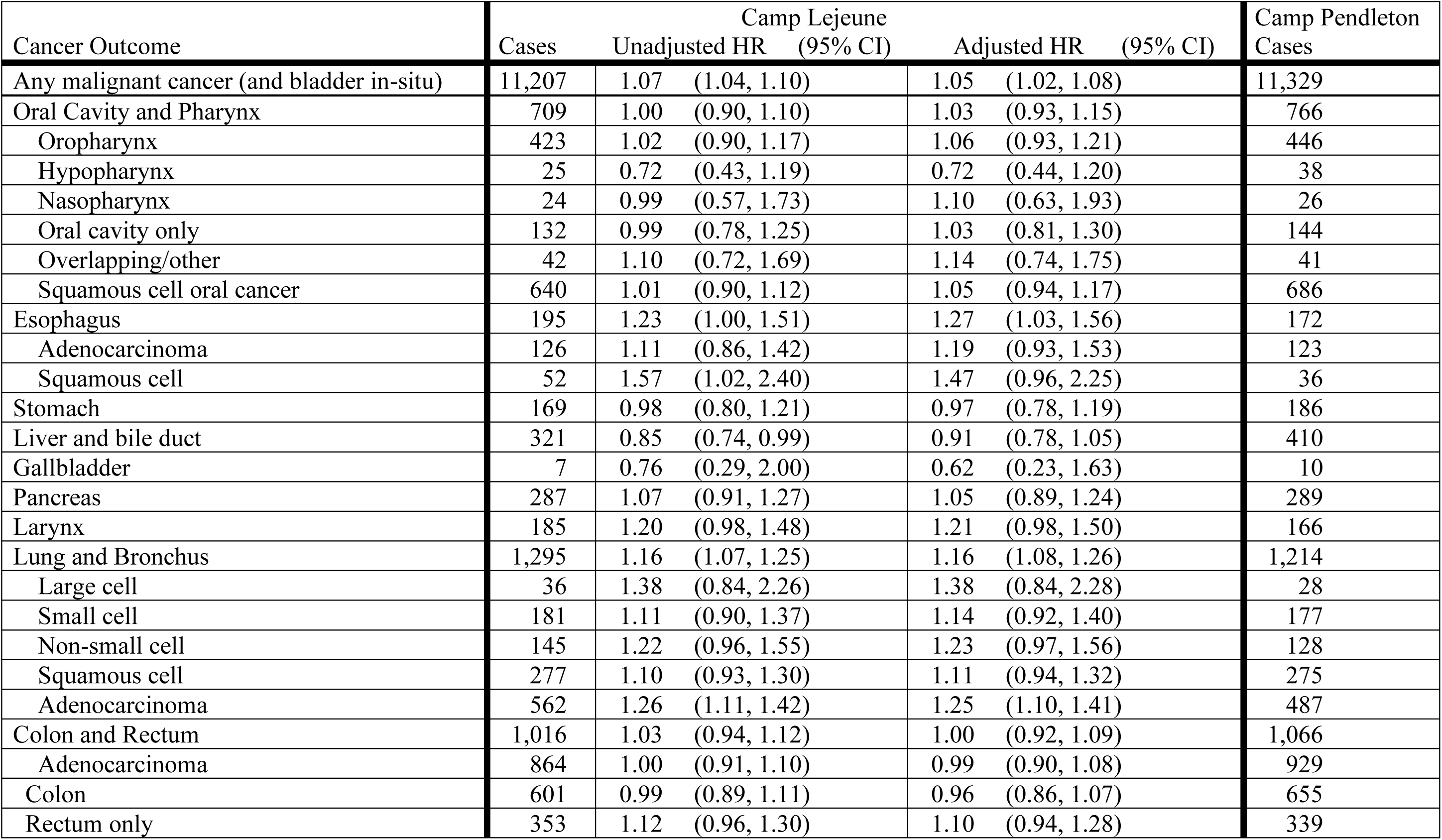
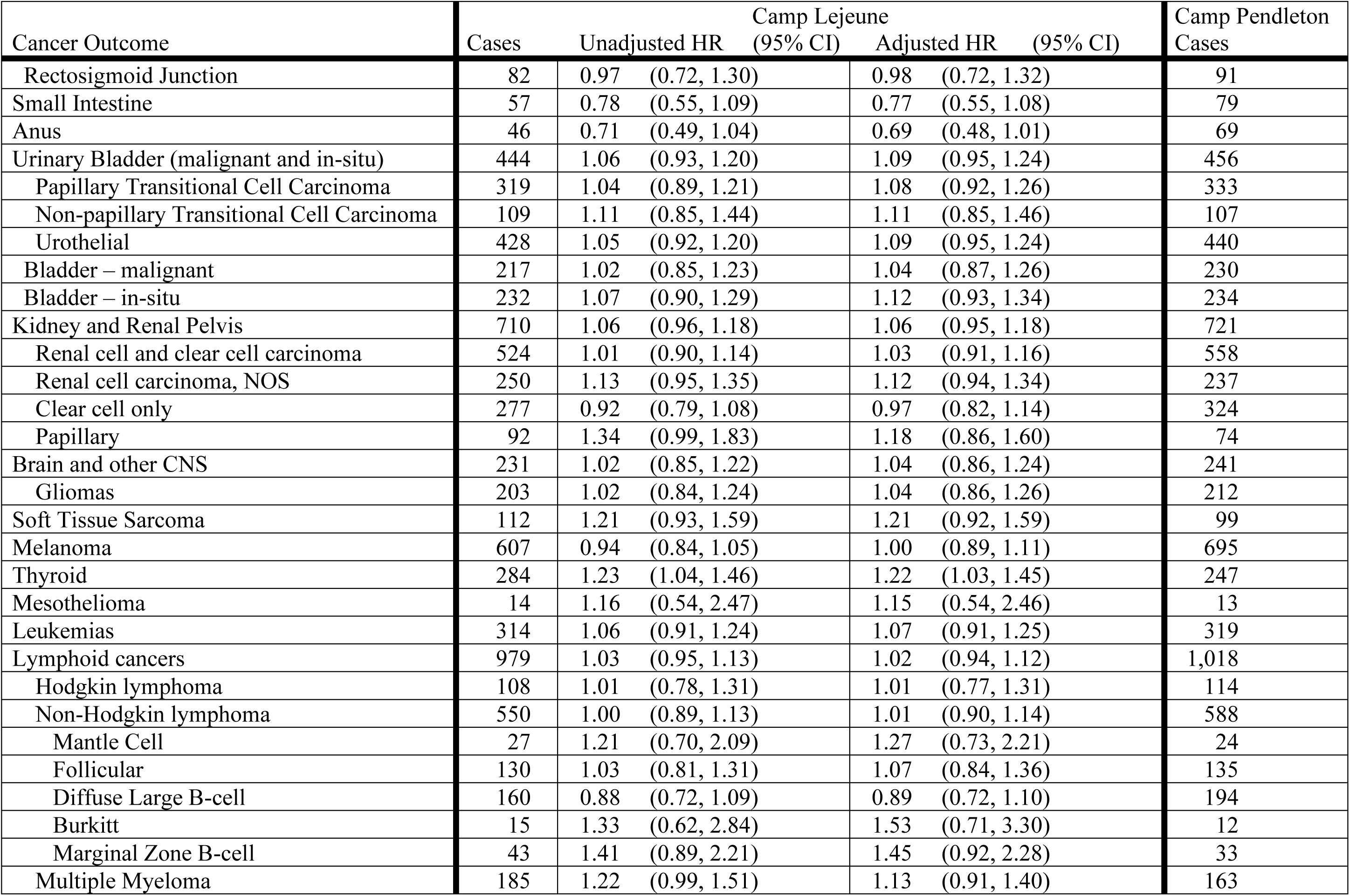
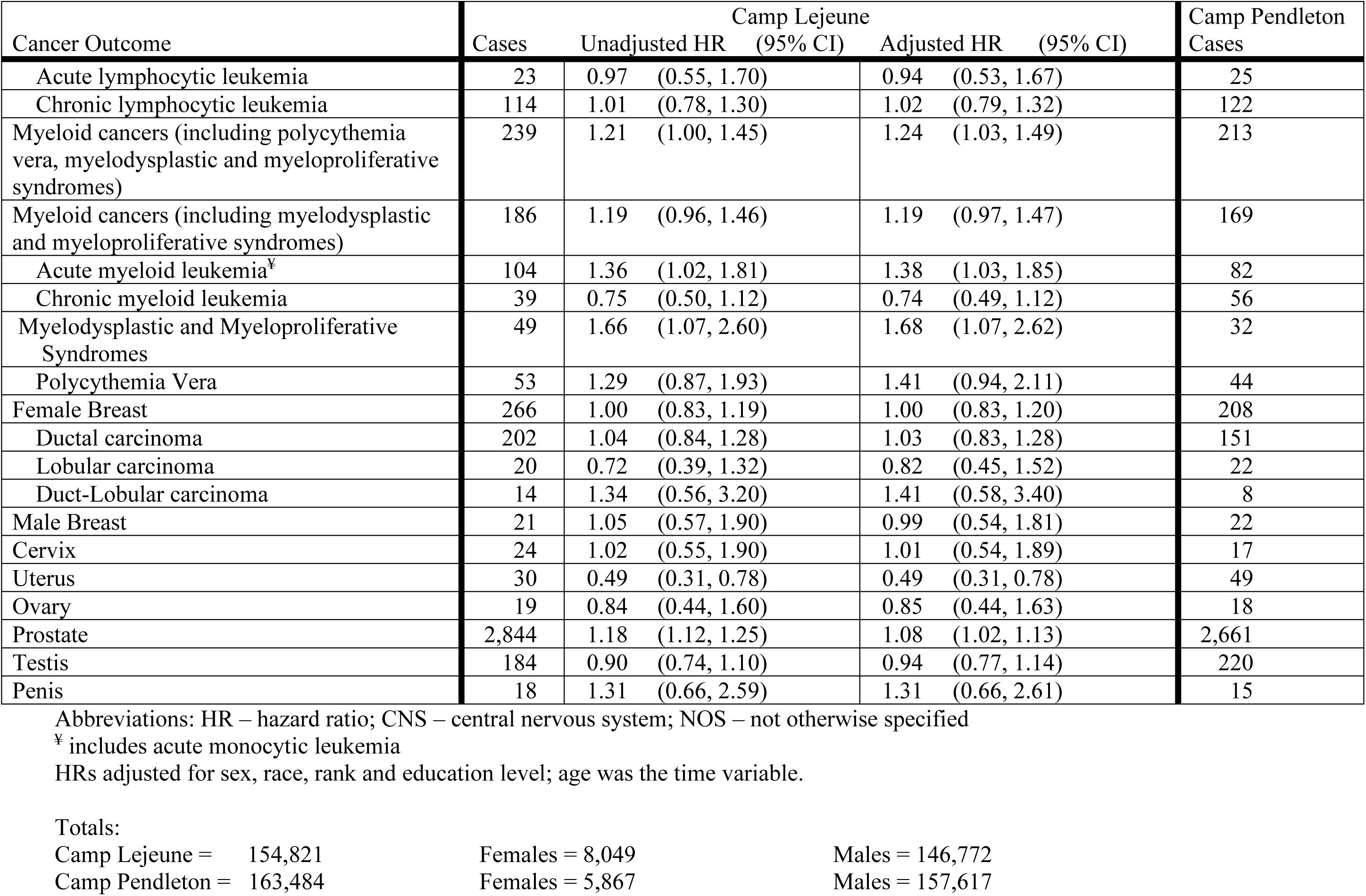
Comparison of base location at Camp Lejeune vs Camp Pendleton: Marines/Navy personnel subgroup.

Most of the Cox regression adjusted results for the Camp Lejeune and Camp Pendleton Marines/Navy personnel full cohort appeared similar to the subgroup results (Supplemental file 1, Table S1-4), except for male breast cancer. In the subgroup analysis, the HR for male breast cancer was 0.99 with a 95% CIR >3, whereas the HR in the full cohort was 1.24 with a 95% CIR ≤3.

For civilian workers, the Cox regression analysis comparing Camp Lejeune to Camp Pendleton is presented in Table 5. Adjusted hazard ratios (HRs) ≥1.20 with 95% CIRs ≤3 were observed for all myeloid cancers including polycythemia vera (HR=1.40, 95% CI: 0.83, 2.36) and squamous cell lung cancer (1.63, 95% CI: 1.10, 2.41). NHL had an adjusted HR of 1.19 (95% CI: 0.83, 1.71) and female breast cancer had an adjusted HR of 1.19 (95% CI: 0.95, 1.49). The female breast cancer histological subtype ductal carcinoma had an adjusted HR of 1.32 (95% CI:1.02, 1.71). Several cancers and histological subtypes had adjusted HRs ≥1.20 but with 95% CIRs >3 including male breast cancer, oral cancers, thyroid cancer, acute myeloid leukemia, myelodysplastic and myeloproliferative syndromes, follicular and diffuse large B-cell lymphomas, and non-papillary transitional cell bladder carcinoma.

**Table 5.**
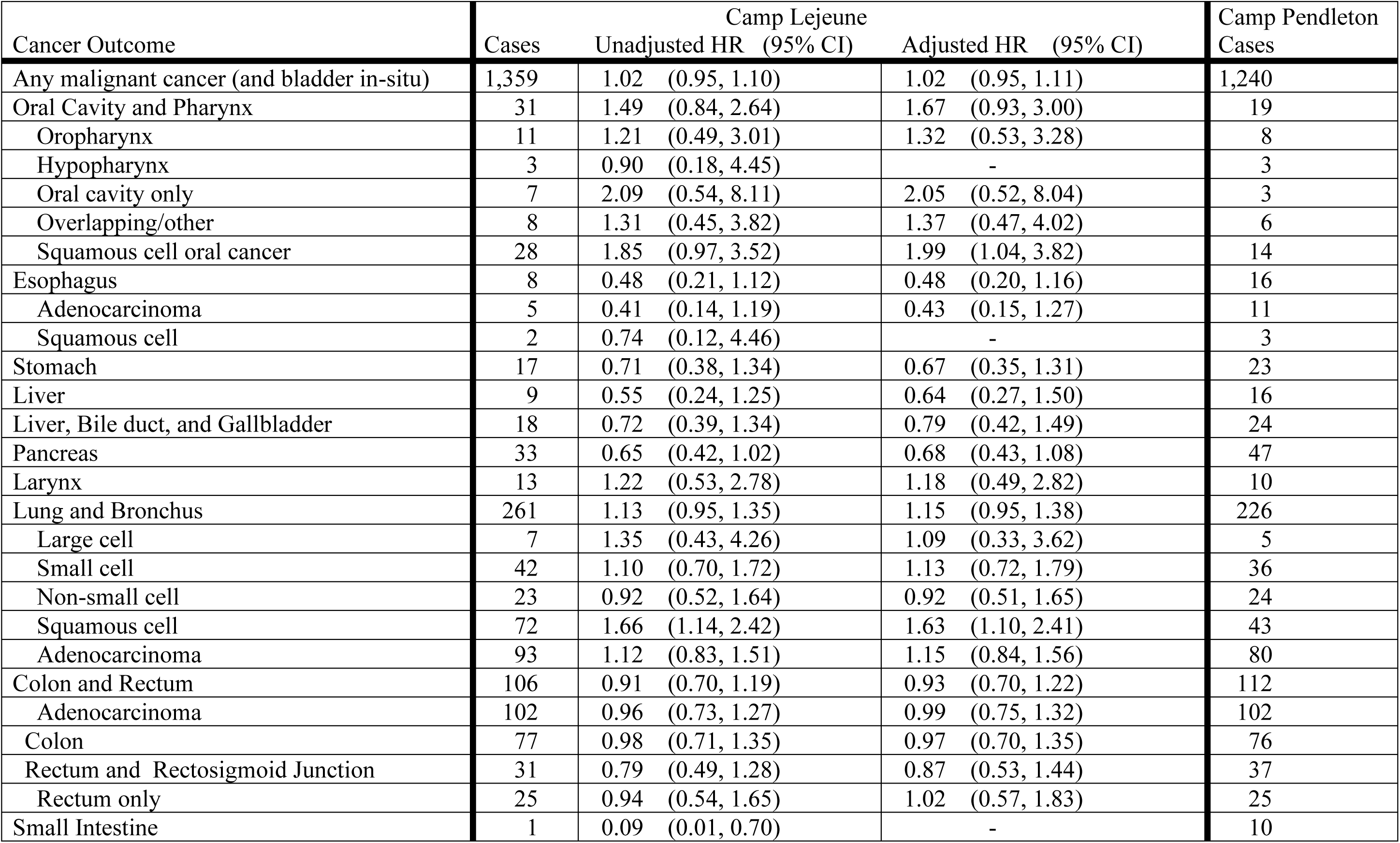
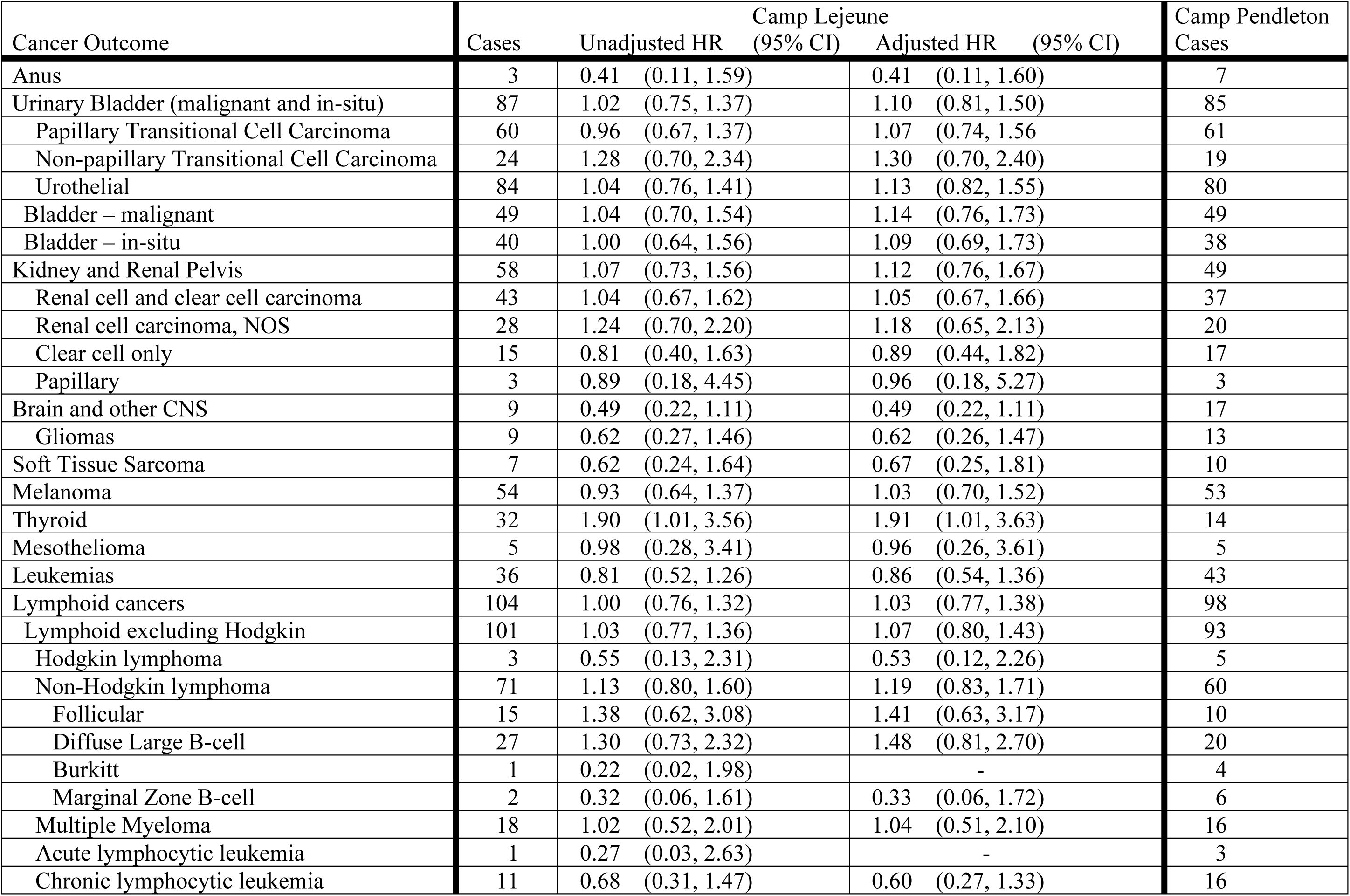
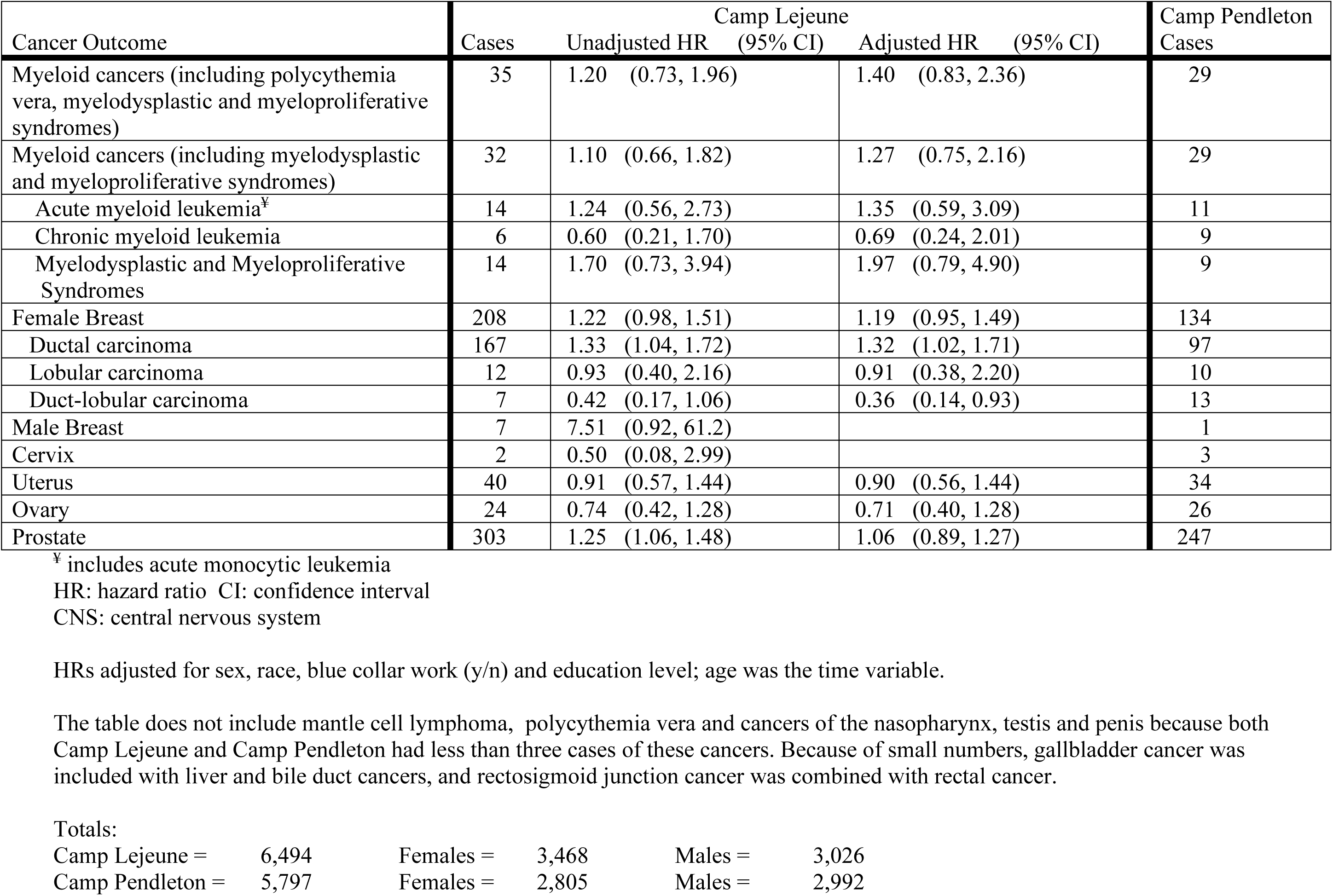
Comparison of Camp Lejeune versus Camp Pendleton civilian workers.

The Marines/Navy subgroup analysis of duration stationed at Camp Lejeune between 1975 and 1985 as a categorical variable is presented in Table 6. The reference group consisted of those Marines/Navy personnel stationed at Camp Pendleton and not Camp Lejeune between 1975 and 1985. The levels of duration were approximately quartiles of the data after removal of the reference group. Since the DMDC data was quarterly, the levels of the categorical variable consisted of the number of quarters the individual was stationed at Camp Lejeune: “low” duration (1 – 2 quarters), “medium” duration (>2 – 6 quarters), “medium/high” duration (>6 – 10 quarters) and “high” duration (>10 quarters). A monotonic trend for thyroid cancer was observed, with the adjusted HR at the highest duration level of 1.32 (95% CI: 1.00, 1.75). No other monotonic trends were identified, and further results are reported in Table 6.

**Table 6.**
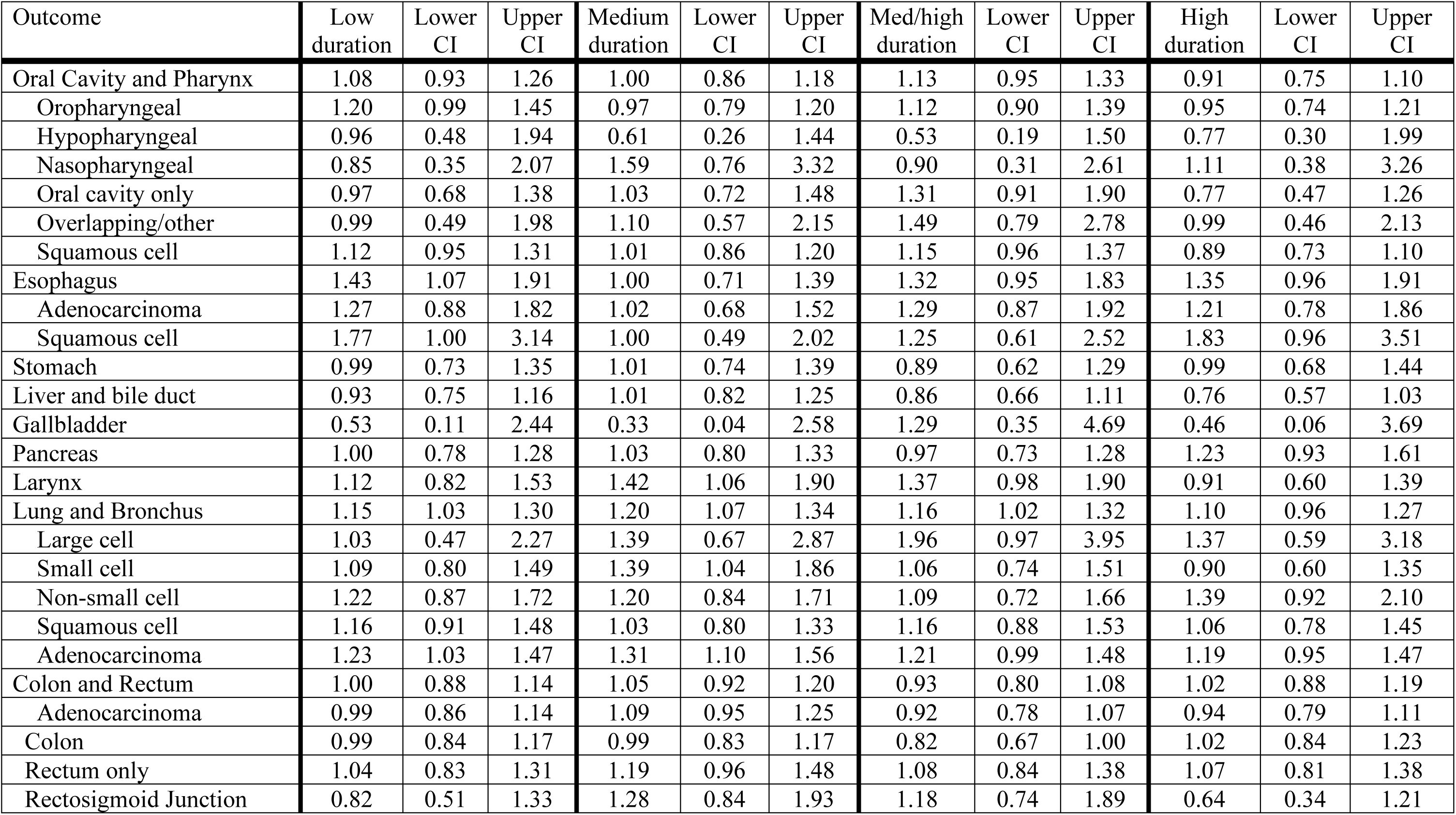
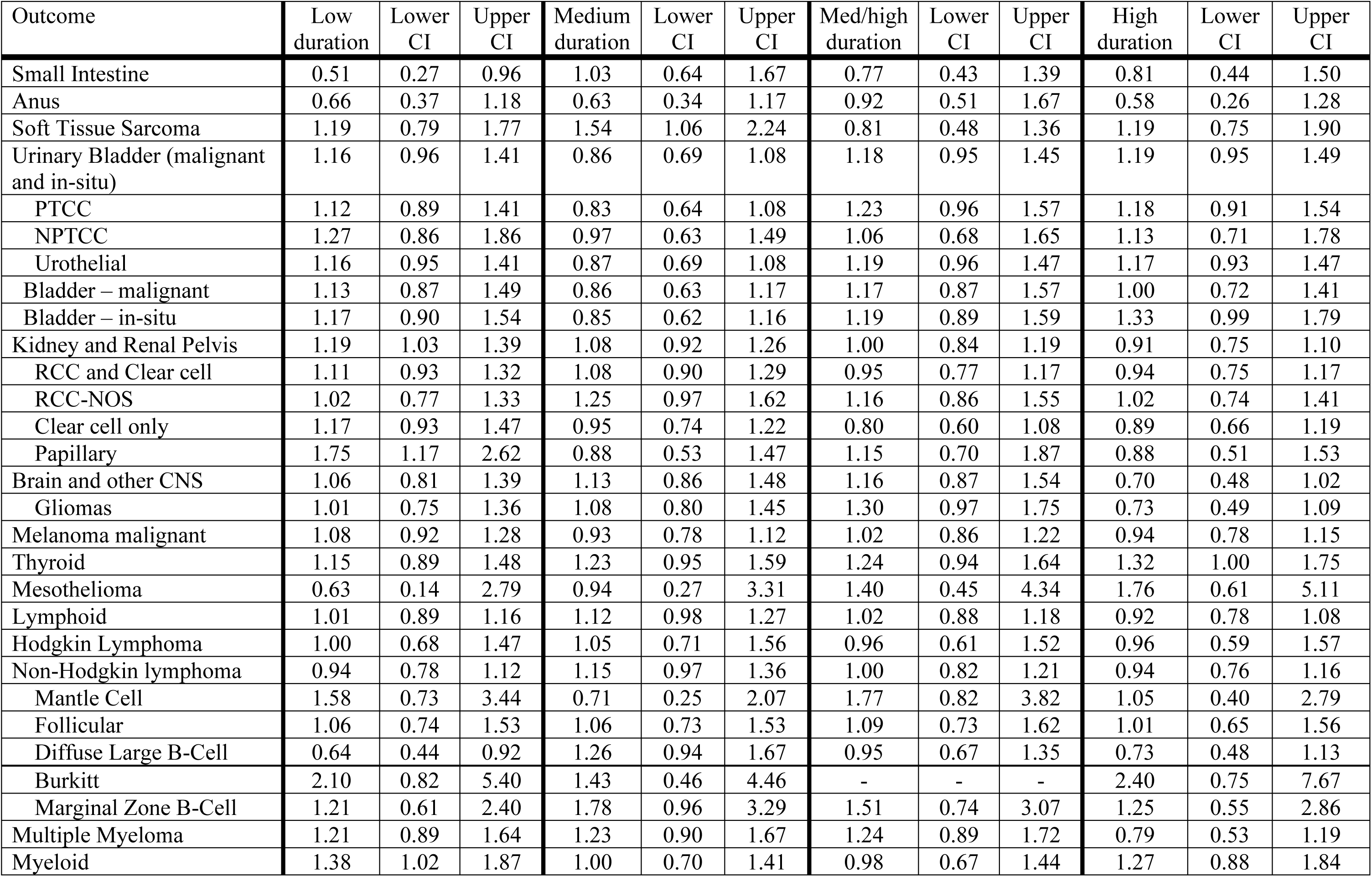
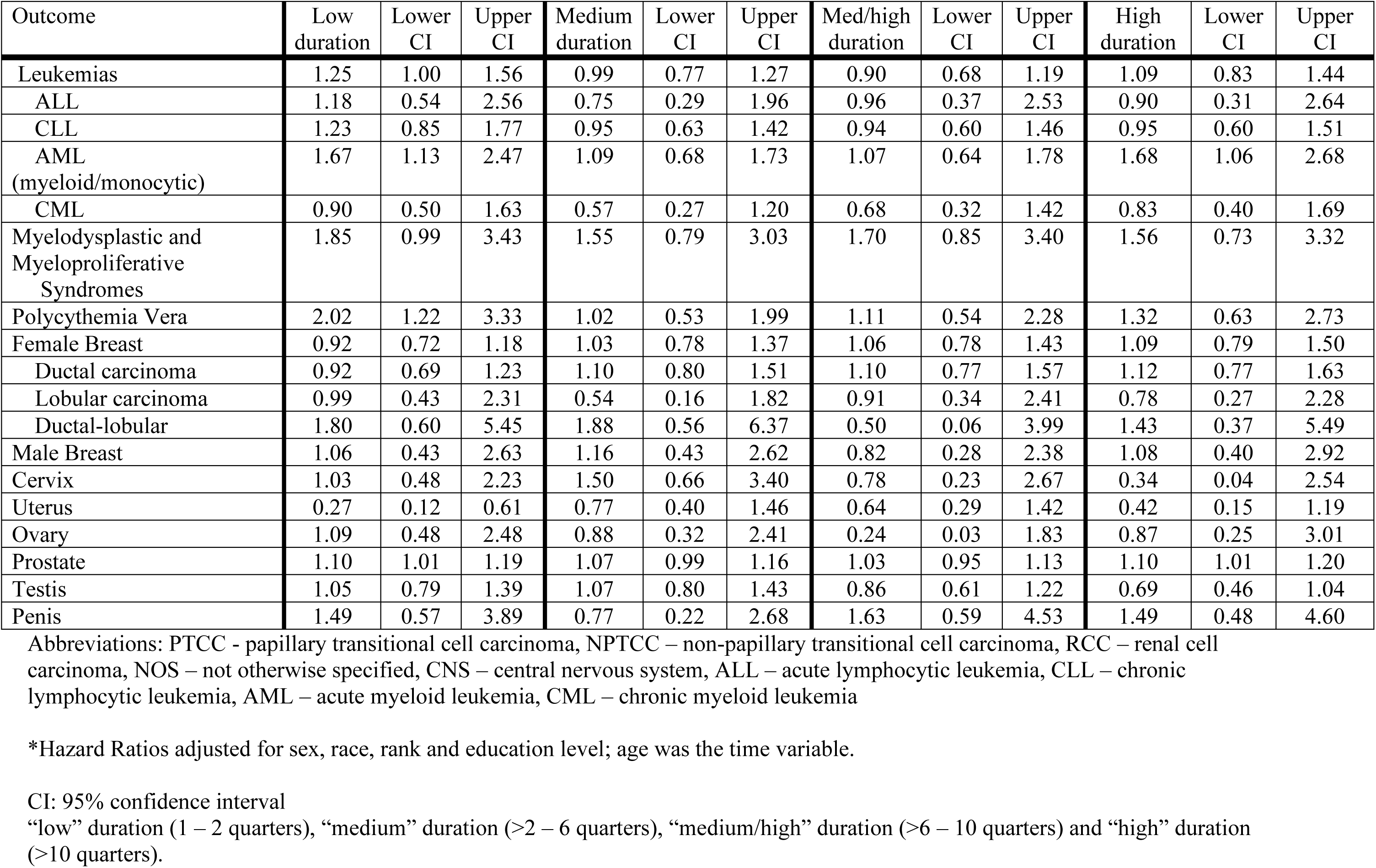
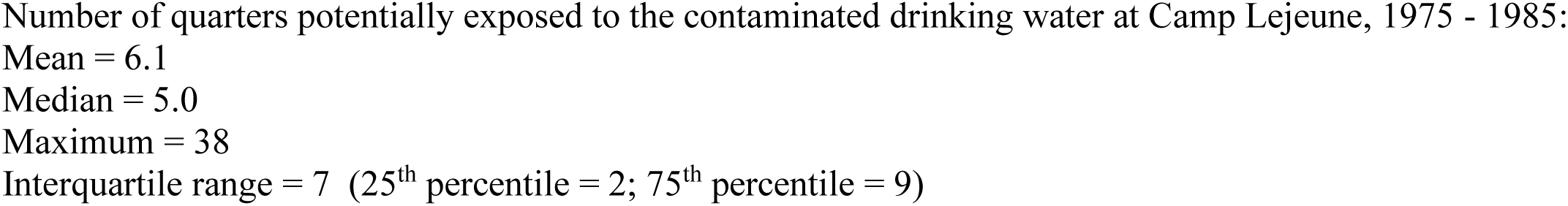
Duration stationed at Camp Lejeune (Camp Pendleton as reference): Marines/Navy personnel subgroup: Hazard ratios and 95% confidence intervals (CIs)

For civilian workers, analysis of duration of employment between October 1972 and December 1985 at Camp Lejeune with Camp Pendleton as the referent group is shown in Table 7. The levels of duration were approximately tertiles of the data after removal of the reference group. The levels of the categorical variable consisted of the number of quarters the worker was employed at Camp Lejeune between October 1972 and December 1985: “low” duration (1 – 4 quarters), “medium” duration (5 – 21 quarters), and “high” duration (22– 53 quarters). A monotonic trend was observed for diffuse large B-cell lymphoma with a HR of 1.99 though this estimate was imprecise with a 95% CIR >3.

**Table 7.** Duration employed at Camp Lejeune, October 1972 – December 1975 (Camp Pendleton as reference): Civilian workers: Hazard ratios and 95% confidence intervals (CIs)

| Outcome | Low duration | Lower CI | Upper CI | Medium duration | Lower CI | Upper CI | High duration | Lower CI | Upper CI |
| --- | --- | --- | --- | --- | --- | --- | --- | --- | --- |
| Oral Cavity and Pharynx | 1.82 | 0.76 | 4.34 | 2.20 | 1.07 | 4.53 | 1.23 | 0.56 | 2.70 |
| Oropharyngeal | 2.43 | 0.70 | 8.50 | 2.39 | 0.82 | 6.95 | 0.24 | 0.03 | 1.94 |
| Hypopharyngeal<sup>‡</sup> | 1.19 | 0.12 | 11.73 | 0.95 | 0.10 | 9.15 | 0.69 | 0.07 | 6.67 |
| Oral cavity only | 2.16 | 0.33 | 14.11 | 0.94 | 0.09 | 9.32 | 2.70 | 0.60 | 12.20 |
| Overlapping/other | 0.66 | 0.07 | 5.89 | 1.53 | 0.37 | 6.29 | 1.64 | 0.45 | 5.93 |
| Squamous cell | 2.32 | 0.90 | 6.00 | 2.91 | 1.35 | 6.25 | 1.23 | 0.51 | 2.96 |
| Esophagus | - | - | - | 0.41 | 0.09 | 1.83 | 0.75 | 0.27 | 2.05 |
| Adenocarcinoma | - | - | - | 0.26 | 0.03 | 2.08 | 0.78 | 0.24 | 2.55 |
| Stomach | 0.34 | 0.08 | 1.49 | 0.55 | 0.19 | 1.64 | 0.94 | 0.43 | 2.04 |
| Liver | 0.82 | 0.18 | 3.78 | 0.71 | 0.20 | 2.49 | 0.54 | 0.17 | 1.68 |
| Liver, bile duct, and gallbladder | 0.78 | 0.26 | 2.37 | 1.00 | 0.42 | 2.38 | 0.65 | 0.27 | 1.56 |
| Pancreas | 0.62 | 0.29 | 1.36 | 0.65 | 0.32 | 1.31 | 0.74 | 0.40 | 1.36 |
| Larynx | 2.78 | 0.93 | 8.32 | 1.05 | 0.28 | 3.93 | 0.64 | 0.19 | 2.16 |
| Lung and Bronchus | 1.05 | 0.76 | 1.43 | 1.23 | 0.95 | 1.59 | 1.15 | 0.91 | 1.44 |
| Large cell | 1.46 | 0.27 | 7.92 | 0.55 | 0.06 | 4.81 | 1.25 | 0.30 | 5.11 |
| Small cell | 1.49 | 0.76 | 2.94 | 1.14 | 0.60 | 2.17 | 0.95 | 0.52 | 1.74 |
| Non-small cell | 0.48 | 0.14 | 1.66 | 0.78 | 0.32 | 1.94 | 1.24 | 0.63 | 2.45 |
| Squamous cell | 1.58 | 0.83 | 3.02 | 1.87 | 1.12 | 3.14 | 1.52 | 0.95 | 2.41 |
| Adenocarcinoma | 0.85 | 0.49 | 1.47 | 1.41 | 0.93 | 2.15 | 1.14 | 0.77 | 1.69 |
| Colon and Rectum | 1.09 | 0.70 | 1.68 | 0.97 | 0.65 | 1.44 | 0.82 | 0.57 | 1.18 |
| Adenocarcinoma | 1.19 | 0.76 | 1.86 | 1.05 | 0.70 | 1.58 | 0.87 | 0.59 | 1.26 |
| Colon | 1.11 | 0.66 | 1.86 | 1.03 | 0.64 | 1.65 | 0.86 | 0.56 | 1.33 |
| Rectum only | 1.31 | 0.54 | 3.18 | 1.06 | 0.46 | 2.41 | 0.86 | 0.40 | 1.85 |
| Rectum and Rectosigmoid Junction | 1.03 | 0.46 | 2.31 | 1.00 | 0.50 | 2.00 | 0.72 | 0.37 | 1.42 |
| Soft Tissue Sarcoma | - | - | - | 1.83 | 0.63 | 5.29 | 0.25 | 0.03 | 2.04 |
| Urinary Bladder (malignant and in-situ) | 1.76 | 1.07 | 2.88 | 1.04 | 0.66 | 1.65 | 0.94 | 0.64 | 1.38 |
| Papillary Transitional Cell Carcinoma | 1.66 | 0.93 | 2.97 | 1.04 | 0.60 | 1.80 | 0.90 | 0.56 | 1.44 |
| Non-papillary Transitional Cell Carcinoma | 2.08 | 0.74 | 5.84 | 1.36 | 0.57 | 3.26 | 1.11 | 0.53 | 2.32 |
| Urothelial | 1.76 | 1.06 | 2.91 | 1.11 | 0.70 | 1.77 | 0.95 | 0.64 | 1.41 |
| Bladder – malignant | 1.86 | 0.95 | 3.62 | 1.06 | 0.57 | 1.99 | 0.99 | 0.59 | 1.64 |
| Bladder – in-situ | 1.77 | 0.88 | 3.58 | 1.01 | 0.51 | 1.99 | 0.93 | 0.52 | 1.65 |
| Kidney and Renal Pelvis | 0.96 | 0.49 | 1.85 | 1.08 | 0.62 | 1.89 | 1.25 | 0.77 | 2.03 |
| Renal cell and clear cell carcinoma | 0.93 | 0.43 | 1.99 | 0.90 | 0.46 | 1.76 | 1.23 | 0.71 | 2.14 |
| Renal cell carcinoma, NOS | 1.43 | 0.58 | 3.53 | 0.90 | 0.37 | 2.17 | 1.26 | 0.62 | 2.56 |
| Clear cell only | 0.38 | 0.08 | 1.70 | 0.96 | 0.34 | 2.68 | 1.28 | 0.53 | 3.11 |
| Papillary | - | - | - | 1.03 | 0.09 | 11.16 | 1.59 | 0.22 | 11.41 |
| Brain and other CNS | - | - | - | 0.76 | 0.27 | 2.13 | 0.58 | 0.19 | 1.78 |
| Melanoma | 1.17 | 0.66 | 2.08 | 0.77 | 0.43 | 1.41 | 1.15 | 0.70 | 1.90 |
| Thyroid | 1.89 | 0.84 | 4.26 | 1.86 | 0.82 | 4.20 | 2.01 | 0.84 | 4.82 |
| Mesothelioma | 0.89 | 0.10 | 8.21 | 0.71 | 0.08 | 6.40 | 1.14 | 0.24 | 5.40 |
| Lymphoid | 1.03 | 0.65 | 1.64 | 1.09 | 0.73 | 1.63 | 0.99 | 0.68 | 1.43 |
| Lymphoid excluding Hodgkin Lymphoma | 1.11 | 0.70 | 1.78 | 1.17 | 0.78 | 1.75 | 0.98 | 0.67 | 1.43 |
| Non-Hodgkin lymphoma | 1.37 | 0.79 | 2.36 | 1.22 | 0.75 | 2.01 | 1.08 | 0.68 | 1.72 |
| Follicular | 2.99 | 1.01 | 8.83 | 1.77 | 0.63 | 4.96 | 0.62 | 0.17 | 2.24 |
| Diffuse Large B-Cell | 1.01 | 0.36 | 2.83 | 1.21 | 0.50 | 2.95 | 1.99 | 0.98 | 4.04 |
| Multiple Myeloma | 0.89 | 0.28 | 2.80 | 1.36 | 0.54 | 3.43 | 0.90 | 0.35 | 2.32 |
| Myeloid cancers | 1.41 | 0.61 | 3.25 | 1.19 | 0.55 | 2.58 | 1.54 | 0.82 | 2.89 |
| Leukemias | 0.67 | 0.27 | 1.61 | 0.86 | 0.44 | 1.70 | 0.94 | 0.53 | 1.65 |
| Chronic lymphocytic leukemia | 0.26 | 0.03 | 2.07 | 0.77 | 0.25 | 2.34 | 0.64 | 0.24 | 1.72 |
| Acute myeloid leukemia<sup>¥</sup> | 0.55 | 0.07 | 4.49 | 1.52 | 0.51 | 4.56 | 1.53 | 0.59 | 3.95 |
| Chronic myeloid leukemia | 1.05 | 0.26 | 4.29 | - | - | - | 0.97 | 0.25 | 3.82 |
| Myelodysplastic and Myeloproliferative Syndromes | 2.46 | 0.69 | 8.84 | 1.56 | 0.40 | 6.15 | 1.90 | 0.63 | 5.76 |
| Female Breast | 1.18 | 0.88 | 1.59 | 1.29 | 0.97 | 1.72 | 1.05 | 0.74 | 1.48 |
| Uterus | 1.09 | 0.59 | 1.98 | 0.73 | 0.37 | 1.43 | 0.86 | 0.41 | 1.79 |
| Ovary | 0.22 | 0.06 | 0.74 | 1.00 | 0.48 | 2.08 | 1.18 | 0.53 | 2.65 |
| Prostate | 0.97 | 0.71 | 1.34 | 0.84 | 0.64 | 1.11 | 1.22 | 0.99 | 1.49 |

HRs adjusted for sex, race, blue collar work (y/n) and education level; age was the time variable.
\* Includes bladder in situ.
¥ includes acute monocytic leukemia
£ Unadjusted results only are presented because of small numbers of cases.
- No cases.
CNS: central nervous system
“low” duration (1 – 4 quarters), “medium” duration (5 – 21 quarters), and “high” duration (22– 53 quarters).
Number of quarters potentially exposed to the contaminated drinking water at Camp Lejeune, October 1972 – December 1985:
Mean = 18.6

The results of the Cox regression analyses of the negative control non-cancer diseases comparing the Camp Lejeune and Camp Pendleton civilian workers and the Marines/Navy personnel subgroup are shown in Supplemental file 2, Tables S2-1a and S2-1b. For the Marines/Navy personnel subgroup, adjusted HRs for underlying and contributing causes of death due to alcoholism, alcohol liver disease, chronic liver disease and cardiovascular disease were ≤1.00. For COPD, the adjusted HRs for underlying and contributing causes of death were 1.08 and 1.02, respectively.

Using a range of RRs from 3.00 to 5.50 for smoking and COPD [44], to fully explain the HR of 1.08 for COPD, the difference in smoking prevalence between Camp Lejeune and Camp Pendleton Marines/Navy personnel would be about 6% (Supplemental file 2, Figure 1). Adjusting for a smoking prevalence difference of 6% and assuming RRs for smoking and lung cancer and laryngeal cancer between 7.00 and 12.00, the HR of 1.16 for lung cancer would decrease to between 1.05 and 1.06 and the HR of 1.21 for laryngeal cancer would decrease to between 1.10 and 1.11 (Supplemental file 2, Figures 2 and 3). Assuming RRs for smoking and esophageal cancer around 2.5 [27, 45], the HR of 1.27 for esophageal cancer would decrease to between 1.18 and 1.25 (Supplemental file 2, Figure 4).

For the subgroup of Marines/Navy personnel, the adjusted HRs for chronic liver disease mortality as an underlying and contributing cause were 0.93 and 0.88. A recent systematic review of alcohol consumption and mortality due to liver cirrhosis found RRs of 2.65, 6.83 and 16.38 for drinking 25g/day (2 drinks/day), 50g/day (4 drinks/day) and 100g/day (8 drinks/day) compared to those who never drank alcoholic beverages [46]. A military survey conducted in 1980 found that about 30% of Marines were heavy drinkers [47].

To determine what prevalence differences in alcohol consumption between Camp Lejeune and Camp Pendleton Marines/Navy personnel would be necessary to fully explain the chronic liver disease mortality HRs of 0.93 and 0.88, a quantitative bias analysis was conducted assuming that at least 2/3 of Marines/Navy personnel at Camp Lejeune consumed ≥1 drink/day. It was also assumed that the RRs for alcohol consumption and chronic liver disease mortality ranged between 2.5 and 10 [46]. To fully explain the HRs of 0.93 and 0.88, the prevalence differences would range between 6% and 10% and between 11% and 16%, respectively (Supplemental file 2, Figures 5-6). (Assuming a lower percentage of Camp Lejeune drinkers would decrease the prevalence difference range, e.g., if only half the Marines/Navy personnel at Camp Lejeune were drinkers, then the percentage difference ranges would be 5% – 9% and 9% – 15% for chronic liver disease mortality as underlying cause and as contributing cause, respectively.)

Adjusting for an alcohol use prevalence difference of 10% between Camp Lejeune and Camp Pendleton Marines/Navy personnel, the HR of 1.47 for squamous cell esophageal cancer would increase to between 1.51 and 1.64 (Supplemental file 2, Figure 7). Adjusting for alcohol use, the HR of 1.27 for esophageal cancer would increase to between 1.30 and 1.41 (Supplemental file 2, Figure 8). Adjusting for an alcohol use prevalence difference of 10% would increase the HR of 1.21 for laryngeal cancer to between 1.22 and 1.32 (Supplemental file 2, Figure 9).

The impact of non-differential exposure assessment on the adjusted HRs for base assignment, comparing the Camp Lejeune and Camp Pendleton Marines/Navy personnel subgroup was evaluated assuming that between 10% and 25% of those assigned to Camp Lejeune were truly unexposed and virtually none of those assigned to Camp Pendleton were truly exposed (Supplemental file 2, Table S2-2a). Adjusted for exposure misclassification, the HR of 1.16 for lung cancer would increase to between 1.18 and 1.22. For laryngeal cancer the HR of 1.21 would increase to between 1.24 and 1.28. For esophageal cancer, the HR of 1.27 would increase to between 1.30 and 1.36. For AML, the HR of 1.38 would increase to between 1.42 and 1.50.

For civilian workers, the adjusted HRs for underlying and contributing causes of death due to alcoholism, alcoholic liver disease, chronic liver disease, and cardiovascular disease were ≤1.00. For COPD mortality, the underlying cause HR was ≤1.00 but the contributing cause HR was 1.05. Using a range of RRs from 3.00 to 5.50 for smoking and COPD [44], to fully explain the HR of 1.05 for COPD, the difference in smoking prevalence between Camp Lejeune and Camp Pendleton would be about 4% (Supplemental file 2, Figure 10).

Adjusting for a smoking prevalence difference of 4% between Camp Lejeune and Camp Pendleton civilian workers, and assuming RRs for smoking and lung cancer and laryngeal between 7.00 and 12.00 [27], the HR of 1.15 for lung cancer would decrease to between 1.08 and 1.09, and the HR of 1.18 for laryngeal cancer would decrease to 1.11 (Supplemental file 2, Figures 11-12). Adjusting for a smoking prevalence difference of 4% and RRs for smoking and oral cancers between 3.50 and 7.00 [27], the HR of 1.67 for oral cancers (oral cavity and pharynx) would decrease to between 1.57 and 1.59 (Supplemental file 2, Figure 13). Finally, assuming RRs for smoking and kidney cancer of between 1.20 and 1.60 [27, 48], adjusting the kidney cancer HR of 1.12 for a 4% smoking prevalence difference would decrease the HR to between 1.10 and 1.11 (Supplemental file 2, Figure 14).

For the civilian workers, the adjusted HR for chronic liver disease mortality as an underlying cause was 0.74. To determine what prevalence differences in alcohol consumption between Camp Lejeune and Camp Pendleton workers would be necessary to fully explain the HR of 0.74 for chronic liver disease mortality, a quantitative bias analysis was conducted. It was assumed that about 1/3 of the Camp Lejeune workers consumed ≥1 drink/day. Using a range of RRs between 2.5 and 10 for alcoholic consumption and chronic liver disease mortality [46], the prevalence differences would need to range between 15% and 25% (Supplemental file 2, Figure 15). (Assuming that only 20% of Camp Lejeune workers consumed ≥1 drink/day, the prevalence difference would range from 11% to 21%. Assuming a higher percentage of Camp Lejeune drinkers would increase the prevalence difference range, e.g., if 50% of Camp Lejeune workers consumed ≥1 drink/day, the prevalence difference would range from 21% to 31%.) Adjusting for an alcohol use prevalence difference of 15% between Camp Lejeune and Camp Pendleton workers, the HRs of 1.19 for female breast cancer and laryngeal cancer would increase to between 1.20 and 1.27, and between 1.20 and 1.39, respectively (Supplemental file 2, Figures 16-17). For oral cancers, the HR of 1.67 would increase to between 1.73 and 2.11 (Supplemental file 2, Figure 18).

The analysis of the impact of non-differential exposure assessment on the adjusted HRs comparing Camp Lejeune and Camp Pendleton civilian workers used sensitivity values of 0.99 and 1.00 and specificity values ranging from 0.81 to 0.91. The chosen values for sensitivity and specificity reflected the assumptions that between 75% and 90% of those stationed or employed at Camp Lejeune were truly exposed, and all (or virtually all) of those stationed or employed at Camp Pendleton were truly unexposed. Based on these values for sensitivity and specificity, the HRs for oral cancers and cancers of the lung, larynx, kidney, female breast and NHL were adjusted for non-differential exposure misclassification (Supplemental file 2, Table S2-2b). Adjusted for exposure misclassification, the HR for lung cancer of 1.15 would increase to between 1.16 and 1.19. For laryngeal cancer, the HR of 1.18 would increase to between 1.20 and 1.23. For oral cancers, the HR of 1.67 would increase to between 1.73 and 1.85. For kidney cancer, the HR of 1.12 would increase to between 1.14 and 1.16, and the HRs of 1.19 for NHL and female breast cancer would increase to between 1.21 and 1.24.

## Discussion

This cohort study evaluated whether Marines/Navy personnel and civilian workers stationed or employed at Camp Lejeune during a portion of the period when the drinking water was contaminated had increased risks of cancers during the period from 1996 to 2017 compared to being stationed or employed at Camp Pendleton. Additional analyses evaluated duration stationed or employed at Camp Lejeune with Camp Pendleton as the reference group. These analyses of duration assumed that contamination levels did not fluctuate greatly from month to month between 1972 and 1985 for the workers and between 1975 and 1985 for the Marines/Navy personnel. However, the estimated monthly average contaminant levels in the Hadnot Point and Tarawa Terrace distribution systems varied widely. Therefore, the results of the duration analyses should be interpreted with caution.

In the Cox regression analyses of the Marines/Navy personnel subgroup, several cancers had HRs ≥1.20 with 95% CIR ≤3, including AML, cancers of the esophagus, larynx, thyroid, and soft tissue, all myeloid cancers (including polycythemia vera), and the lung cancer histological subtypes, non-small cell, large cell, and adenocarcinoma. HRs ≥1.20 with 95% CIR ≤3 were also observed for myelodysplastic and myeloproliferative syndromes, polycythemia vera, MZBCL, and squamous cell esophageal cancer. A monotonic trend for thyroid cancer with longer duration at Camp Lejeune was consistent with the elevated HR for thyroid cancer observed in the Marines/Navy personnel subgroup.

In the Cox regression analysis comparing Camp Lejeune and Camp Pendleton civilian workers, cancers with HRs ≥1.20 with 95% CIR ≤3 were observed for all myeloid cancers (including polycythemia vera), squamous cell lung cancer and female ductal breast cancer. Several other cancers had HRs ≥1.20 but with 95% CIRs >3. These included oral cancers, cancers of the thyroid and male breast, and acute myeloid leukemia, myelodysplastic and myeloproliferative syndromes, follicular and diffuse large B-cell lymphomas, and non-papillary transitional cell bladder carcinoma.

In the comparisons between Camp Lejeune and Camp Pendleton workers and Marines/Navy personnel, the HRs for AML and myelodysplastic and myeloproliferative syndromes were greater than 1.20. The grouping of all myeloid cancers (including polycythemia vera) had HRs greater than 1.20 in the comparisons between Camp Lejeune and Camp Pendleton workers and Marines/Navy personnel. AML is known to be caused by benzene exposure [8]. ATSDR previously concluded that the evidence for a causal association between TCE and AML was at least as likely as not based on TCE’s effects on the immune system [15]. Benzene exposure has also been associated with myelodysplastic syndrome [49–50]. Another blood cancer, polycythemia vera, had a HR greater than 1.20 for Marines/Navy personnel, but the HR for civilian workers could not be calculated because there were 3 cases among Camp Lejeune workers and no cases among Camp Pendleton workers. Benzene exposure is possibly associated with polycythemia vera [51].

Thyroid cancer had HRs ≥1.20 for Camp Lejeune Marines/Navy personnel and civilian workers compared to Camp Pendleton. The finding for the subgroup of Marines/Navy personnel was supported by a monotonic trend with duration at Camp Lejeune. Thyroid cancer has been associated with occupational exposures to solvents (e.g., benzene), particularly in the footwear industry, among women but not men [52]. However, a review of occupations and thyroid cancer concluded that the findings for solvents were “largely null” but recommended additional study [53].

Although NHL had a HR of 1.01 for Marines/Navy personnel, several of its histological subtypes had HRs ≥1.20. In the analysis of civilian workers, NHL had a HR of 1.19. Adjusting for non-differential exposure misclassification in the civilian workers analysis, the HR for NHL would have increased above 1.20 (Supplemental file 2, Table S2-2b). In addition, HRs >1.20 were observed for follicular and diffuse large B-cell lymphomas, though 95% CIRs were >3. ATSDR previously concluded that the evidence for a causal association between TCE and NHL and between benzene exposure and NHL was sufficient [15]. Both TCE and benzene exposures have also been associated with some of the histological subtypes of NHL [54–56].

Soft tissue cancer had a HR of 1.21 with 95% CIR ≤3 in the subgroup analyses comparing Camp Lejeune and Camp Pendleton Marines/Navy personnel. Soft tissue cancer had a HR of 1.38 (95% CI: 0.73, 2.64) in the previous Camp Lejeune mortality study of Marines/Navy personnel [12]. Studies of occupational exposures to PCE or TCE and soft tissue cancer have generally included a small number of cases. Two studies found elevated risks among females only for TCE [57] and working as a dry cleaner [58]. Two other studies that did not conduct sex-specific analyses observed elevated risks for soft tissue cancer and PCE exposure [59] and both PCE and TCE exposures [60], but these findings were based on few cases.

Kidney cancer is known to be associated with TCE exposure [5]. In the current study the HRs were ≤1.20 in the comparisons between Camp Lejeune and Camp Pendleton. Papillary cell kidney cancer had a HR of 1.18 with 95% CIR ≤ 2 (95% CI 0.86-1.60) in the Marines/Navy personnel subgroup, and renal cell carcinoma NOS had a HR of 1.18 with 95% CIR > 3 in the analysis of civilian workers.

Male breast cancer had a HR ≥1.20 with 95% CIR ≤3 only in the full cohort analysis of Marines/Navy personnel (HR=1.24, 95% CI: 0.79, 1.93) which had almost double the number of cases than the subgroup analysis. A possible reason for the greater number of male breast cancer cases in the full cohort was that a much greater percentage (41.3%) in the full cohort were ≥60 years of age at the end of follow-up compared to the subgroup (23.6%). In the U.S., about 75% of male breast cancers are diagnosed at age ≥60 years [61]. In the analysis of civilian workers, there were seven cases of male breast cancer among Camp Lejeune workers compared to one case among Camp Pendleton workers. Occupational TCE exposure has been associated with male breast cancer in three studies [57, 62–63]. In a case-control study of male breast cancer using data from the U.S. Department of Veterans Affairs cancer registry, men stationed at Camp Lejeune had an odds ratio of 1.14 (95% CI: 0.65, 1.97) compared to Marines at all other bases [14].

Female breast cancer had a HR of 1.00 in the analysis of Marines and Navy personnel, but its histological subtype duct-lobular carcinoma had a HR ≥1.20. In the analysis of civilian workers, female breast cancer had a HR of 1.19. Adjusting for non-differential exposure misclassification, the HR for female breast cancer would increase to above 1.20 (Supplemental file 2, Table S2-2b). Moreover, the female breast cancer HR of 1.19 would also increase if adjusted for possible confounding due to alcohol consumption (Supplemental file 2, Figure 16). The female ductal carcinoma breast cancer had an adjusted HR of 1.32 with 95% CIR ≤3.

Some occupational studies of female breast cancer incidence and mortality have not supported a causal association with exposures to TCE, PCE, vinyl chloride, or benzene [15]. However, one case-control study found an increased risk of female breast cancer among pre-menopausal women who predominantly worked in dry cleaning [64]. A study of exposure to PCE-contaminated drinking water in Cape Cod, MA found an increased risk for breast cancer among women with the highest cumulative exposures [65]. Two recently published occupational studies of female breast cancer provide support for a causal association with TCE and/or PCE exposure [35–36]. A study in Taiwan found elevated risks for female breast cancer among workers exposed to TCE/PCE and benzene [35]. A case-control study of postmenopausal women found increased ORs for occupationally ever exposed to benzene and PCE and postmenopausal breast cancer ranging between 1.18 and 1.32 and between 1.92 and 2.83, respectively, (with the ranges depending on the adjustment model), but with 95% CIRs >3 [36].

Several smoking associated cancers that have also been linked to exposures to TCE, PCE, and/or benzene were evaluated in this study including oral cancers and cancers of the esophagus, bladder, larynx, and lung. Meta-analyses have found relative risks for these cancers associated with smoking ≥2.50. [27, 48].

Oral cancers had a HR ≥1.20 in the analysis of civilian workers (95% CI 0.93, 3.00). There is some evidence linking PCE and TCE occupational exposures and oral cancers among females [30], but the evidence is much weaker for males [31]. Dry cleaning workers had a standardized mortality ratio of 1.10 for mortality due to cancers of the buccal cavity and pharynx [66]. Occupational benzene exposure has not been associated with oral cancers [67–68].

In the subgroup analyses of Marines/Navy personnel, the HR for esophageal cancer was 1.27 with 95% CIR ≤3, and the HR for squamous cell esophageal cancer was 1.47 with 95% CIR ≤3. Three occupational cohort studies have found associations between TCE exposures and esophageal cancer [15]. In addition, a previous cohort mortality study comparing Marines/Navy personnel stationed at Camp Lejeune versus Camp Pendleton obtained a HR of 1.43 (95% CI: 0.85, 2.38) for esophageal cancer [12].

In the analysis of civilian workers, the bladder cancer histological subtype non-papillary transitional cell carcinoma had a HR ≥1.20 with 95% CIR >3 (HR=1.30, 95% CI 0.70, 2.40). In the Marines/Navy personnel subgroup analysis of duration at Camp Lejeune, a HR ≥1.20 with 95% CIR ≤ 3 (HR=1.33, 95% CI 0.99, 1.79) was only observed in the high duration category for bladder cancer in situ. Occupational exposure to PCE is associated with bladder cancer [15].

For laryngeal cancer, HRs were 1.21 with 95% CIR ≤3 (95% CI 0.98, 1.50) for the Marines/Navy personnel subgroup and 1.18 with 95% CIR >3 (95% CI 0.49, 2.82) for civilian workers. Laryngeal cancer has been associated with occupational exposure to PCE in men [31] and with occupational PCE and TCE exposure in women [30]. In a previous study, an odds ratio of 1.29 was found for men who were ever occupationally exposed to PCE or who were exposed to the low PCE cumulative exposure index [31].

The lung cancer histological subtypes large cell, non-small cell, and adenocarcinoma had HRs ≥1.20 with 95% CIR ≤3 in the analysis of the Marines/Navy personnel subgroup. Non-small cell lung cancer also had a HR ≥1.20 in the high duration category. In the analysis of civilian workers, squamous cell lung cancer had a HR ≥1.20 with 95% CIR ≤3. Occupational exposure to PCE in dry cleaning has been associated with lung cancer in three cohort studies and one case-control study with relative risks in the range of 1.3 and 1.4 [28, 58, 69–70]. Two case-control studies of occupational exposure to PCE also found associations with lung cancer, especially in women [29, 71]. In addition, a study of drinking water exposures to PCE at Cape Cod, MA found an odds ratio of 3.7 for lung cancer among those with the highest cumulative exposure [18]. A case-control study of lung cancer and occupational exposures to benzene, toluene and xylene found an association for benzene with an odds ratio of 1.35 (95% CI: 0.99, 1.84) [32].

This study did not have information on important risk factors such as smoking and alcohol consumption since these are not routinely collected by cancer registries. However, confounding due to failure to adjust for unmeasured risk factors was likely to be minor because of the demographic and socio-economic similarity of the Camp Lejeune and Camp Pendleton cohorts. The prevalence of smoking and “heavy alcohol” consumption among Marines in 1980 was estimated at 53.4% and 28.6%, respectively [47]. Marines had a smoking prevalence slightly less than the Navy and Army but had the highest heavy alcohol consumption prevalence among the services [47]. Smoking and alcohol consumption among Marines were encouraged by the military culture, the stress of service, targeted advertising by the tobacco and alcoholic beverage industry, and the lower cost and tax-free availability of these products on base at both Camp Lejeune and Camp Pendleton compared to off-base civilian stores [47, 72].

In the subgroup analysis of Marines/Navy personnel, the HRs for COPD and cardiovascular mortality were 1.08 and 0.99, suggesting minor if any difference in smoking behavior between Camp Lejeune and Camp Pendleton (Supplemental file 2, Table S2-1a). On the other hand, the HRs for mortality due to alcoholism, alcoholic liver disease and chronic liver disease as underlying causes were 0.90, 0.86, and 0.93 suggesting that the prevalence of alcohol use among Camp Lejeune Marines/Navy personnel may be lower than among Camp Pendleton Marines/Navy personnel (Supplemental file 2, Table S2-1a). For civilian workers, the HRs for COPD as an underlying and contributing cause of mortality were 0.91 and 1.05, and the HRs for cardiovascular disease were ≤1.00 suggesting minor if any difference in smoking behavior between Camp Lejeune and Camp Pendleton workers. On the other hand, the HRs for mortality due to alcoholism, alcoholic liver disease, and chronic liver disease were 0.62, 0.54 and 0.74, suggesting the prevalence of alcohol use among Camp Lejeune workers may have been lower than among Camp Pendleton workers.

For smoking to fully explain the HRs observed for cancers of the lung and larynx in the analyses of Marines/Navy personnel and civilian workers, a difference of ≥10% in smoking prevalence would be necessary (see Supplemental file 2, Figures 2-3, 11-12). Given the similarity of the two bases, a percentage difference of this magnitude in the prevalence of smoking was unlikely. Based on the findings for COPD mortality, it is more likely that the difference in smoking prevalence between Camp Lejeune and Camp Pendleton Marines/Navy personnel and civilian workers is between 4% and 6% (Supplemental file 2, Figures 1, 10). Adjusting for a smoking prevalence difference of 4% or 6% would reduce the HRs for the smoking-related cancers by less than 10% (Supplemental file 2, Figures 2-4, 11-14).

The findings for the negative control diseases for alcohol consumption, i.e., mortality due to alcoholism, alcoholic liver disease and chronic liver disease, suggest that Camp Lejeune Marines/Navy personnel and civilian workers had a lower prevalence of alcohol use than Camp Pendleton. The findings for these negative controls suggest that possible confounding due to alcohol consumption might have biased HRs towards the null for alcohol-related cancers such as oral cancers and cancers of the esophagus, larynx, and female breast. For laryngeal cancer, adjusting for possible differences in alcohol consumption between the two bases might cancel out the impact of adjusting for possible smoking differences between the two bases (Supplemental file 2, Figures 3, 9, 12, 17.). Similarly, for oral cancers among workers, and esophageal cancer among Marines/Navy personnel, the impact on the HRs of adjusting for alcohol use might cancel out the impact of adjusting for smoking. (Supplemental file 2, Figures 4, 8, 13 and 18).

To evaluate the potential impact of non-differential and independent exposure misclassification, the sensitivity of the exposure classification, i.e., the probability that the truly exposed were correctly classified as exposed (i.e., assigned to Camp Lejeune) was assumed to be near 1.0. The specificity of the exposure classification i.e., the probability that the truly unexposed were correctly classified as unexposed (i.e., assigned to Camp Pendleton) was assumed to range between 0.81 to 0.91. Adjusting for exposure misclassification using these values for sensitivity and specificity would increase the HRs by no more than 10% (Supplemental file 2, Tables 2a-2b). These results suggested that for cancers that are smoking-related, the bias due to non-differential exposure misclassification in this study may cancel out the potential confounding bias due to smoking.

Overall, the results of the quantitative bias analyses suggested that in this study, the impacts of adjusting for confounding by smoking or alcohol consumption, and adjusting for non-differential exposure misclassification, would likely be minor and may cancel each other. In particular, for cancers that are both smoking-related and alcohol-related, the impact of potential confounding bias due to smoking may be more than counteracted by the impact of potential confounding bias due to alcohol consumption as well as the bias due to exposure misclassification.

A major strength of this study was the collection of cancer incidence data from every state and territorial cancer registry, the D.C. registry, the VA cancer registry, and the DOD cancer registry. Collecting data from all these cancer registries was necessary because the Marines/Navy personnel resided in every state. Moreover, unlike the National Death Index, there is no central cancer incidence registry in the US that can provide individual-level cancer incidence data linked to the personal identifier information of persons in a study.

Another major strength was the evaluation of histological subtypes for several of the cancer types including hematopoietic cancers and cancers of the lung, esophagus, oral cavity, kidney, bladder, and female breast cancer. The epidemiological findings of associations with exposures to certain chemicals such as those found in the drinking water at Camp Lejeune have differed among the histological subtypes of hematopoietic cancers [55, 73], lung cancer [74], and head and neck cancers [31]. It is possible that differences in associations may also occur among the histological subtypes of other cancers. In this study, both cancer types and histological subtypes were evaluated.

Weaknesses of this study included several sources of non-differential exposure misclassification bias as well as the lack of information on smoking, alcohol consumption, and the occupations prior to and after active-duty service or employment at Camp Lejeune and Camp Pendleton. In addition, many of the HRs in the analyses had 95% CIRs >3 due predominantly to the small numbers of cases for the rare cancers and histological subtypes. In particular, many HRs in the analyses of the civilian workers had 95% CIRs >3 due to small numbers of cases.

Many of the HRs observed in this study were less than 1.50. This result was not unexpected because the exposures to the drinking water contamination at Camp Lejeune were likely lower and of shorter duration than occupational exposures to these chemicals. Nevertheless, risk estimates for many of these cancers from occupational exposures to these chemicals also tend to be less than 1.50. For example, the HR of 1.21 for laryngeal cancer in the subgroup analysis comparing Camp Lejeune and Camp Pendleton Marines/Navy personnel was similar in size to the odds ratio of 1.29 for ever/never occupational exposure to PCE among males in a case-control study conducted in France [31]. Three meta-analyses of occupational exposures to TCE and kidney cancer found relative risks in the 1.3 to 1.4 range [15]. A meta-analysis of TCE and NHL observed a summary relative risk of 1.32 [75]. The meta-analysis of occupational exposure to PCE and bladder cancer found a RR of 1.08 for PCE-exposed workers and a RR of 1.47 for employment as a dry cleaner [76]. A meta-analysis of occupational benzene exposure and NHL found a summary relative risk of 1.27 for those studies that had quantitative exposure assessments [77].

An additional factor affecting both the magnitude of the HRs and the 95% CIRs in the subgroup analyses of the Marines/Navy personnel was that at the end of follow-up, the median age was 57 years and over 75% of the subgroup members were under the age of 60 years. According to the NCI’s SEER Program, between 2014 and 2018 the median age of a cancer diagnosis was 66 years [78]. For cancers of the bladder, lung, pancreas, and gallbladder, as well as chronic lymphocytic leukemia and myelodysplastic syndrome, the median age at diagnosis is ≥70 years. For cancers that have been associated with occupational TCE exposure such as NHL, and cancers of the kidney and liver, the median ages at diagnosis are 67, 64, and 65 years, respectively. For several other cancers that have been associated with occupational exposures to TCE or benzene, such as AML and multiple myeloma, the median ages at diagnosis are 68 and 69 years, respectively [78].

## Conclusion

In the analyses of the Marines/Navy personnel subgroup, adjusted HRs ≥1.20 with 95% CIRs ≤3 were observed for all myeloid cancers including polycythemia vera, AML, myelodysplastic and myeloproliferative syndromes, polycythemia vera, cancers of the esophagus, larynx, thyroid, and soft tissue, and the histological subtypes marginal zone B-cell lymphoma, squamous cell esophageal cancer and the lung cancer subtypes large cell, non-small cell and adenocarcinoma. The finding for thyroid cancer was supported by a monotonic trend for duration at Camp Lejeune. In the full cohort of Marines/Navy personnel, male breast cancer had an adjusted HR ≥1.20 with a 95% CIR ≤3.

In the analyses of civilian workers, adjusted HRs ≥1.20 with 95% CIRs ≤3 were observed for all myeloid cancers including polycythemia vera, and the histological subtypes squamous cell lung cancer and female ductal breast cancer. Adjusted HRs ≥1.20 that did not meet the criterion for precision (i.e., 95% CIRs >3) due to small numbers of cases included oral cancers, thyroid cancer, AML, myelodysplastic and myeloproliferative syndromes, follicular and diffuse large B-cell lymphomas, and non-papillary transitional cell bladder carcinoma. NHL and female breast cancer had adjusted HRs of 1.19 with 95% CIRs ≤3.

Few studies have evaluated drinking water exposures to these chemicals and cancer incidence. The adult cancer incidence of the family members of the Marines and Navy personnel who resided in base family housing at Camp Lejeune has not been evaluated. Families living in base housing that received contaminated drinking water may have had exposure durations that were longer than most Marines and Navy personnel on base. The results of this study are relevant to all individuals exposed to the contaminated drinking water at Camp Lejeune and add to the literature on the health effects of these contaminants. It is hoped that this study encourages future research on the health effects of drinking water exposure to these chemicals.

## Authors’ contributions

FJB designed the study, oversaw the data collection, managed, analyzed and interpreted the data, and prepared the manuscript. Battelle and NAACCR staff recruited the cancer registries, designed and oversaw the data collection, conducted data linkages for some of the registries and managed the data.

## Data Availability

All data referred to in the manuscript are available upon reasonable request from ATSDR Office of Science.

## Abbreviations

ATSDR: Agency for Toxic Substances and Disease Registry
AML: acute myeloid leukemia
BOQ: bachelor officer quarters
CDC: Centers for Disease Control and Prevention
CI: confidence interval
CIR: confidence interval ratio
COPD: chronic obstructive pulmonary disease
DCE: t-1,2-dichloroethylene
DMDC: Defense Manpower Data Center
DOD: US Department of Defense
EPA: US Environmental Protection Agency
HB: Holcomb Boulevard treatment plant
HP: Hadnot Point treatment plant
HR: hazard ratio
IARC: International Agency for Research on Cancer
ICD-O-3: third edition of the International Classification of Diseases for Oncology
MCL: EPA maximum contaminant level in drinking water
MZBCL: marginal zone B-cell lymphoma
NHL: non-Hodgkin lymphoma
NOS: not otherwise specified
NTP: National Toxicology Program
µg/L: micrograms per liter
PCE: tetrachloroethylene (also known as perchloroethylene)
RR: risk ratio
SEER: Surveillance, Epidemiology, and End Results Program
SIR: Standardized incidence ratio
SSA: Social Security Administration
SSN: Social security number
TCE: trichloroethylene
TT: Tarawa Terrace treatment plant
USMC: United States Marine Corps
VA: U.S. Department of Veteran Affairs
WHO: World Health Organization

## Acknowledgement

The author would like to thank the following lead project staff of Battelle Memorial Institute who coordinated the data collection from the cancer registries and provided data management support: April Greek (Project Director), Ruth Gatiba (Project Manager), Rona Boehm (Data Management team lead), Gene Shin (Registry Outreach team lead), and the supporting staff at Battelle. The author would also like to thank lead project staff of the North American Association of Central Cancer Registries (NAACCR) who also coordinated data collection: Betsy Kohler (Assistant Project Director), Recinda Sherman (registry linkage coordinator) and supporting staff at NAACCR. Others who assisted the data collection effort included Donald Green, William Howe, and Richard Lee from the Information Management Services, Inc.

Essential to the study was the participation of the 55 state, federal and territorial cancer registries who conducted the data linkages and provided the cancer incidence data. Assistance during the early stages of the study was provided by ATSDR/CDC staff: Perri Ruckart, Scott van Heest, Geoffrey Whitfield, and Joseph Ralph. Aaron Bernstein, director of NCEH and ATSDR, provided editing assistance. Finally, the author would like to acknowledge the strong and essential support for the study by the Camp Lejeune Community Assistance Panel members.

This work was supported by funding through interagency agreements with the U.S. Department of Health and Human Services’ Agency for Toxic Substances and Disease Registry and the U.S. Department of the Navy. The author did not receive payment or services from a third party for any aspect of the submitted work.

## ATSDR/CDC Disclaimer

The findings and conclusions in this manuscript are those of the author and do not necessarily represent the official position of the Centers for Disease Control and Prevention/Agency for Toxic Substances and Disease Registry.

