## Supplemental tables and figures for "Evaluation of cancer incidence among Marines and Navy personnel and civilian workers exposed to contaminated drinking water at USMC Base Camp Lejeune: a cohort study"

### Supplemental File 1

Table S1-1. Surveillance, Epidemiology, and End Results (SEER) recodes, ICD-10 codes and Morphology codes (ICD O-3)

| Cancer Outcome | SEER Recodes, ICD-10 codes &/or Morphology Codes (ICD O-3) |
|---|---|
| Oral Cavity and Pharynx | SEER recodes 20010 – 20100 (excluding ICD O-3 codes: 9050-9055, 9140, 9590-9992) |
| Oropharynx | ICD-10 codes: C01.9, C02.4, C05.1-2, C09.0-1, C09.8-9, C10.0-4, C10.8-9 |
| Hypopharynx | ICD-10 codes: C12.9, C13.0-2, C13.8-9 |
| Nasopharynx | ICD-10 codes: C11.0-9 |
| Oral cavity only | ICD-10 codes: C00.0–9; C02.0–3; C03.0–1; C03.9, C04.0–1; C04.8–9; C05.0; C06.0–2; C06.8–9 |
| Overlapping/other | ICD-10 codes: C02.8-9, C05.8-9, C07.9, C08.0-1, C14.0, C14.2, C14.8 |
| Squamous cell oral cancer | SEER recodes 20010 – 20100 and ICD O-3 codes: 8070 – 8074, 8076, 8083, 8560 |
| Esophagus | SEER recode 21010 (excluding ICD O-3 codes: 9050-9055, 9140, 9590-9992) |
| Adenocarcinoma | ICD O-3 codes: 8140, 8143–44, 8255, 8260, 8480–81, 8560 |
| Squamous cell | 8070–72, 8083–8084, 8560 |
| Stomach | SEER recode: 21020 (excluding ICD O-3 codes: 9050-9055, 9140, 9590-9992) |
| Liver and bile duct | SEER recodes: 21071 and 21072 (excluding ICD O-3 codes: 9050-9055, 9140, 9590-9992) |
| Gallbladder | SEER recode: 21080 (excluding ICD O-3 codes: 9050-9055, 9140, 9590-9992) |
| Pancreas | SEER recode: 21100 (excluding ICD O-3 codes: 9050-9055, 9140, 9590-9992) |
| Larynx | SEER recode: 22020 (excluding ICD O-3 codes: 9050-9055, 9140, 9590-9992) |
| Lung and Bronchus | SEER recode: 22030 (excluding ICD O-3 codes: 9050-9055, 9140, 9590-9992) |
| Large cell | ICD O-3 codes: 8012-8014, 8021, 8034, 8082 |
| Small cell | ICD O-3 codes: 8002-8005, 8041-8045 |
| Non-small cell | ICD O-3 code: 8046 |
| Squamous cell | ICD O-3 codes: 8052, 8070-8076, 8078, 8083-8084, 8094, 8120, 8123, 8560 |
| Adenocarcinoma | ICD O-3 codes: 8050, 8140-8141, 8144, 8201, 8250-8255, 8260, 8290, 8310, 8320, 8323, 8333, 8470, 8480-8481, 8490, 8507, 8550, 8560, 8570, 8572, 8574, 8576 |
| Colon and Rectum | SEER recodes: 21041 – 21052 (excluding ICD O-3 codes: 9050-9055, 9140, 9590-9992) |
| Adenocarcinoma | ICD O-3 codes: 8140, 8210-8211, 8220-8221, 8245, 8255, 8261-8263, 8480-8481, 8560, 8574 |
| Colon | SEER recodes: 21041 - 21049 |
| Rectum only | SEER recode: 21052 |
| Rectosigmoid Junction | SEER recode: 21051 |
| Small Intestine | SEER recode: 21030 (excluding ICD O-3 codes: 9050-9055, 9140, 9590-9992) |
| Anus | SEER recode: 21060 (excluding ICD O-3 codes: 9050-9055, 9140, 9590-9992) |

| Cancer Outcome | SEER Recodes, ICD-10 codes &/or Morphology Codes (ICD O-3) |
|---|---|
| Urinary Bladder (malignant and in-situ) | SEER recode: 29010 (excluding ICD O-3 codes: 9050-9055, 9140, 9590-9992) |
| Papillary Transitional Cell Carcinoma | ICD O-3 codes: 8050, 8121, 8130-8131 |
| Non-papillary Transitional Cell Carcinoma | ICD O-3 codes: 8120, 8122 |
| Urothelial | ICD O-3 codes: 8050, 8120-8122, 8130-8131 |
| Bladder – malignant | SEER recode: 29010, behavior code = 3 |
| Bladder – in-situ | SEER recode: 29010, behavior code = 2 |
| Kidney and Renal Pelvis | SEER recode: 29020 (excluding ICD O-3 codes: 9050-9055, 9140, 9590-9992) |
| Renal cell and clear cell carcinoma | ICD O-3 codes: 8310, 8312 |
| Renal cell carcinoma, NOS | ICD O-3 code: 8312 |
| Clear cell only | ICD O-3 code: 8310 |
| Papillary | ICD O-3 code: 8260 |
| Brain and other CNS | SEER recodes: 31010 and 31040 (excluding ICD O-3 codes: 9050-9055, 9140, 9590-9992) |
| Gliomas | ICD O-3 codes: 9380–9384, 9391–9460 |
| Soft Tissue Sarcoma | SEER recode: 24000 (excluding ICD O-3 codes: 9050-9055, 9140, 9590-9992) |
| Melanoma | SEER recode: 25010 (ICD-10 codes: C44.0-44.9; ICD O-3 codes: 8720-8790) |
| Thyroid | SEER recode: 32010 (excluding ICD O-3 codes: 9050-9055, 9140, 9590-9992) |
| Mesothelioma | SEER recode: 36010, ICD O-3 codes: 9050-9055 |
| Leukemias | SEER recodes: 35011 - 35043 |
| Lymphoid cancers | SEER recodes: 33011-33012, 33041-33042, 34000, 35011-35013 |
| Hodgkin lymphoma | SEER recodes: 33011-33012 |
| Non-Hodgkin lymphoma | SEER recodes: 33041-33042 |
| Mantle Cell | ICD O-3 code: 9673 |
| Follicular | ICD O-3 codes: 9690-9691, 9695, 9698 |
| Diffuse Large B-cell | ICD O-3 code: 9680 |
| Burkitt | ICD O-3 codes: 9687, 9826 |
| Marginal Zone B-cell | ICD O-3 code: 9699 |
| Multiple Myeloma | SEER recode: 34000 |
| Acute lymphocytic leukemia | SEER recode: 35011 |
| Chronic lymphocytic leukemia | SEER recode: 35012 |
| Myeloid cancers | SEER recodes: 35021-35023, 35031, and ICD O-3 codes: 9960, 9975, 9980, 9982-9986, 9989 |
| Acute myeloid leukemia[¥] | SEER recodes: 35021, 35031 |

| Cancer Outcome | SEER Recodes, ICD-10 codes &/or Morphology Codes (ICD O-3) |
|---|---|
| Chronic myeloid leukemia | SEER recode: 35022 |
| Myelodysplastic and myeloproliferative syndromes | ICD O-3 codes: 9960, 9975, 9980, 9982-9986, 9989 |
| Polycythemia Vera | ICD O-3 code: 9950 |
| Female Breast | SEER recode: 26000 (excluding ICD O-3 codes: 9050-9055, 9140, 9590-9992) |
| Ductal carcinoma | ICD O-3 code: 8500 |
| Lobular carcinoma | ICD O-3 code: 8520 |
| Duct-lobular carcinoma | ICD O-3 code: 8522 |
| Male Breast | SEER recode: 26000 (excluding ICD O-3 codes: 9050-9055, 9140, 9590-9992) |
| Cervix | SEER recode: 27010 (excluding ICD O-3 codes: 9050-9055, 9140, 9590-9992) |
| Uterus | SEER recode: 27020, 27030 (excluding ICD O-3 codes: 9050-9055, 9140, 9590-9992) |
| Ovary | SEER recode: 27040(excluding ICD O-3 codes: 9050-9055, 9140, 9590-9992) |
| Prostate | SEER recode: 28010 (excluding ICD O-3 codes: 9050-9055, 9140, 9590-9992) |
| Testis | SEER recode: 28020 (excluding ICD O-3 codes: 9050-9055, 9140, 9590-9992) |
| Penis | SEER recode: 28030 (excluding ICD O-3 codes: 9050-9055, 9140, 9590-9992) |

Abbreviations: ICD – International Classification of Diseases, ICD O-3 – International Classification of Diseases for Oncology, third edition, NOS – Not otherwise specified, CNS – Central nervous system

¥ includes acute monocytic leukemia.

Table S1-2. Demographic information for the Marines/Navy personnel full cohort at risk during the follow-up period

| Factor | Camp Lejeune N=208,063 (47.9%) | | Camp Pendleton (ref) N=225,999 (52.1%) | | Total N=434,062 | |
|---|---|---|---|---|---|---|
| Male | 199,285 | (95.8%) | 219,217 | (97.0%) | 418,502 | (96.4%) |
| Female | 8,778 | (4.2%) | 6,782 | (3.0%) | 15,560 | (3.6%) |
| White | 154,093 | (74.1%) | 179,869 | (79.6%) | 333,962 | (76.9%) |
| African American | 49,258 | (23.7%) | 35,922 | (15.9%) | 85,180 | (19.6%) |
| Other or unknown race | 4,712 | (2.3%) | 10,208 | (4.5%) | 14,920 | (3.4%) |
| Rank E1 – E4 | 151,647 | (72.9%) | 158,747 | (70.2%) | 310,394 | (71.5%) |
| Rank E5 – E9 | 44,796 | (21.5%) | 51,346 | (22.7%) | 96,142 | (22.1%) |
| WO or CO | 11,620 | (5.6%) | 15,906 | (7.0%) | 27,526 | (6.3%) |
| Not a high school graduate | 33,717 | (16.2%) | 40,037 | (17.7%) | 73,754 | (17.0%) |
| High school graduate | 163,855 | (78.8%) | 171,200 | (75.8%) | 335,055 | (77.2%) |
| College graduate | 10,491 | (5.0%) | 14,762 | (6.5%) | 25,253 | (5.8%) |
| Age at start of follow-up (1/1/1996) | | | | | | |
| Mean (years) | 37.3 | | 37.8 | | 37.6 | |
| Median (years) | 37 | | 37 | | 37 | |
| Age at end of follow-up (12/31/2017 or date of death) | | | | | | |
| Mean | 58.3 | | 58.8 | | 58.5 | |
| Median | 58 | | 58 | | 58 | |
| Age ≥60 years | 85,395 | (40.5%) | 94,385 | (42.1%) | 179,780 | (41.4%) |
| Age >69 years | 8,739 | (4.2%) | 11,964 | (5.3%) | 20,703 | (4.8%) |
| Died during 1/2/1996 – 12/31/2017 | 24,689 | (11.9%) | 27,451 | (12.1%) | 52,140 | (12.0%) |
| Length of follow-up (years | | | | | | |
| Mean (years) | 20.1 | | 20.1 | | 20.1 | |
| Median (years) | 21 | | 21 | | 21 | |
| Total person-years of follow-up | 4,176,744 | | 4,533,157 | | 8,709,901 | |

| Cancers | Camp Lejeune | Camp Pendleton | Total |
|---|---|---|---|
| Total number of malignancies (including bladder cancer in situ | 23,131 | 23,581 | 46,712 |
| Total number of individuals with any malignancy or bladder cancer in situ | 21,013 | 21,500 | 42,513 |

Abbreviations: E1 – E4: private to corporal;
E5 – E9: sergeant to sergeant major;
WO: warrant officer; CO: commissioned officer

The table does not include aggregate cancer data obtained from the West Virginia and Kansas cancer registries.

Table S1-3. Standardized incidence rates and Poisson regression results: full cohort of Marines/Navy personnel

| CANCER | Camp Lejeune N | SIR | 95% CI | Camp Pendleton N | SIR | 95% CI | RR (CL vs CP)* | |
|---|---|---|---|---|---|---|---|---|
| Oral Cavity and Pharynx | 1,234 | 1.14 | (1.08, 1.21) | 1,372 | 1.13 | (1.07, 1.19) | 1.04 | (0.98, 1.11) |
| Esophagus | 377 | 0.98 | (0.89, 1.08) | 407 | 0.94 | (0.85, 1.03) | 1.08 | (0.99, 1.18) |
| Stomach | 344 | 0.86 | (0.77, 0.95) | 391 | 0.88 | (0.79, 0.97) | 0.98 | (0.85, 1.12) |
| Liver and bile duct | 631 | 1.01 | (0.93, 1.09) | 763 | 1.11 | (1.03, 1.19) | 0.94 | (0.86, 1.02) |
| Pancreas | 547 | 0.96 | (0.88, 1.04) | 581 | 0.91 | (0.84, 0.99) | 1.05 | (0.95, 1.18) |
| Gallbladder | 18 | 0.66 | (0.35, 0.96) | 24 | 0.78 | (0.47, 1.10) | 0.83 | (0.61, 1.13) |
| Larynx | 401 | 1.08 | (0.98, 1.19) | 361 | 0.89 | (0.80, 0.99) | 1.26 | (1.15, 1.39) |
| Lung and Bronchus | 3,030 | 1.04 | (1.00, 1.08) | 3,084 | 0.95 | (0.91, 0.98) | 1.13 | (1.07, 1.20) |
| Melanoma | 1,580 | 1.45 | (1.38, 1.52) | 1,909 | 1.44 | (1.37, 1.50) | 1.01 | (0.96, 1.07) |
| Urinary Bladder | 982 | 0.98 | (0.92, 1.05) | 1,109 | 0.92 | (0.87, 0.98) | 1.06 | (0.96, 1.16) |
| Kidney and Renal Pelvis | 1,224 | 1.04 | (0.98, 1.10) | 1,303 | 1.00 | (0.94, 1.05) | 1.05 | (0.96, 1.14) |
| Brain and CNS | 631 | 1.71 | (1.57, 1.84) | 662 | 1.56 | (1.44, 1.68) | 1.09 | (0.99, 1.19) |
| Thyroid | 372 | 0.87 | (0.78, 0.95) | 377 | 0.78 | (0.71, 0.86) | 1.10 | (0.97, 1.24) |
| NHL | 922 | 0.88 | (0.83, 0.94) | 1,010 | 0.86 | (0.80, 0.91) | 1.03 | (0.96, 1.12) |
| Multiple Myeloma | 318 | 0.89 | (0.79, 0.98) | 339 | 0.89 | (0.80, 0.99) | 0.98 | (0.86, 1.11) |
| Leukemias | 554 | 0.90 | (0.82, 0.97) | 604 | 0.85 | (0.78, 0.92) | 1.05 | (0.94, 1.17) |
| Colon and rectum | 1,924 | 0.82 | (0.78, 0.85) | 2,146 | 0.82 | (0.78, 0.85) | 0.99 | (0.92, 1.07) |
| Colon | 1,209 | 0.81 | (0.76, 0.85) | 1,373 | 0.83 | (0.79, 0.87) | 0.97 | (0.88, 1.06) |
| Rectum | 733 | 0.86 | (0.79, 0.92) | 803 | 0.83 | (0.77, 0.89) | 1.02 | (0.94, 1.11) |
| Anus | 105 | 0.98 | (0.79, 1.17) | 138 | 1.23 | (1.02, 1.43) | 0.82 | (0.70, 0.96) |
| Soft Tissue Sarcoma | 169 | 0.95 | (0.80, 1.09) | 167 | 0.85 | (0.72, 0.98) | 1.12 | (0.96, 1.30) |
| Hodgkin lymphoma | 163 | 1.02 | (0.86, 1.18) | 156 | 0.92 | (0.77, 1.06) | 1.13 | (0.95, 1.33) |
| ALL | 32 | 0.83 | (0.54, 1.11) | 34 | 0.78 | (0.52, 1.05) | 1.03 | (0.83, 1.29) |
| CLL | 214 | 0.92 | (0.80, 1.05) | 270 | 0.99 | (0.87, 1.11) | 0.91 | (0.79, 1.04) |
| AML | 172 | 1.02 | (0.87, 1.18) | 146 | 0.76 | (0.64, 0.88) | 1.34 | (1.14, 1.57) |
| CML | 69 | 0.72 | (0.55, 0.89) | 88 | 0.82 | (0.65, 0.99) | 0.89 | (0.73, 1.08) |
| Mesothelioma | 28 | 0.71 | (0.44, 0.97) | 33 | 0.69 | (0.45, 0.92) | 1.00 | (0.80, 1.26) |
| Breast Cancer - male | 51 | 0.89 | (0.64, 1.13) | 40 | 0.64 | (0.44, 0.83) | 1.39 | (1.05, 1.85) |
| Breast Cancer - female | 398 | 1.17 | (1.06, 1.29) | 332 | 1.18 | (1.05, 1.31) | 0.99 | (0.89, 1.10) |
| Prostate | 5,894 | 0.95 | (0.92, 0.97) | 6,075 | 0.89 | (0.86, 0.91) | 1.04 | (0.95, 1.13) |
| Testis | 213 | 0.82 | (0.71, 0.93) | 264 | 0.89 | (0.79, 1.00) | 0.92 | (0.81, 1.05) |
| Cervix | 24 | 0.87 | (0.52, 1.21) | 20 | 0.95 | (0.53, 1.36) | 0.92 | (0.67, 1.27) |
| Uterus | 46 | 0.77 | (0.55, 0.99) | 62 | 1.18 | (0.89, 1.47) | 0.66 | (0.52, 0.83) |
| Ovary | 27 | 0.95 | (0.59, 1.31) | 26 | 1.07 | (0.66, 1.49) | 0.91 | (0.64, 1.28) |

Abbreviations: CP - Camp Pendleton; CL – Camp Lejeune; SIR - standardized incidence ratios; RR – risk ratio; CI – confidence interval; CNS – central nervous system; NHL – non-Hodgkin lymphoma; ALL – acute lymphocytic leukemia; CLL – chronic lymphocytic leukemia; AML – acute myeloid leukemia; CML – chronic myeloid leukemia

* Poisson regression controlling for sex, race and 5-year age groups

SIRs calculated relative to sex, race and five-year age-specific cancer incidence statistics for 1999-2017 for the United States and Puerto Rico from the CDC WONDER.

Includes cancer cases from the aggregate data provided by the West Virginia and Kansas cancer registries.

Table S1-4. Marines/Navy personnel full cohort comparison of base location at Camp Lejeune vs Camp Pendleton

| Cancer Outcome | Camp Lejeune Cases | Unadjusted HR | (95% CI) | Adjusted HR | (95% CI) | Camp Pendleton Cases |
|---|---|---|---|---|---|---|
| Malignant cancers (and bladder in-situ) | 21,013 | 1.07 | (1.05, 1.09) | 1.04 | (1.02, 1.06) | 21,500 |
| Oral Cavity and Pharynx | 1,193 | 1.00 | (0.93, 1.08) | 1.02 | (0.94, 1.11) | 1,297 |
| Oropharynx | 699 | 1.04 | (0.93, 1.15) | 1.06 | (0.96, 1.18) | 735 |
| Hypopharynx | 59 | 0.91 | (0.65, 1.29) | 0.89 | (0.63, 1.26) | 71 |
| Nasopharynx | 39 | 0.90 | (0.59, 1.38) | 0.94 | (0.61, 1.45) | 47 |
| Oral cavity only | 222 | 0.96 | (0.80, 1.15) | 0.98 | (0.82, 1.18) | 251 |
| Overlapping/other | 83 | 1.14 | (0.84, 1.55) | 1.16 | (0.85, 1.58) | 79 |
| Squamous cell oral cancer | 1,061 | 1.02 | (0.94, 1.11) | 1.04 | (0.96, 1.13) | 1,187 |
| Esophagus | 381 | 1.07 | (0.93, 1.23) | 1.07 | (0.93, 1.24) | 390 |
| Adenocarcinoma | 235 | 1.00 | (0.84, 1.20) | 1.06 | (0.89, 1.26) | 256 |
| Squamous cell | 108 | 1.20 | (0.91, 1.58) | 1.09 | (0.83, 1.43) | 99 |
| Stomach | 349 | 1.00 | (0.86, 1.15) | 0.96 | (0.83, 1.12) | 383 |
| Liver and bile duct | 634 | 0.92 | (0.83, 1.02) | 0.92 | (0.83, 1.02) | 755 |
| Gallbladder | 18 | 0.90 | (0.49, 1.69) | 0.84 | (0.45, 1.57) | 22 |
| Pancreas | 558 | 1.07 | (0.95, 1.20) | 1.04 | (0.93, 1.17) | 571 |
| Larynx | 375 | 1.24 | (1.07, 1.43) | 1.21 | (1.04, 1.40) | 332 |
| Lung and Bronchus | 3,083 | 1.15 | (1.10, 1.21) | 1.13 | (1.07, 1.19) | 2,939 |
| Large cell | 89 | 1.17 | (0.87, 1.58) | 1.14 | (0.85, 1.55) | 83 |
| Small cell | 412 | 1.14 | (1.00, 1.31) | 1.14 | (1.00, 1.31) | 396 |
| Non-small cell | 338 | 1.21 | (1.04, 1.42) | 1.18 | (1.01, 1.38) | 305 |
| Squamous cell | 768 | 1.26 | (1.14, 1.40) | 1.23 | (1.11, 1.37) | 672 |
| Adenocarcinoma | 1,214 | 1.12 | (1.03, 1.21) | 1.08 | (1.00, 1.17) | 1,195 |
| Colon and Rectum | 1,799 | 1.01 | (0.95, 1.08) | 0.98 | (0.92, 1.05) | 1,948 |
| Adenocarcinoma | 1,562 | 0.99 | (0.93, 1.06) | 0.97 | (0.91, 1.04) | 1,719 |
| Colon | 1,119 | 0.99 | (0.91, 1.07) | 0.95 | (0.87, 1.03) | 1,240 |
| Rectum only | 571 | 1.07 | (0.95, 1.20) | 1.05 | (0.93, 1.18) | 584 |

| Cancer Outcome | Camp Lejeune Cases | Unadjusted HR | (95% CI) | Adjusted HR | (95% CI) | Camp Pendleton Cases |
|---|---|---|---|---|---|---|
| Rectosigmoid Junction | 143 | 0.94 | (0.75, 1.17) | 0.95 | (0.76, 1.19) | 166 |
| Small Intestine | 110 | 0.83 | (0.65, 1.07) | 0.79 | (0.62, 1.02) | 144 |
| Anus | 83 | 0.88 | (0.66, 1.18) | 0.85 | (0.64, 1.14) | 102 |
| Urinary Bladder (malignant and in-situ) | 1,001 | 1.03 | (0.94, 1.12) | 1.05 | (0.96, 1.14) | 1,072 |
| Papillary Transitional Cell Carcinoma | 712 | 1.06 | (0.96, 1.18) | 1.09 | (0.98, 1.21) | 736 |
| Non-papillary Transitional Cell Carcinoma | 257 | 0.99 | (0.83, 1.17) | 0.99 | (0.83, 1.17) | 287 |
| Urothelial | 969 | 1.04 | (0.95, 1.14) | 1.06 | (0.97, 1.16) | 1,023 |
| Bladder – malignant | 497 | 0.98 | (0.87, 1.10) | 0.99 | (0.87, 1.11) | 559 |
| Bladder – in-situ | 524 | 1.08 | (0.95, 1.21) | 1.11 | (0.98, 1.25) | 535 |
| Kidney and Renal Pelvis | 1,211 | 1.06 | (0.98, 1.14) | 1.04 | (0.96, 1.12) | 1,251 |
| Renal cell and clear cell carcinoma | 879 | 0.99 | (0.90, 1.09) | 1.00 | (0.91, 1.09) | 967 |
| Renal cell carcinoma, NOS | 448 | 1.12 | (0.98, 1.28) | 1.09 | (0.96, 1.25) | 434 |
| Clear cell only | 437 | 0.88 | (0.78, 1.00) | 0.92 | (0.81, 1.04) | 540 |
| Papillary | 162 | 1.30 | (1.03, 1.63) | 1.11 | (0.89, 1.40) | 137 |
| Brain and other CNS | 351 | 1.03 | (0.89, 1.19) | 1.05 | (0.91, 1.22) | 367 |
| Gliomas | 317 | 1.06 | (0.90, 1.23) | 1.08 | (0.93, 1.26) | 324 |
| Soft Tissue Sarcoma | 171 | 1.15 | (0.93, 1.43) | 1.14 | (0.92, 1.41) | 161 |
| Melanoma | 1,048 | 0.97 | (0.90, 1.06) | 1.05 | (0.96, 1.14) | 1,174 |
| Thyroid | 371 | 1.07 | (0.93, 1.24) | 1.08 | (0.93, 1.25) | 374 |
| Mesothelioma | 28 | 0.94 | (0.57, 1.55) | 0.93 | (0.56, 1.53) | 33 |
| Leukemias | 566 | 1.05 | (0.93, 1.18) | 1.05 | (0.93, 1.18) | 589 |
| Lymphoid cancers | 1,686 | 1.04 | (0.97, 1.11) | 1.02 | (0.96, 1.09) | 1,765 |
| Hodgkin lymphoma | 164 | 1.13 | (0.91, 1.41) | 1.11 | (0.89, 1.39) | 156 |
| Non-Hodgkin lymphoma | 937 | 1.05 | (0.96, 1.15) | 1.05 | (0.96, 1.15) | 971 |
| Mantle Cell | 51 | 1.21 | (0.81, 1.80) | 1.25 | (0.84, 1.87) | 46 |
| Follicular | 206 | 1.07 | (0.88, 1.30) | 1.12 | (0.92, 1.36) | 208 |
| Diffuse Large B-cell | 291 | 1.00 | (0.85, 1.17) | 1.01 | (0.86, 1.19) | 315 |
| Burkitt | 21 | 1.12 | (0.61, 2.07) | 1.26 | (0.68, 2.34) | 20 |
| Marginal Zone B-cell | 72 | 1.33 | (0.95, 1.88) | 1.33 | (0.94, 1.89) | 59 |
| Multiple Myeloma | 325 | 1.08 | (0.93, 1.26) | 0.99 | (0.84, 1.15) | 329 |

| Cancer Outcome | Camp Lejeune Cases | Unadjusted HR | (95% CI) | Adjusted HR | (95% CI) | Camp Pendleton Cases |
|---|---|---|---|---|---|---|
| Acute lymphocytic leukemia | 33 | 1.03 | (0.64, 1.67) | 1.03 | (0.64, 1.67) | 34 |
| Chronic lymphocytic leukemia | 219 | 0.90 | (0.75, 1.07) | 0.90 | (0.75, 1.07) | 267 |
| Myeloid cancers (including polycythemia vera, myelodysplastic and myeloproliferative syndromes) | 434 | 1.17 | (1.02, 1.34) | 1.17 | (1.02, 1.34) | 434 |
| Myeloid cancers (including myelodysplastic and myeloproliferative syndromes) | 352 | 1.17 | (1.01, 1.35) | 1.16 | (1.00, 1.35) | 354 |
| Acute myeloid leukemia[¥] | 174 | 1.33 | (1.07, 1.66) | 1.34 | (1.07, 1.67) | 142 |
| Chronic myeloid leukemia | 70 | 0.87 | (0.64, 1.20) | 0.87 | (0.64, 1.20) | 87 |
| Myelodysplastic and myeloproliferative syndromes | 126 | 1.24 | (0.97, 1.59) | 1.23 | (0.96, 1.58) | 124 |
| Polycythemia Vera | 84 | 1.17 | (0.86, 1.60) | 1.23 | (0.90, 1.68) | 78 |
| Female Breast | 316 | 1.07 | (0.91, 1.26) | 1.07 | (0.90, 1.26) | 256 |
| Ductal carcinoma | 237 | 1.12 | (0.92, 1.36) | 1.11 | (0.91, 1.35) | 182 |
| Lobular carcinoma | 23 | 0.68 | (0.40, 1.18) | 0.75 | (0.43, 1.30) | 30 |
| Duct-lobular carcinoma | 17 | 1.28 | (0.60, 2.73) | 1.39 | (0.64, 2.99) | 11 |
| Male Breast | 42 | 1.28 | (0.82, 2.00) | 1.24 | (0.79, 1.93) | 36 |
| Cervix | 24 | 0.90 | (0.50, 1.63) | 0.89 | (0.49, 1.62) | 20 |
| Uterus | 44 | 0.64 | (0.43, 0.95) | 0.64 | (0.43, 0.95) | 61 |
| Ovary | 24 | 0.90 | (0.51, 1.58) | 0.90 | (0.51, 1.60) | 24 |
| Prostate | 6,055 | 1.14 | (1.10, 1.18) | 1.05 | (1.01, 1.09) | 5,887 |
| Testis | 212 | 0.87 | (0.73, 1.04) | 0.91 | (0.76, 1.09) | 263 |
| Penis | 32 | 1.46 | (0.86, 2.48) | 1.49 | (0.87, 2.53) | 24 |

Abbreviations: HR – hazard ratio; CI – confidence interval; NOS-;CNS-

[¥] includes acute monocytic leukemia

HRs adjusted for sex, race, rank and education level; age was the time variable

Supplemental File 2

Figure 1: Smoking prevalence required to fully explain the HR of 1.08 in the Marines/Navy personnel sub-group analysis of COPD mortality as an underlying cause and base location, assuming RRs for smoking and COPD between 3.0 and 5.5.

**Multidimensional HR CL-COPD Relationship Adjusted for smoking**

| p(smoking+\|CL+)* | P(smoking+\|CL-)# | RR(smoking-COPD): | 3 | 3.25 | 3.5 | 3.75 | 4 | 4.25 | 4.5 | 4.75 | 5 | 5.25 | 5.5 |
|---|---|---|---|---|---|---|---|---|---|---|---|---|---|
| 0.45 | 0.44 | | 1.07 | 1.07 | 1.07 | 1.07 | 1.07 | 1.07 | 1.07 | 1.07 | 1.07 | 1.07 | 1.07 |
| 0.47 | 0.45 | | 1.06 | 1.06 | 1.06 | 1.06 | 1.06 | 1.05 | 1.05 | 1.05 | 1.05 | 1.05 | 1.05 |
| 0.49 | 0.46 | | 1.05 | 1.05 | 1.05 | 1.04 | 1.04 | 1.04 | 1.04 | 1.04 | 1.04 | 1.04 | 1.04 |
| 0.51 | 0.47 | | 1.04 | 1.04 | 1.04 | 1.03 | 1.03 | 1.03 | 1.03 | 1.03 | 1.03 | 1.02 | 1.02 |
| 0.53 | 0.48 | | 1.03 | 1.03 | 1.02 | 1.02 | 1.02 | 1.02 | 1.02 | 1.01 | 1.01 | 1.01 | 1.01 |
| 0.55 | 0.49 | | 1.02 | 1.02 | 1.01 | 1.01 | 1.01 | 1.01 | 1.00 | 1.00 | 1.00 | 1.00 | 1.00 |
| 0.57 | 0.50 | | 1.01 | 1.01 | 1.00 | 1.00 | 1.00 | 1.00 | 0.99 | 0.99 | 0.99 | 0.99 | 0.99 |
| 0.59 | 0.51 | | 1.00 | 1.00 | 1.00 | 0.99 | 0.99 | 0.99 | 0.98 | 0.98 | 0.98 | 0.98 | 0.98 |
| 0.61 | 0.52 | | 0.99 | 0.99 | 0.99 | 0.98 | 0.98 | 0.98 | 0.97 | 0.97 | 0.97 | 0.97 | 0.97 |
| 0.63 | 0.53 | | 0.99 | 0.98 | 0.98 | 0.97 | 0.97 | 0.97 | 0.96 | 0.96 | 0.96 | 0.96 | 0.96 |
| 0.65 | 0.54 | | 0.98 | 0.97 | 0.97 | 0.97 | 0.96 | 0.96 | 0.96 | 0.95 | 0.95 | 0.95 | 0.95 |

* Prevalence of smoking at Camp Lejeune
### Prevalence of smoking at Camp Pendleton

For smoking to fully account for the COPD HR of 1.08, the smoking prevalence difference between Camp Lejeune and Camp Pendleton would be 6%, corresponding to 55% prevalence at Camp Lejeune and 49% prevalence at Camp Pendleton (i.e., the prevalence difference resulting in the HR=1.00).

HR – hazard ratio
RR – risk ratio
COPD – Chronic obstructive pulmonary disease

Figure 2: Impact of adjusting for a smoking prevalence difference of 6% between CL and CP on the HR of 1.16 for lung cancer in the Marines/Navy personnel subgroup analysis of base location, and the smoking prevalence difference necessary to fully explain the HR of 1.16 for lung cancer. The RRs for smoking and lung cancer are assumed to be between 7.0 and 12.0.

**Multidimensional HR CL-lung cancer Relationship Adjusted for smoking**

| p(smoking+\|CL+)* | p(smoking+\|CL-)# | RR(smoking-lung cancer): | 7 | 7.5 | 8 | 8.5 | 9 | 9.5 | 10 | 10.5 | 11 | 11.5 | 12 |
|---|---|---|---|---|---|---|---|---|---|---|---|---|---|
| 0.45 | 0.42 | | 1.11 | 1.11 | 1.11 | 1.10 | 1.10 | 1.10 | 1.10 | 1.10 | 1.10 | 1.10 | 1.10 |
| 0.47 | 0.43 | | 1.09 | 1.09 | 1.09 | 1.09 | 1.09 | 1.09 | 1.08 | 1.08 | 1.08 | 1.08 | 1.08 |
| 0.49 | 0.44 | | 1.08 | 1.07 | 1.07 | 1.07 | 1.07 | 1.07 | 1.07 | 1.07 | 1.07 | 1.07 | 1.06 |
| 0.51 | 0.45 | | 1.06 | 1.06 | 1.06 | 1.06 | 1.05 | 1.05 | 1.05 | 1.05 | 1.05 | 1.05 | 1.05 |
| 0.53 | 0.46 | | 1.05 | 1.05 | 1.04 | 1.04 | 1.04 | 1.04 | 1.04 | 1.04 | 1.04 | 1.03 | 1.03 |
| 0.55 | 0.47 | | 1.03 | 1.03 | 1.03 | 1.03 | 1.03 | 1.03 | 1.02 | 1.02 | 1.02 | 1.02 | 1.02 |
| 0.57 | 0.48 | | 1.02 | 1.02 | 1.02 | 1.02 | 1.01 | 1.01 | 1.01 | 1.01 | 1.01 | 1.01 | 1.01 |
| 0.59 | 0.49 | | 1.01 | 1.01 | 1.01 | 1.00 | 1.00 | 1.00 | 1.00 | 1.00 | 1.00 | 0.99 | 0.99 |
| 0.61 | 0.50 | | 1.00 | 1.00 | 0.99 | 0.99 | 0.99 | 0.99 | 0.99 | 0.99 | 0.98 | 0.98 | 0.98 |
| 0.63 | 0.51 | | 0.99 | 0.99 | 0.98 | 0.98 | 0.98 | 0.98 | 0.98 | 0.97 | 0.97 | 0.97 | 0.97 |
| 0.65 | 0.52 | | 0.98 | 0.98 | 0.97 | 0.97 | 0.97 | 0.97 | 0.97 | 0.96 | 0.96 | 0.96 | 0.96 |

* Prevalence of smoking at Camp Lejeune
### Prevalence of smoking at Camp Pendleton
Abbreviations: HR – hazard ratio, RR – risk ratio, CL – Camp Lejeune, CP – Camp Pendleton

First highlighting: reduction in the HR for lung cancer by adjusting for a 6% difference in smoking prevalence between Camp Lejeune and Camp Pendleton. The HR of 1.16 for lung cancer would decrease to 1.05.

Second highlighting: Prevalence difference in smoking between the two bases to fully explain the lung cancer HR of 1.16 (i.e., the prevalence difference resulting in the HR=1.00). The prevalence difference would need to be at least 10%.

Figure 3: Impact of adjusting for a smoking prevalence difference of 6% between CL and CP on the HR of 1.21 for laryngeal cancer in the Marines/Navy personnel subgroup analysis of base location, and the smoking prevalence difference necessary to fully explain the HR of 1.21 for laryngeal cancer. The RRs for smoking and laryngeal cancer are assumed to be between 7.0 and 12.0.

**Multidimensional HR CL-laryngeal cancer Relationship Adjusted for smoking**

| p(smoking+\|CL+)* | P(smoking+\|CL-)# | RR(smoking-laryngeal cancer): | 7 | 7.5 | 8 | 8.5 | 9 | 9.5 | 10 | 10.5 | 11 | 11.5 | 12 |
|---|---|---|---|---|---|---|---|---|---|---|---|---|---|
| 0.45 | 0.44 | | 1.19 | 1.19 | 1.19 | 1.19 | 1.19 | 1.19 | 1.19 | 1.19 | 1.19 | 1.19 | 1.19 |
| 0.47 | 0.45 | | 1.18 | 1.17 | 1.17 | 1.17 | 1.17 | 1.17 | 1.17 | 1.17 | 1.17 | 1.17 | 1.17 |
| 0.49 | 0.46 | | 1.16 | 1.16 | 1.16 | 1.15 | 1.15 | 1.15 | 1.15 | 1.15 | 1.15 | 1.15 | 1.15 |
| 0.51 | 0.47 | | 1.14 | 1.14 | 1.14 | 1.14 | 1.14 | 1.14 | 1.14 | 1.13 | 1.13 | 1.13 | 1.13 |
| 0.53 | 0.48 | | 1.13 | 1.12 | 1.12 | 1.12 | 1.12 | 1.12 | 1.12 | 1.12 | 1.12 | 1.12 | 1.12 |
| 0.55 | 0.49 | | 1.11 | 1.11 | 1.11 | 1.11 | 1.11 | 1.10 | 1.10 | 1.10 | 1.10 | 1.10 | 1.10 |
| 0.57 | 0.50 | | 1.10 | 1.10 | 1.09 | 1.09 | 1.09 | 1.09 | 1.09 | 1.09 | 1.09 | 1.09 | 1.08 |

**Multidimensional HR CL-laryngeal cancer Relationship Adjusted for smoking**

| p(smoking+\|CL+)* | P(smoking+\|CL-)# | RR(smoking-laryngeal cancer): | 7 | 7.5 | 8 | 8.5 | 9 | 9.5 | 10 | 10.5 | 11 | 11.5 | 12 |
|---|---|---|---|---|---|---|---|---|---|---|---|---|---|
| 0.50 | 0.42 | | 1.07 | 1.06 | 1.06 | 1.06 | 1.06 | 1.06 | 1.05 | 1.05 | 1.05 | 1.05 | 1.05 |
| 0.52 | 0.43 | | 1.05 | 1.05 | 1.05 | 1.05 | 1.04 | 1.04 | 1.04 | 1.04 | 1.04 | 1.04 | 1.03 |
| 0.54 | 0.44 | | 1.04 | 1.04 | 1.04 | 1.03 | 1.03 | 1.03 | 1.03 | 1.03 | 1.02 | 1.02 | 1.02 |
| 0.56 | 0.45 | | 1.03 | 1.03 | 1.02 | 1.02 | 1.02 | 1.02 | 1.01 | 1.01 | 1.01 | 1.01 | 1.01 |
| 0.58 | 0.46 | | 1.02 | 1.01 | 1.01 | 1.01 | 1.01 | 1.00 | 1.00 | 1.00 | 1.00 | 1.00 | 1.00 |
| 0.60 | 0.47 | | 1.01 | 1.00 | 1.00 | 1.00 | 1.00 | 0.99 | 0.99 | 0.99 | 0.99 | 0.99 | 0.99 |
| 0.62 | 0.48 | | 1.00 | 0.99 | 0.99 | 0.99 | 0.99 | 0.98 | 0.98 | 0.98 | 0.98 | 0.98 | 0.97 |

* Prevalence of smoking at Camp Lejeune
### Prevalence of smoking at Camp Pendleton

Abbreviations: HR – hazard ratio, RR – risk ratio, CL – Camp Lejeune, CP – Camp Pendleton

The first table indicates the reduction in the HR for laryngeal cancer by adjusting for a 6% difference in smoking prevalence between Camp Lejeune and Camp Pendleton (highlighted). The HR of 1.21 for laryngeal cancer would decrease to between 1.10 and 1.11.

The second table indicates the prevalence difference required to fully explain the laryngeal cancer HR of 1.21, (i.e., the prevalence difference resulting in the HR=1.00, highlighted). The prevalence difference would need to be at least 12%.

Figure 4: Impact of adjusting the esophageal cancer HR of 1.27 by smoking assuming a 6% smoking prevalence difference between the Camp Lejeune and Camp Pendleton subgroup of Marines/Navy personnel. The RRs for smoking and esophageal cancer are assumed to be between 1.3 and 4.5.

**Multidimensional HR CL-esophagus Relationship Adjusted for smoking**

<table>
<tr><th>p(smoking+|CL+)*</th><th>P(smoking+|CL-)#</th><th>RR(smoking-esophagus):</th><th>1.3</th><th>1.5</th><th>1.8</th><th>2</th><th>2.3</th><th>2.5</th><th>2.8</th><th>3</th><th>3.5</th><th>4</th><th>4.5</th></tr>
<tr><td>0.45</td><td>0.44</td><td></td><td>1.27</td><td>1.27</td><td>1.26</td><td>1.26</td><td>1.26</td><td>1.26</td><td>1.26</td><td>1.26</td><td>1.26</td><td>1.25</td><td>1.25</td></tr>
<tr><td>0.47</td><td>0.45</td><td></td><td>1.26</td><td>1.26</td><td>1.26</td><td>1.25</td><td>1.25</td><td>1.25</td><td>1.25</td><td>1.24</td><td>1.24</td><td>1.24</td><td>1.24</td></tr>
<tr><td>0.49</td><td>0.46</td><td></td><td>1.26</td><td>1.26</td><td>1.25</td><td>1.25</td><td>1.24</td><td>1.24</td><td>1.23</td><td>1.23</td><td>1.23</td><td>1.22</td><td>1.22</td></tr>
<tr><td>0.51</td><td>0.47</td><td></td><td>1.26</td><td>1.25</td><td>1.24</td><td>1.24</td><td>1.23</td><td>1.23</td><td>1.22</td><td>1.22</td><td>1.22</td><td>1.21</td><td>1.21</td></tr>
<tr><td>0.53</td><td>0.48</td><td></td><td>1.25</td><td>1.25</td><td>1.24</td><td>1.23</td><td>1.22</td><td>1.22</td><td>1.21</td><td>1.21</td><td>1.20</td><td>1.20</td><td>1.19</td></tr>
<tr><td>0.55</td><td>0.49</td><td></td><td>1.25</td><td>1.24</td><td>1.23</td><td>1.22</td><td>1.21</td><td>1.21</td><td>1.20</td><td>1.20</td><td>1.19</td><td>1.18</td><td>1.18</td></tr>
<tr><td>0.57</td><td>0.50</td><td></td><td>1.25</td><td>1.24</td><td>1.22</td><td>1.21</td><td>1.20</td><td>1.20</td><td>1.19</td><td>1.19</td><td>1.18</td><td>1.17</td><td>1.17</td></tr>
<tr><td>0.59</td><td>0.51</td><td></td><td>1.25</td><td>1.23</td><td>1.22</td><td>1.21</td><td>1.20</td><td>1.19</td><td>1.18</td><td>1.18</td><td>1.17</td><td>1.16</td><td>1.15</td></tr>
<tr><td>0.61</td><td>0.52</td><td></td><td>1.24</td><td>1.23</td><td>1.21</td><td>1.20</td><td>1.19</td><td>1.18</td><td>1.17</td><td>1.17</td><td>1.16</td><td>1.15</td><td>1.14</td></tr>
<tr><td>0.63</td><td>0.53</td><td></td><td>1.24</td><td>1.22</td><td>1.20</td><td>1.19</td><td>1.18</td><td>1.17</td><td>1.16</td><td>1.16</td><td>1.15</td><td>1.14</td><td>1.13</td></tr>
<tr><td>0.65</td><td>0.54</td><td></td><td>1.24</td><td>1.22</td><td>1.20</td><td>1.19</td><td>1.17</td><td>1.16</td><td>1.16</td><td>1.15</td><td>1.14</td><td>1.13</td><td>1.12</td></tr>
</table>

* Prevalence of smoking at Camp Lejeune
### Prevalence of smoking at Camp Pendleton

The reduction in the HR for esophageal cancer by adjusting for a 6% difference in smoking prevalence between Camp Lejeune and Camp Pendleton is highlighted. The HR of 1.27 for esophageal cancer would decrease to between 1.18 and 1.25.

Abbreviations: HR – hazard ratio, RR – risk ratio, CL – Camp Lejeune, CP – Camp Pendleton

Figure 5. Alcohol use prevalence required to fully explain the HR of 0.93 in the Marines/Navy personnel subgroup analysis of chronic liver disease mortality as an underlying cause and base location. The RRs for alcohol consumption and chronic liver disease mortality are assumed to be between 2.5 and 10.0.

**Multidimensional HR CL-chronic liver disease mortality Relationship Adjusted for alcohol use**

| p(alcohol use+\|CL+)* | p(alcohol use+\|CL-)# | RR(alcohol use-chronic liver disease mortality): | 2.5 | 3 | 3.5 | 4 | 5 | 6 | 6.5 | 7 | 8 | 9 | 10 |
|---|---|---|---|---|---|---|---|---|---|---|---|---|---|
| 0.67 | 0.69 | | 0.94 | 0.94 | 0.95 | 0.95 | 0.95 | 0.95 | 0.95 | 0.95 | 0.95 | 0.95 | 0.95 |
| 0.67 | 0.70 | | 0.95 | 0.95 | 0.95 | 0.96 | 0.96 | 0.96 | 0.96 | 0.96 | 0.96 | 0.96 | 0.96 |
| 0.67 | 0.71 | | 0.96 | 0.96 | 0.96 | 0.97 | 0.97 | 0.97 | 0.97 | 0.97 | 0.97 | 0.97 | 0.98 |
| 0.67 | 0.72 | | 0.96 | 0.97 | 0.97 | 0.97 | 0.98 | 0.98 | 0.98 | 0.98 | 0.99 | 0.99 | 0.99 |
| 0.67 | 0.73 | | 0.97 | 0.98 | 0.98 | 0.98 | 0.99 | 0.99 | 0.99 | 0.99 | 1.00 | 1.00 | 1.00 |
| 0.67 | 0.74 | | 0.98 | 0.98 | 0.99 | 0.99 | 1.00 | 1.00 | 1.00 | 1.01 | 1.01 | 1.01 | 1.01 |
| 0.67 | 0.75 | | 0.98 | 0.99 | 1.00 | 1.00 | 1.01 | 1.01 | 1.02 | 1.02 | 1.02 | 1.02 | 1.02 |
| 0.67 | 0.76 | | 0.99 | 1.00 | 1.01 | 1.01 | 1.02 | 1.02 | 1.03 | 1.03 | 1.03 | 1.03 | 1.04 |
| 0.67 | 0.77 | | 1.00 | 1.01 | 1.02 | 1.02 | 1.03 | 1.03 | 1.04 | 1.04 | 1.04 | 1.05 | 1.05 |
| 0.67 | 0.78 | | 1.00 | 1.02 | 1.02 | 1.03 | 1.04 | 1.05 | 1.05 | 1.05 | 1.05 | 1.06 | 1.06 |
| 0.67 | 0.79 | | 1.01 | 1.02 | 1.03 | 1.04 | 1.05 | 1.06 | 1.06 | 1.06 | 1.07 | 1.07 | 1.07 |

To fully explain the HR of 0.93 for chronic liver disease mortality and base location, the prevalence difference between CL and CP would range between 6% and 10% (highlighted).

Alcohol use and liver cirrhosis mortality RRs: 2.65 for 25 g/day, 6.83 for 50 g/day, and 16.38 for 100 g/day.
Llamosas-Falcon L, Probst C, Buckler C, Jiang H et al. How does alcohol use impact morbidity and mortality of liver cirrhosis? A systematic review and dose-response meta-analysis. Hepatology International 08 September 2023 online ahead of print.

* Prevalence of alcohol use at Camp Lejeune
\# Prevalence of alcohol use at Camp Pendleton

Abbreviations: HR – hazard ratio, RR – risk ratio, CL – Camp Lejeune, CP – Camp Pendleton

Figure 6. Alcohol use prevalence required to fully explain the hazard ratio of 0.88 in the Marines/Navy personnel subgroup analysis of chronic liver disease mortality as a contributing cause and base location.

| Multidimensional HR CL-chronic liver disease mortality Relationship Adjusted for alcohol use | | | | | | | | | | | | | |
|---|---|---|---|---|---|---|---|---|---|---|---|---|---|
| **p(alcohol use+\|CL+)*** | **P(alcohol use+\|CL-)#** | **RR(alcohol use-chronic liver disease mortality):** | **2.5** | **3** | **3.5** | **4** | **5** | **6** | **6.5** | **7** | **8** | **9** | **10** |
| **0.67** | **0.74** | | 0.92 | 0.93 | 0.94 | 0.94 | 0.95 | 0.95 | 0.95 | 0.95 | 0.95 | 0.96 | 0.96 |
| **0.67** | **0.75** | | 0.93 | 0.94 | 0.94 | 0.95 | 0.95 | 0.96 | 0.96 | 0.96 | 0.96 | 0.97 | 0.97 |
| **0.67** | **0.76** | | 0.94 | 0.95 | 0.95 | 0.96 | 0.96 | 0.97 | 0.97 | 0.97 | 0.98 | 0.98 | 0.98 |
| **0.67** | **0.77** | | 0.94 | 0.95 | 0.96 | 0.97 | 0.97 | 0.98 | 0.98 | 0.98 | 0.99 | 0.99 | 0.99 |
| **0.67** | **0.78** | | 0.95 | 0.96 | 0.97 | 0.97 | 0.98 | 0.99 | 0.99 | 0.99 | 1.00 | 1.00 | 1.00 |
| **0.67** | **0.79** | | 0.96 | 0.97 | 0.98 | 0.98 | 0.99 | 1.00 | 1.00 | 1.00 | 1.01 | 1.01 | 1.01 |
| **0.67** | **0.80** | | 0.96 | 0.98 | 0.99 | 0.99 | 1.00 | 1.01 | 1.01 | 1.01 | 1.02 | 1.02 | 1.02 |
| **0.67** | **0.81** | | 0.97 | 0.98 | 0.99 | 1.00 | 1.01 | 1.02 | 1.02 | 1.03 | 1.03 | 1.03 | 1.04 |
| **0.67** | **0.82** | | 0.98 | 0.99 | 1.00 | 1.01 | 1.02 | 1.03 | 1.03 | 1.04 | 1.04 | 1.04 | 1.05 |
| **0.67** | **0.83** | | 0.98 | 1.00 | 1.01 | 1.02 | 1.03 | 1.04 | 1.04 | 1.05 | 1.05 | 1.06 | 1.06 |
| **0.67** | **0.84** | | 0.99 | 1.01 | 1.02 | 1.03 | 1.04 | 1.05 | 1.05 | 1.06 | 1.06 | 1.07 | 1.07 |

To fully explain the HR of 0.88 for chronic liver disease mortality and base location, the prevalence difference between CL and CP would range between 11% and 16% (highlighted).

* Prevalence of alcohol use at Camp Lejeune
### Prevalence of alcohol use at Camp Pendleton

Abbreviations: HR – hazard ratio, RR – risk ratio, CL – Camp Lejeune, CP – Camp Pendleton

Figure 7. Impact of adjusting for alcohol use prevalence difference of 10% between CL and CP on the HR of 1.47 for squamous cell esophageal cancer in the Marines/Navy personnel subgroup analysis of base location. The RRs for alcohol use and squamous cell esophageal cancer are assumed to be between 1.25 and 5.25.

**Multidimensional HR CL-sq. esophageal cancer Relationship Adjusted for alcohol use**

| p(alcohol use+\|CL+) | p(alcohol use+\|CL-) | RR(alcohol use-sq. esophageal cancer): | 1.25 | 1.5 | 1.75 | 2 | 2.5 | 3 | 3.5 | 4 | 4.5 | 5 | 5.25 |
|---|---|---|---|---|---|---|---|---|---|---|---|---|---|
| 0.67 | 0.68 | | 1.48 | 1.48 | 1.48 | 1.48 | 1.49 | 1.49 | 1.49 | 1.49 | 1.49 | 1.49 | 1.49 |
| 0.67 | 0.69 | | 1.48 | 1.49 | 1.49 | 1.49 | 1.50 | 1.50 | 1.50 | 1.50 | 1.51 | 1.51 | 1.51 |
| 0.67 | 0.70 | | 1.48 | 1.49 | 1.50 | 1.50 | 1.51 | 1.51 | 1.52 | 1.52 | 1.52 | 1.52 | 1.52 |
| 0.67 | 0.71 | | 1.49 | 1.50 | 1.50 | 1.51 | 1.52 | 1.53 | 1.53 | 1.53 | 1.54 | 1.54 | 1.54 |
| 0.67 | 0.72 | | 1.49 | 1.50 | 1.51 | 1.52 | 1.53 | 1.54 | 1.54 | 1.55 | 1.55 | 1.56 | 1.56 |
| 0.67 | 0.73 | | 1.49 | 1.51 | 1.52 | 1.53 | 1.54 | 1.55 | 1.56 | 1.56 | 1.57 | 1.57 | 1.57 |
| 0.67 | 0.74 | | 1.50 | 1.51 | 1.53 | 1.54 | 1.55 | 1.56 | 1.57 | 1.58 | 1.58 | 1.59 | 1.59 |
| 0.67 | 0.75 | | 1.50 | 1.52 | 1.53 | 1.55 | 1.56 | 1.58 | 1.59 | 1.59 | 1.60 | 1.60 | 1.61 |
| 0.67 | 0.76 | | 1.50 | 1.52 | 1.54 | 1.55 | 1.57 | 1.59 | 1.60 | 1.61 | 1.61 | 1.62 | 1.62 |
| 0.67 | 0.77 | | 1.51 | 1.53 | 1.55 | 1.56 | 1.59 | 1.60 | 1.61 | 1.62 | 1.63 | 1.64 | 1.64 |
| 0.67 | 0.78 | | 1.51 | 1.54 | 1.56 | 1.57 | 1.60 | 1.61 | 1.63 | 1.64 | 1.64 | 1.65 | 1.65 |

The increase in the HR for squamous cell esophageal cancer by adjusting for a 10% difference in alcohol use prevalence between Camp Lejeune and Camp Pendleton is highlighted. The HR of 1.47 would increase to between 1.51 and 1.64.

RR for squamous cell esophageal cancer and moderate alcohol use (12.5g/day – 50g/day) = 2.23
RR for squamous cell esophageal cancer and heavy alcohol use (>50g/day) = 4.95
Ref: 34.

Abbreviations: HR – hazard ratio, RR – risk ratio, CL – Camp Lejeune, CP – Camp Pendleton

Figure 8. Impact of adjusting for alcohol use prevalence difference of 10% between CL and CP on the HR of 1.27 for esophageal cancer in the Marines/Navy personnel subgroup analysis of base location. The RRs for alcohol use and esophageal cancer are assumed to be between 1.25 and 5.25.

**Multidimensional HR CL-esophageal cancer Relationship Adjusted for alcohol**

| p(alcohol+\|CL+) | p(alcohol+\|CL-) | RR(alcohol-esophageal cancer): | 1.25 | 1.5 | 1.75 | 2 | 2.5 | 3 | 3.5 | 4 | 4.5 | 5 | 5.25 |
|---|---|---|---|---|---|---|---|---|---|---|---|---|---|
| 0.67 | 0.68 | | 1.27 | 1.28 | 1.28 | 1.28 | 2.50 | 1.28 | 1.28 | 1.28 | 1.28 | 1.28 | 1.29 |
| 0.67 | 0.69 | | 1.28 | 1.28 | 1.28 | 1.29 | 2.75 | 1.29 | 1.29 | 1.30 | 1.30 | 1.30 | 1.30 |
| 0.67 | 0.70 | | 1.28 | 1.29 | 1.29 | 1.29 | 1.30 | 1.30 | 1.31 | 1.31 | 1.31 | 1.31 | 1.31 |
| 0.67 | 0.71 | | 1.28 | 1.29 | 1.30 | 1.30 | 1.31 | 1.31 | 1.32 | 1.32 | 1.32 | 1.33 | 1.33 |
| 0.67 | 0.72 | | 1.28 | 1.29 | 1.30 | 1.31 | 1.32 | 1.33 | 1.33 | 1.33 | 1.34 | 1.34 | 1.34 |
| 0.67 | 0.73 | | 1.29 | 1.30 | 1.31 | 1.32 | 1.33 | 1.34 | 1.34 | 1.35 | 1.35 | 1.35 | 1.36 |
| 0.67 | 0.74 | | 1.29 | 1.30 | 1.32 | 1.32 | 1.34 | 1.35 | 1.35 | 1.36 | 1.36 | 1.37 | 1.37 |
| 0.67 | 0.75 | | 1.29 | 1.31 | 1.32 | 1.33 | 1.35 | 1.36 | 1.37 | 1.37 | 1.38 | 1.38 | 1.38 |
| 0.67 | 0.76 | | 1.30 | 1.31 | 1.33 | 1.34 | 1.36 | 1.37 | 1.38 | 1.39 | 1.39 | 1.40 | 1.40 |
| 0.67 | 0.77 | | 1.30 | 1.32 | 1.33 | 1.35 | 1.37 | 1.38 | 1.39 | 1.40 | 1.40 | 1.41 | 1.41 |
| 0.67 | 0.78 | | 1.30 | 1.32 | 1.34 | 1.35 | 1.38 | 1.39 | 1.40 | 1.41 | 1.42 | 1.42 | 1.43 |

The increase in the HR for esophageal cancer by adjusting for a 10% difference in alcohol use prevalence between Camp Lejeune and Camp Pendleton is highlighted. The HR of 1.27 would increase to between 1.30 and 1.41.

RRs for alcohol consumption and esophageal cancer were assumed to be similar to the RRs for squamous cell esophageal cancer. Kunzmann AT, Coleman HG, Huang WY, Berndt SI. The association of lifetime alcohol use with mortality and cancer risk in older adults: A cohort study. PLoS Med 2018;15(6): e1002585 (Supplemental table 3).

Abbreviations: HR – hazard ratio, RR – risk ratio, CL – Camp Lejeune, CP – Camp Pendleton

Figure 9. Impact of adjusting for alcohol use prevalence difference of 10% between CL and CP on the HR of 1.21 for laryngeal cancer in the Marines/Navy personnel subgroup analysis of base location. RRs for alcohol use and laryngeal cancer are assumed to be between 1.1 and 3.0.

**Multidimensional HR CL-laryngeal cancer Relationship Adjusted for alcohol use**

| p(alcohol use+\|CL+) | p(alcohol use+\|CL-) | RR(alcohol use-laryngeal cancer): | 1.1 | 1.3 | 1.5 | 1.7 | 1.9 | 2.1 | 2.3 | 2.5 | 2.7 | 2.9 | 3 |
|---|---|---|---|---|---|---|---|---|---|---|---|---|---|
| 0.67 | 0.68 | | 1.21 | 1.22 | 1.22 | 1.22 | 1.22 | 1.22 | 1.22 | 1.22 | 1.22 | 1.22 | 1.22 |
| 0.67 | 0.69 | | 1.22 | 1.22 | 1.22 | 1.22 | 1.23 | 1.23 | 1.23 | 1.23 | 1.23 | 1.23 | 1.23 |
| 0.67 | 0.70 | | 1.22 | 1.22 | 1.23 | 1.23 | 1.23 | 1.24 | 1.24 | 1.24 | 1.24 | 1.24 | 1.24 |
| 0.67 | 0.71 | | 1.22 | 1.23 | 1.23 | 1.24 | 1.24 | 1.24 | 1.25 | 1.25 | 1.25 | 1.25 | 1.25 |
| 0.67 | 0.72 | | 1.22 | 1.23 | 1.24 | 1.24 | 1.25 | 1.25 | 1.26 | 1.26 | 1.26 | 1.26 | 1.27 |
| 0.67 | 0.73 | | 1.22 | 1.23 | 1.24 | 1.25 | 1.25 | 1.26 | 1.26 | 1.27 | 1.27 | 1.27 | 1.28 |
| 0.67 | 0.74 | | 1.22 | 1.23 | 1.25 | 1.25 | 1.26 | 1.27 | 1.27 | 1.28 | 1.28 | 1.28 | 1.29 |
| 0.67 | 0.75 | | 1.22 | 1.24 | 1.25 | 1.26 | 1.27 | 1.27 | 1.28 | 1.29 | 1.29 | 1.29 | 1.30 |
| 0.67 | 0.76 | | 1.22 | 1.24 | 1.25 | 1.27 | 1.27 | 1.28 | 1.29 | 1.30 | 1.30 | 1.30 | 1.31 |
| 0.67 | 0.77 | | 1.22 | 1.24 | 1.26 | 1.27 | 1.28 | 1.29 | 1.30 | 1.30 | 1.31 | 1.31 | 1.32 |
| 0.67 | 0.78 | | 1.23 | 1.25 | 1.26 | 1.28 | 1.29 | 1.30 | 1.31 | 1.31 | 1.32 | 1.32 | 1.33 |

The increase in the HR for laryngeal cancer by adjusting for a 10% difference in alcohol use prevalence between Camp Lejeune and Camp Pendleton is highlighted. The HR of 1.21 would increase to between 1.22 and 1.32.

RR for laryngeal cancer and moderate alcohol use (12.5g/day – 50g/day) = 1.44
RR for laryngeal cancer and heavy alcohol use (>50g/day) = 2.65
Ref: 34.

Abbreviations: HR – hazard ratio, RR – risk ratio, CL – Camp Lejeune, CP – Camp Pendleton

Figure 10: Smoking prevalence difference between Camp Lejeune and Camp Pendleton workers required to fully explain the HR of 1.05 in the analysis of COPD as a contributing cause of death.

**Multidimensional HR CL-COPD Relationship Adjusted for smoking**

| p(smoking+\|CL+) | p(smoking+\|CL-) | RR(smoking-COPD): | 3 | 3.25 | 3.5 | 3.75 | 4 | 4.25 | 4.5 | 4.75 | 5 | 5.25 | 5.5 |
|---|---|---|---|---|---|---|---|---|---|---|---|---|---|
| **0.45** | **0.44** | | 1.04 | 1.04 | 1.04 | 1.04 | 1.04 | 1.04 | 1.04 | 1.04 | 1.04 | 1.04 | 1.04 |
| **0.47** | **0.45** | | 1.03 | 1.03 | 1.03 | 1.03 | 1.03 | 1.02 | 1.02 | 1.02 | 1.02 | 1.02 | 1.02 |
| **0.49** | **0.46** | | 1.02 | 1.02 | 1.02 | 1.01 | 1.01 | 1.01 | 1.01 | 1.01 | 1.01 | 1.01 | 1.01 |
| **0.51** | **0.47** | | 1.01 | 1.01 | 1.01 | 1.00 | 1.00 | 1.00 | 1.00 | 1.00 | 1.00 | 1.00 | 0.99 |
| **0.53** | **0.48** | | 1.00 | 1.00 | 1.00 | 0.99 | 0.99 | 0.99 | 0.99 | 0.99 | 0.98 | 0.98 | 0.98 |
| **0.55** | **0.49** | | 0.99 | 0.99 | 0.99 | 0.98 | 0.98 | 0.98 | 0.98 | 0.97 | 0.97 | 0.97 | 0.97 |
| **0.57** | **0.50** | | 0.98 | 0.98 | 0.98 | 0.97 | 0.97 | 0.97 | 0.97 | 0.96 | 0.96 | 0.96 | 0.96 |
| **0.59** | **0.51** | | 0.97 | 0.97 | 0.97 | 0.96 | 0.96 | 0.96 | 0.96 | 0.95 | 0.95 | 0.95 | 0.95 |
| **0.61** | **0.52** | | 0.97 | 0.96 | 0.96 | 0.95 | 0.95 | 0.95 | 0.95 | 0.94 | 0.94 | 0.94 | 0.94 |
| **0.63** | **0.53** | | 0.96 | 0.95 | 0.95 | 0.95 | 0.94 | 0.94 | 0.94 | 0.93 | 0.93 | 0.93 | 0.93 |
| **0.65** | **0.54** | | 0.95 | 0.95 | 0.94 | 0.94 | 0.93 | 0.93 | 0.93 | 0.93 | 0.92 | 0.92 | 0.92 |

* Prevalence of smoking at Camp Lejeune
### Prevalence of smoking at Camp Pendleton

For smoking to fully account for the COPD HR of 1.05, the smoking prevalence difference between Camp Lejeune and Camp Pendleton would be 4%, corresponding to 51% prevalence at Camp Lejeune and 47% prevalence at Camp Pendleton (i.e., the prevalence difference resulting in the HR=1.00, highlighted).

Abbreviations: HR – hazard ratio, RR – risk ratio, COPD – Chronic obstructive pulmonary disease

Figure 11: Impact of adjusting for a smoking prevalence difference of 4% between Camp Lejeune and Camp Pendleton workers on the HR of 1.15 for lung cancer, and the smoking prevalence difference necessary to fully explain the HR of 1.15 for lung cancer. The RRs for smoking and lung cancer are assumed to be between 7.0 and 12.0.

**Multidimensional HR CL-lung cancer Relationship Adjusted for smoking**

| **p(smoking+\|CL+)** | **p(smoking+\|CL-)** | **RR(smoking-lung cancer):** | **7** | **7.5** | **8** | **8.5** | **9** | **9.5** | **10** | **10.5** | **11** | **11.5** | **12** |
|---|---|---|---|---|---|---|---|---|---|---|---|---|---|
| **0.45** | **0.44** | | 1.13 | 1.13 | 1.13 | 1.13 | 1.13 | 1.13 | 1.13 | 1.13 | 1.13 | 1.13 | 1.13 |
| **0.47** | **0.45** | | 1.12 | 1.12 | 1.12 | 1.12 | 1.11 | 1.11 | 1.11 | 1.11 | 1.11 | 1.11 | 1.11 |
| **0.49** | **0.46** | | 1.10 | 1.10 | 1.10 | 1.10 | 1.10 | 1.10 | 1.10 | 1.10 | 1.09 | 1.09 | 1.09 |
| **0.51** | **0.47** | | 1.09 | 1.08 | 1.08 | 1.08 | 1.08 | 1.08 | 1.08 | 1.08 | 1.08 | 1.08 | 1.08 |
| **0.53** | **0.48** | | 1.07 | 1.07 | 1.07 | 1.07 | 1.07 | 1.06 | 1.06 | 1.06 | 1.06 | 1.06 | 1.06 |
| **0.55** | **0.49** | | 1.06 | 1.06 | 1.05 | 1.05 | 1.05 | 1.05 | 1.05 | 1.05 | 1.05 | 1.05 | 1.05 |
| **0.57** | **0.50** | | 1.04 | 1.04 | 1.04 | 1.04 | 1.04 | 1.04 | 1.03 | 1.03 | 1.03 | 1.03 | 1.03 |
| **0.59** | **0.51** | | 1.03 | 1.03 | 1.03 | 1.03 | 1.02 | 1.02 | 1.02 | 1.02 | 1.02 | 1.02 | 1.02 |
| **0.61** | **0.52** | | 1.02 | 1.02 | 1.02 | 1.01 | 1.01 | 1.01 | 1.01 | 1.01 | 1.01 | 1.01 | 1.01 |
| **0.63** | **0.53** | | 1.01 | 1.01 | 1.00 | 1.00 | 1.00 | 1.00 | 1.00 | 1.00 | 1.00 | 0.99 | 0.99 |
| **0.65** | **0.54** | | 1.00 | 1.00 | 0.99 | 0.99 | 0.99 | 0.99 | 0.99 | 0.99 | 0.98 | 0.98 | 0.98 |

* Prevalence of smoking at Camp Lejeune
### Prevalence of smoking at Camp Pendleton

First highlighting: reduction in the HR for lung cancer by adjusting for a 4% difference in smoking prevalence between Camp Lejeune and Camp Pendleton. The HR of 1.15 would decrease to between 1.08 and 1.09.

Second highlighting: Prevalence difference in smoking between the two bases to fully explain the lung cancer HR of 1.15 (i.e., the prevalence difference resulting in the HR=1.00). The prevalence difference would need to be at least 10%.

Abbreviations: HR – hazard ratio, RR – risk ratio, CL – Camp Lejeune, CP – Camp Pendleton

Figure 12: Impact of adjusting for a smoking prevalence difference of 4% between Camp Lejeune and Camp Pendleton workers on the HR for laryngeal cancer of 1.18. The RRs for smoking and laryngeal cancer are assumed to be between 7.0 and 12.0.

**Multidimensional HR CL-laryngeal cancer Relationship Adjusted for smoking**

| p(smoking+\|CL+) | p(smoking+\|CL-) | RR(smoking-laryngeal cancer): | 7 | 7.5 | 8 | 8.5 | 9 | 9.5 | 10 | 10.5 | 11 | 11.5 | 12 |
|---|---|---|---|---|---|---|---|---|---|---|---|---|---|
| **0.45** | **0.44** | | 1.17 | 1.16 | 1.16 | 1.16 | 1.16 | 1.16 | 1.16 | 1.16 | 1.16 | 1.16 | 1.16 |
| **0.47** | **0.45** | | 1.15 | 1.15 | 1.15 | 1.15 | 1.14 | 1.14 | 1.14 | 1.14 | 1.14 | 1.14 | 1.14 |
| **0.49** | **0.46** | | 1.13 | 1.13 | 1.13 | 1.13 | 1.13 | 1.13 | 1.13 | 1.12 | 1.12 | 1.12 | 1.12 |
| **0.51** | **0.47** | | 1.11 | 1.11 | 1.11 | 1.11 | 1.11 | 1.11 | 1.11 | 1.11 | 1.11 | 1.11 | 1.11 |
| **0.53** | **0.48** | | 1.10 | 1.10 | 1.10 | 1.10 | 1.09 | 1.09 | 1.09 | 1.09 | 1.09 | 1.09 | 1.09 |
| **0.55** | **0.49** | | 1.09 | 1.08 | 1.08 | 1.08 | 1.08 | 1.08 | 1.08 | 1.08 | 1.08 | 1.07 | 1.07 |
| **0.57** | **0.50** | | 1.07 | 1.07 | 1.07 | 1.07 | 1.07 | 1.06 | 1.06 | 1.06 | 1.06 | 1.06 | 1.06 |
| **0.59** | **0.51** | | 1.06 | 1.06 | 1.06 | 1.05 | 1.05 | 1.05 | 1.05 | 1.05 | 1.05 | 1.05 | 1.05 |
| **0.61** | **0.52** | | 1.05 | 1.05 | 1.04 | 1.04 | 1.04 | 1.04 | 1.04 | 1.04 | 1.03 | 1.03 | 1.03 |
| **0.63** | **0.53** | | 1.04 | 1.03 | 1.03 | 1.03 | 1.03 | 1.03 | 1.02 | 1.02 | 1.02 | 1.02 | 1.02 |
| **0.65** | **0.54** | | 1.03 | 1.02 | 1.02 | 1.02 | 1.02 | 1.01 | 1.01 | 1.01 | 1.01 | 1.01 | 1.01 |

* Prevalence of smoking at Camp Lejeune
### Prevalence of smoking at Camp Pendleton

Highlighting: reduction in the HR for laryngeal cancer by adjusting for a 4% difference in smoking prevalence between Camp Lejeune and Camp Pendleton. The HR of 1.18 would decrease to 1.11.

Note: To fully explain the laryngeal cancer HR of 1.18, the smoking prevalence difference between the two bases would be >11%.

Abbreviations: HR – hazard ratio, RR – risk ratio, CL – Camp Lejeune, CP – Camp Pendleton

Figure 13: Impact of adjusting for a smoking prevalence difference of 4% between Camp Lejeune and Camp Pendleton workers on the HR for oral cancers of 1.67. The RRs for smoking and oral cancers was assumed to be between 3.5 and 7.0.

**Multidimensional HR CL-oral cancers Relationship Adjusted for smoking**

| p(smoking+\|CL+) | p(smoking+\|CL-) | RR(smoking-oral cancers): | 3.5 | 3.75 | 4 | 4.25 | 4.5 | 4.75 | 5 | 5.5 | 6 | 6.5 | 7 |
|---|---|---|---|---|---|---|---|---|---|---|---|---|---|
| **0.45** | **0.44** | | 1.65 | 1.65 | 1.65 | 1.65 | 1.64 | 1.64 | 1.64 | 1.64 | 1.64 | 1.64 | 1.64 |
| **0.47** | **0.45** | | 1.63 | 1.63 | 1.63 | 1.62 | 1.62 | 1.62 | 1.62 | 1.62 | 1.62 | 1.62 | 1.61 |
| **0.49** | **0.46** | | 1.61 | 1.61 | 1.61 | 1.60 | 1.60 | 1.60 | 1.60 | 1.60 | 1.59 | 1.59 | 1.59 |
| **0.51** | **0.47** | | 1.59 | 1.59 | 1.59 | 1.59 | 1.58 | 1.58 | 1.58 | 1.58 | 1.57 | 1.57 | 1.57 |
| **0.53** | **0.48** | | 1.58 | 1.57 | 1.57 | 1.57 | 1.56 | 1.56 | 1.56 | 1.56 | 1.55 | 1.55 | 1.55 |
| **0.55** | **0.49** | | 1.56 | 1.56 | 1.55 | 1.55 | 1.55 | 1.54 | 1.54 | 1.54 | 1.53 | 1.53 | 1.53 |
| **0.57** | **0.50** | | 1.55 | 1.54 | 1.54 | 1.53 | 1.53 | 1.53 | 1.52 | 1.52 | 1.52 | 1.51 | 1.51 |
| **0.59** | **0.51** | | 1.53 | 1.53 | 1.52 | 1.52 | 1.51 | 1.51 | 1.51 | 1.50 | 1.50 | 1.49 | 1.49 |
| **0.61** | **0.52** | | 1.52 | 1.51 | 1.51 | 1.50 | 1.50 | 1.50 | 1.49 | 1.49 | 1.48 | 1.48 | 1.47 |
| **0.63** | **0.53** | | 1.51 | 1.50 | 1.49 | 1.49 | 1.49 | 1.48 | 1.48 | 1.47 | 1.47 | 1.46 | 1.46 |
| **0.65** | **0.54** | | 1.49 | 1.49 | 1.48 | 1.48 | 1.47 | 1.47 | 1.46 | 1.46 | 1.45 | 1.45 | 1.44 |

* Prevalence of smoking at Camp Lejeune
### Prevalence of smoking at Camp Pendleton

Highlighting: reduction in the HR for oral cancers by adjusting for a 4% difference in smoking prevalence between Camp Lejeune and Camp Pendleton. The HR of 1.67 would decrease to between 1.57 and 1.59.

Abbreviations: HR – hazard ratio, RR – risk ratio, CL – Camp Lejeune, CP – Camp Pendleton

Figure 14: Impact of adjusting for a smoking prevalence difference of 4% between Camp Lejeune and Camp Pendleton workers on the HR for kidney cancer of 1.12.

**Multidimensional HR CL-kidney cancer Relationship Adjusted for smoking**

| p(smoking+\|CL+) | p(smoking+\|CL-) | RR(smoking-kidney cancer): | 1.2 | 1.25 | 1.3 | 1.35 | 1.4 | 1.45 | 1.5 | 1.53 | 1.56 | 1.59 | 1.6 |
|---|---|---|---|---|---|---|---|---|---|---|---|---|---|
| 0.45 | 0.44 | | 1.12 | 1.12 | 1.12 | 1.12 | 1.12 | 1.12 | 1.11 | 1.11 | 1.11 | 1.11 | 1.11 |
| 0.47 | 0.45 | | 1.12 | 1.11 | 1.11 | 1.11 | 1.11 | 1.11 | 1.11 | 1.11 | 1.11 | 1.11 | 1.11 |
| 0.49 | 0.46 | | 1.11 | 1.11 | 1.11 | 1.11 | 1.11 | 1.11 | 1.11 | 1.11 | 1.10 | 1.10 | 1.10 |
| 0.51 | 0.47 | | 1.11 | 1.11 | 1.11 | 1.11 | 1.10 | 1.10 | 1.10 | 1.10 | 1.10 | 1.10 | 1.10 |
| 0.53 | 0.48 | | 1.11 | 1.11 | 1.11 | 1.10 | 1.10 | 1.10 | 1.10 | 1.10 | 1.10 | 1.09 | 1.09 |
| 0.55 | 0.49 | | 1.11 | 1.10 | 1.10 | 1.10 | 1.10 | 1.10 | 1.09 | 1.09 | 1.09 | 1.09 | 1.09 |
| 0.57 | 0.50 | | 1.11 | 1.10 | 1.10 | 1.10 | 1.09 | 1.09 | 1.09 | 1.09 | 1.09 | 1.08 | 1.08 |
| 0.59 | 0.51 | | 1.10 | 1.10 | 1.10 | 1.09 | 1.09 | 1.09 | 1.08 | 1.08 | 1.08 | 1.08 | 1.08 |
| 0.61 | 0.52 | | 1.10 | 1.10 | 1.09 | 1.09 | 1.09 | 1.08 | 1.08 | 1.08 | 1.08 | 1.08 | 1.08 |
| 0.63 | 0.53 | | 1.10 | 1.10 | 1.09 | 1.09 | 1.08 | 1.08 | 1.08 | 1.08 | 1.07 | 1.07 | 1.07 |
| 0.65 | 0.54 | | 1.10 | 1.09 | 1.09 | 1.08 | 1.08 | 1.08 | 1.07 | 1.07 | 1.07 | 1.07 | 1.07 |

* Prevalence of smoking at Camp Lejeune
### Prevalence of smoking at Camp Pendleton

Highlighting: reduction in the HR for kidney cancer by adjusting for a 4% difference in smoking prevalence between Camp Lejeune and Camp Pendleton. The HR of 1.12 would decrease between 1.10 and 1.11.

Note: To fully explain the kidney cancer HR of 1.12, the smoking prevalence difference between the two bases would have to be much greater than 11%.

Abbreviations: HR – hazard ratio, RR – risk ratio, CL – Camp Lejeune, CP – Camp Pendleton

Figure 15. Alcohol use prevalence required to fully explain the hazard ratio of 0.74 in the analysis of the base location of civilian workers and chronic liver disease mortality as an underlying cause. The RRs for alcohol use and chronic liver disease mortality were assumed to be between 2.5 and 10.0.

**Multidimensional HR CL-chronic liver disease mortality Relationship Adjusted for alcohol use**

| p(alcohol use+\|CL+)* | p(alcohol use+\|CL-)# | RR(alcohol use-chronic liver disease mortality): | 2.5 | 3 | 3.5 | 4 | 5 | 6 | 6.5 | 7 | 8 | 9 | 10 |
|---|---|---|---|---|---|---|---|---|---|---|---|---|---|
| 0.33 | 0.48 | | 0.86 | 0.88 | 0.90 | 0.91 | 0.94 | 0.96 | 0.96 | 0.97 | 0.98 | 0.99 | 1.00 |
| 0.33 | 0.49 | | 0.86 | 0.89 | 0.91 | 0.92 | 0.95 | 0.97 | 0.98 | 0.98 | 1.00 | 1.01 | 1.02 |
| 0.33 | 0.50 | | 0.87 | 0.90 | 0.92 | 0.94 | 0.96 | 0.98 | 0.99 | 1.00 | 1.01 | 1.02 | 1.03 |
| 0.33 | 0.51 | | 0.88 | 0.91 | 0.93 | 0.95 | 0.98 | 1.00 | 1.01 | 1.01 | 1.03 | 1.04 | 1.05 |
| 0.33 | 0.52 | | 0.89 | 0.92 | 0.94 | 0.96 | 0.99 | 1.01 | 1.02 | 1.03 | 1.04 | 1.06 | 1.07 |
| 0.33 | 0.53 | | 0.89 | 0.92 | 0.95 | 0.97 | 1.00 | 1.03 | 1.04 | 1.04 | 1.06 | 1.07 | 1.08 |
| 0.33 | 0.54 | | 0.90 | 0.93 | 0.96 | 0.98 | 1.01 | 1.04 | 1.05 | 1.06 | 1.08 | 1.09 | 1.10 |
| 0.33 | 0.55 | | 0.91 | 0.94 | 0.97 | 0.99 | 1.03 | 1.05 | 1.07 | 1.07 | 1.09 | 1.11 | 1.12 |
| 0.33 | 0.56 | | 0.92 | 0.95 | 0.98 | 1.00 | 1.04 | 1.07 | 1.08 | 1.09 | 1.11 | 1.12 | 1.13 |
| 0.33 | 0.57 | | 0.92 | 0.96 | 0.99 | 1.01 | 1.05 | 1.08 | 1.09 | 1.10 | 1.12 | 1.14 | 1.15 |
| 0.33 | 0.58 | | 0.93 | 0.97 | 1.00 | 1.03 | 1.07 | 1.10 | 1.11 | 1.12 | 1.14 | 1.15 | 1.17 |

* Prevalence of alcohol use at Camp Lejeune
### Prevalence of alcohol use at Camp Pendleton

To fully explain the HR of 0.74 for chronic liver disease mortality and base location, the prevalence difference between CL and CP would range between 15% and 25% (highlighted).

Alcohol consumption and liver cirrhosis mortality RRs: 2.65 for 25 g/day, 6.83 for 50 g/day, and 16.38 for 100 g/day. Llamosas-Falcon L, Probst C, Buckler C, Jiang H et al. How does alcohol use impact morbidity and mortality of liver cirrhosis? A systematic review and dose-response meta-analysis. Hepatology International 08 September 2023 online ahead of print.

Abbreviations: HR – hazard ratio, RR – risk ratio, CL – Camp Lejeune, CP – Camp Pendleton

Figure 16. Impact of adjusting for alcohol use prevalence difference of 15% between CL and CP on the HR of 1.19 for female breast cancer in the civilian workers analysis of base location. The RRs for alcohol use and female breast cancer were assumed to be between 1.1 and 1.6.

**Multidimensional HR CL-female breast cancer Relationship Adjusted for alcohol use**

| p(alcohol use+\|CL+)* | p(alcohol use+\|CL-)# | RR(alcohol use-female breast cancer): | 1.1 | 1.15 | 1.2 | 1.25 | 1.3 | 1.35 | 1.4 | 1.45 | 1.5 | 1.55 | 1.6 |
|---|---|---|---|---|---|---|---|---|---|---|---|---|---|
| 0.33 | 0.44 | | 1.20 | 1.20 | 1.21 | 1.22 | 1.22 | 1.23 | 1.23 | 1.24 | 1.24 | 1.25 | 1.25 |
| 0.33 | 0.45 | | 1.20 | 1.21 | 1.21 | 1.22 | 1.22 | 1.23 | 1.24 | 1.24 | 1.25 | 1.25 | 1.26 |
| 0.33 | 0.46 | | 1.20 | 1.21 | 1.21 | 1.22 | 1.23 | 1.23 | 1.24 | 1.25 | 1.25 | 1.26 | 1.26 |
| 0.33 | 0.47 | | 1.20 | 1.21 | 1.22 | 1.22 | 1.23 | 1.24 | 1.24 | 1.25 | 1.26 | 1.26 | 1.27 |
| 0.33 | 0.48 | | 1.20 | 1.21 | 1.22 | 1.23 | 1.23 | 1.24 | 1.25 | 1.26 | 1.26 | 1.27 | 1.27 |
| 0.33 | 0.49 | | 1.20 | 1.21 | 1.22 | 1.23 | 1.24 | 1.24 | 1.25 | 1.26 | 1.27 | 1.27 | 1.28 |
| 0.33 | 0.50 | | 1.20 | 1.21 | 1.22 | 1.23 | 1.24 | 1.25 | 1.26 | 1.26 | 1.27 | 1.28 | 1.29 |
| 0.33 | 0.51 | | 1.21 | 1.22 | 1.23 | 1.23 | 1.24 | 1.25 | 1.26 | 1.27 | 1.28 | 1.28 | 1.29 |
| 0.33 | 0.52 | | 1.21 | 1.22 | 1.23 | 1.24 | 1.25 | 1.26 | 1.26 | 1.27 | 1.28 | 1.29 | 1.30 |
| 0.33 | 0.53 | | 1.21 | 1.22 | 1.23 | 1.24 | 1.25 | 1.26 | 1.27 | 1.28 | 1.29 | 1.30 | 1.30 |
| 0.33 | 0.54 | | 1.21 | 1.22 | 1.23 | 1.24 | 1.25 | 1.26 | 1.27 | 1.28 | 1.29 | 1.30 | 1.31 |

* Prevalence of alcohol use at Camp Lejeune
\# Prevalence of alcohol use at Camp Pendleton

The increase in the HR for female breast cancer by adjusting for a 15% difference in alcohol use prevalence between Camp Lejeune and Camp Pendleton is highlighted. The HR of 1.19 would increase to between 1.20 and 1.27.

RR for female breast cancer and moderate alcohol use (12.5g/day – 50g/day) = 1.23
RR for female breast cancer and heavy alcohol use (>50g/day) = 1.61
Ref. 34

Abbreviations: HR – hazard ratio, RR – risk ratio, CL – Camp Lejeune, CP – Camp Pendleton

Figure 17. Impact of adjusting for alcohol use prevalence difference of 15% between CL and CP on the HR of 1.19 for laryngeal cancer in the civilian workers analysis of base location. The RRs for alcohol use and laryngeal cancer are assumed to be between 1.1 and 3.0.

**Multidimensional HR CL-laryngeal cancer Relationship Adjusted for alcohol use**

| p(alcohol use+\|CL+)* | p(alcohol use+\|CL-)[#] | RR(alcohol use-laryngeal cancer): | 1.1 | 1.3 | 1.5 | 1.7 | 1.9 | 2.1 | 2.3 | 2.5 | 2.7 | 2.9 | 3 |
|---|---|---|---|---|---|---|---|---|---|---|---|---|---|
| **0.33** | **0.45** | | 1.20 | 1.22 | 1.24 | 1.26 | 1.28 | 1.30 | 1.31 | 1.32 | 1.34 | 1.35 | 1.35 |
| **0.33** | **0.46** | | 1.20 | 1.22 | 1.25 | 1.27 | 1.29 | 1.31 | 1.32 | 1.34 | 1.35 | 1.36 | 1.37 |
| **0.33** | **0.47** | | 1.20 | 1.23 | 1.25 | 1.28 | 1.30 | 1.31 | 1.33 | 1.35 | 1.36 | 1.37 | 1.38 |
| **0.33** | **0.48** | | 1.20 | 1.23 | 1.26 | 1.28 | 1.30 | 1.32 | 1.34 | 1.36 | 1.37 | 1.39 | 1.39 |
| **0.33** | **0.49** | | 1.20 | 1.23 | 1.26 | 1.29 | 1.31 | 1.33 | 1.35 | 1.37 | 1.39 | 1.40 | 1.41 |
| **0.33** | **0.50** | | 1.20 | 1.24 | 1.27 | 1.30 | 1.32 | 1.34 | 1.36 | 1.38 | 1.40 | 1.42 | 1.42 |
| **0.33** | **0.51** | | 1.20 | 1.24 | 1.27 | 1.30 | 1.33 | 1.35 | 1.37 | 1.39 | 1.41 | 1.43 | 1.44 |
| **0.33** | **0.52** | | 1.20 | 1.24 | 1.28 | 1.31 | 1.34 | 1.36 | 1.39 | 1.41 | 1.43 | 1.44 | 1.45 |
| **0.33** | **0.53** | | 1.20 | 1.25 | 1.28 | 1.32 | 1.35 | 1.37 | 1.40 | 1.42 | 1.44 | 1.46 | 1.47 |
| **0.33** | **0.54** | | 1.21 | 1.25 | 1.29 | 1.32 | 1.35 | 1.38 | 1.41 | 1.43 | 1.45 | 1.47 | 1.48 |
| **0.33** | **0.55** | | 1.21 | 1.25 | 1.29 | 1.33 | 1.36 | 1.39 | 1.42 | 1.44 | 1.46 | 1.48 | 1.49 |

* Prevalence of alcohol use at Camp Lejeune
### Prevalence of alcohol use at Camp Pendleton

The increase in the HR for laryngeal cancer by adjusting for a 15% difference in alcohol use prevalence between Camp Lejeune and Camp Pendleton is highlighted. The HR of 1.19 would increase to between 1.20 and 1.39.

RR for laryngeal cancer and moderate alcohol use (12.5g/day – 50g/day) = 1.44
RR for laryngeal cancer and heavy alcohol use (>50g/day) = 2.65
Ref. 34

Abbreviations: HR – hazard ratio, RR – risk ratio, CL – Camp Lejeune, CP – Camp Pendleton

Figure 18. Impact of adjusting for alcohol use prevalence difference of 15% between CL and CP on the HR of 1.67 for oral cancers in the civilian workers analysis of base location. The RRs for alcohol use and oral cancers were assumed to be between 1.25 and 5.25.

| Multidimensional HR CL-oral cavity and pharynx Relationship Adjusted for alcohol | | | | | | | | | | | | | |
|---|---|---|---|---|---|---|---|---|---|---|---|---|---|
| p(alcohol+\|CL+)* | p(alcohol+\|CL-)# | RR(alcohol-oral cancers): | 1.25 | 1.5 | 1.75 | 2 | 2.5 | 3 | 3.5 | 4 | 4.5 | 5 | 5.25 |
| 0.33 | 0.43 | | 1.71 | 1.74 | 1.77 | 1.80 | 2.50 | 1.87 | 1.90 | 1.92 | 1.94 | 1.96 | 1.97 |
| 0.33 | 0.44 | | 1.71 | 1.75 | 1.78 | 1.81 | 2.75 | 1.89 | 1.92 | 1.95 | 1.97 | 1.99 | 2.00 |
| 0.33 | 0.45 | | 1.72 | 1.76 | 1.79 | 1.82 | 1.87 | 1.91 | 1.95 | 1.97 | 2.00 | 2.02 | 2.03 |
| 0.33 | 0.46 | | 1.72 | 1.76 | 1.80 | 1.83 | 1.89 | 1.93 | 1.97 | 2.00 | 2.02 | 2.04 | 2.05 |
| 0.33 | 0.47 | | 1.72 | 1.77 | 1.81 | 1.85 | 1.91 | 1.95 | 1.99 | 2.02 | 2.05 | 2.07 | 2.08 |
| 0.33 | 0.48 | | 1.73 | 1.78 | 1.82 | 1.86 | 1.92 | 1.97 | 2.01 | 2.05 | 2.08 | 2.10 | 2.11 |
| 0.33 | 0.49 | | 1.73 | 1.79 | 1.83 | 1.87 | 1.94 | 1.99 | 2.04 | 2.07 | 2.10 | 2.13 | 2.14 |
| 0.33 | 0.50 | | 1.74 | 1.79 | 1.84 | 1.88 | 1.96 | 2.01 | 2.06 | 2.10 | 2.13 | 2.16 | 2.17 |
| 0.33 | 0.51 | | 1.74 | 1.80 | 1.85 | 1.90 | 1.97 | 2.03 | 2.08 | 2.12 | 2.16 | 2.19 | 2.20 |
| 0.33 | 0.52 | | 1.74 | 1.81 | 1.86 | 1.91 | 1.99 | 2.05 | 2.11 | 2.15 | 2.19 | 2.22 | 2.23 |
| 0.33 | 0.53 | | 1.75 | 1.81 | 1.87 | 1.92 | 2.01 | 2.07 | 2.13 | 2.17 | 2.21 | 2.25 | 2.26 |

* Prevalence of alcohol use at Camp Lejeune
### Prevalence of alcohol use at Camp Pendleton

The increase in the HR for oral cancers by adjusting for a 15% difference in alcohol use prevalence between Camp Lejeune and Camp Pendleton is highlighted. The HR of 1.67 would increase to between 1.73 and 2.11.

Abbreviations: HR – hazard ratio, RR – risk ratio, CL – Camp Lejeune, CP – Camp Pendleton

RR for oral cancer and moderate alcohol use (12.5g/day – 50g/day) = 1.83.
RR for oral cancer and heavy alcohol use (>50g/day) = 5.13. Ref. 34

Table S2-1a. Hazard ratios for cause of death related to smoking or alcohol consumption for the subgroup of Marines/Navy personnel comparing CL versus CP.

| | Underlying cause | | | | Contributing cause | | | |
|---|---|---|---|---|---|---|---|---|
| Outcome | Total cases | Camp Lejeune (# cases) | Camp Pendleton (# cases) | Adjusted HR (95% CI) | Total cases | Camp Lejeune (# cases) | Camp Pendleton (# cases) | Adjusted HR (95% CI) |
| alcoholism | 544 | 242 | 302 | 0.90 (0.76, 1.07) | 2,413 | 1,072 | 1,341 | 0.90 (0.83, 0.98) |
| alcoholic liver disease | 901 | 381 | 520 | 0.86 (0.76, 0.99) | 1,208 | 506 | 702 | 0.84 (0.75, 0.95) |
| chronic liver disease | 1,389 | 614 | 775 | 0.93 (0.83, 1.03) | 2,310 | 996 | 1,314 | 0.88 (0.81, 0.96) |
| COPD | 632 | 312 | 320 | 1.08 (0.93, 1.27) | 1,700 | 809 | 891 | 1.02 (0.93, 1.12) |
| cardiovascular disease | 8,966 | 4,316 | 4,650 | 0.99 (0.95, 1.03) | 14,848 | 7,107 | 7,741 | 0.98 (0.95, 1.01) |

Abbreviations: CP - Camp Pendleton, CL – Camp Lejeune, HR – hazard ratio, CI – confidence interval, COPD – Chronic obstructive pulmonary disease

HRs adjusted for sex, race, rank and education level.

Table S2-1b. Hazard ratios for cause of death related to smoking or alcohol consumption for the civilian workers comparing CL versus CP.

| | Underlying cause | | | | | Contributing cause | | | | |
|---|---|---|---|---|---|---|---|---|---|---|
| Outcome | Total cases | Camp Lejeune (# cases) | Camp Pendleton (# cases) | Adjusted HR | (95% CI) | Total cases | Camp Lejeune (# cases) | Camp Pendleton (# cases) | Adjusted HR | (95% CI) |
| alcoholism | 17 | 7 | 10 | 0.62 | (0.23, 1.71) | 81 | 33 | 48 | 0.66 | (0.41, 1.05) |
| alcoholic liver disease | 49 | 16 | 33 | 0.54 | (0.29, 1.00) | 61 | 21 | 40 | 0.58 | (0.34, 1.01) |
| chronic liver disease | 87 | 36 | 51 | 0.74 | (0.48, 1.15) | 150 | 59 | 91 | 0.66 | (0.47, 0.92) |
| COPD | 384 | 171 | 213 | 0.91 | (0.74, 1.12) | 871 | 418 | 453 | 1.05 | (0.92, 1.20) |
| cardiovascular disease | 2,377 | 1,105 | 1,272 | 0.91 | (0.83, 0.99) | 3,831 | 1,785 | 2,046 | 0.90 | (0.84, 0.96) |

Abbreviations: CP - Camp Pendleton, CL – Camp Lejeune, HR – hazard ratio, CI – confidence interval, COPD – Chronic obstructive pulmonary disease

HRs adjusted for sex, race, blue collar work (y/n) and education level.

Table S2-2a. Hazard ratios comparing the subgroup of Camp Lejeune and Camp Pendleton Marines/Navy personnel, adjusted for nondifferential exposure misclassification.

| Sensitivity | Specificity | % false positive | Lung cancer | Laryngeal cancer | Esophageal cancer | AML |
|---|---|---|---|---|---|---|
| | | | 1.16£ | 1.21£ | 1.27£ | 1.38£ |
| 1.00 | 0.91 | 10% | 1.18 | 1.24 | 1.30 | 1.42 |
| 1.00 | 0.875 | 15% | 1.19 | 1.25 | 1.32 | 1.44 |
| 1.00 | 0.84 | 20% | 1.21 | 1.27 | 1.34 | 1.47 |
| 1.00 | 0.81 | 25% | 1.22 | 1.28 | 1.36 | 1.50 |

AML: acute myeloid leukemia

£: Adjusted Hazard Ratio from Table 4

Table S2-2b. Hazard ratios comparing the Camp Lejeune and Camp Pendleton workers, adjusted for nondifferential exposure misclassification.

| Sensitivity | Specificity | % false positive | Lung cancer | Laryngeal cancer | Oral cancers | Kidney cancer | Non-Hodgkin lymphoma | Female breast cancer |
|---|---|---|---|---|---|---|---|---|
| | | | 1.15£ | 1.18£ | 1.67£ | 1.12£ | 1.19£ | 1.19£ |
| 1.00 | 0.91 | 10% | 1.16 | 1.20 | 1.73 | 1.14 | 1.21 | 1.21 |
| 1.00 | 0.875 | 15% | 1.16 | 1.20 | 1.74 | 1.14 | 1.22 | 1.22 |
| 1.00 | 0.84 | 20% | 1.18 | 1.22 | 1.81 | 1.15 | 1.23 | 1.23 |
| 1.00 | 0.81 | 25% | 1.19 | 1.23 | 1.85 | 1.16 | 1.24 | 1.24 |

£: Adjusted Hazard Ratio from Table 5
